# Impact of the COVID-19 pandemic on tuberculosis control in Indonesia: a nationwide analysis of programme data and health system vulnerabilities

**DOI:** 10.1101/2023.02.09.23285740

**Authors:** Henry Surendra, Iqbal RF Elyazar, Evelyn Puspaningrum, Deddy Darmawan, Tiffany T Pakasi, Endang Lukitosari, Sulistyo Sulistyo, Shena M Deviernur, Ahmad Fuady, Guy Thwaites, Reinout van Crevel, Anuraj H Shankar, J Kevin Baird, Raph L Hamers

**Author notes:** **Corresponding author:** Henry Surendra PhD., Oxford University Clinical Research Unit Indonesia Faculty of Medicine, Universitas Indonesia, Jalan Salemba Raya No. 6, 10430 Jakarta, Indonesia.

## Abstract

**Background:** There are limited measures of the impact of the COVID-19 pandemic on tuberculosis (TB) control in high-burden countries like Indonesia.

**Methods:** We analysed district-level data of reported TB cases, treatment and deaths, COVID-19 incidence and mortality, health care capacity, economic status, education level, and public health development index from all 514 districts in Indonesia. We compared data before (2016-2019) and during (2020-2021) the pandemic.

**Findings:** Compared to the preceding year (2019), in the first pandemic year (2020) the TB case notification declined by 31% (from median 172 [IQR 129-244] in 2019 to 119 [IQR 87-170] in 2020 per 100,000 population; 565,669 vs 393,323 cases, respectively); mortality increased by 8% (from median 4·2 [IQR 2·0-7·4] to 5·0 (IQR 3·1-7·5) per 100,000 population; 13,059 vs 14,148 deaths, respectively); and the overall proportion of cases who started treatment declined by 7% (from 98% to 91%). The second pandemic year (2021) saw a partial recovery of case notifications (median 142 [IQR 99-204]; 473,006) and deaths (4·1 [IQR 2·5-6·8]; 12,016), but a persistently reduced treatment coverage (84%). Reductions in TB notifications between districts were associated with higher COVID-19 incidence and fewer per capita GeneXpert machines for TB diagnosis. Likewise, reduced TB treatment coverage was associated with fewer per capita doctors, and increased reported TB deaths was associated with fewer per capita primary health centres, lower per capita domestic expenditure and higher education.

**Interpretation:** The COVID-19 pandemic significantly, yet unevenly, impacted the national TB control programme across Indonesia, with the greatest impacts in districts with the least resilient health systems.

**Funding:** Wellcome Africa Asia Programme Vietnam

## Background

Coronavirus disease (COVID-19) caused by the SARS-CoV-2 virus, has resulted in over 6.8 million reported deaths as per February 9, 2023 ^1^, and the actual excess COVID-19 death toll may be triple that amount.^2^ The COVID-19 pandemic severely impacted almost every aspect of global health, including the delivery of essential services for other major health conditions such as tuberculosis (TB), the leading cause of death from a single infectious agent other than SARS-CoV-2.^3^ Mitigating the transmission, morbidity, and mortality of TB involves patient-centred approaches to accessible and high-quality TB diagnostic and treatment services.^4,5^ According to WHO estimates, TB case notifications declined from 7.1 million in 2019 to 5.8 million (18% decline) in 2020 ^6^, with pandemic-related disruptions in health services being especially exacerbated in low-and middle-income countries (LMIC) in sub-Saharan Africa ^7–11^ and Asia ^12,13^, because of the vulnerabilities in their health systems.^14^ Only three countries accounted for 67% of the global reduction in case reporting: India, Indonesia and the Philippines, each experiencing major COVID-19 epidemics through 2020. Global TB mortality increased for the first time in more than a decade; from an estimated 1.4 million deaths in 2019, 1.5 million in 2020, to 1.6 million in 2021.^15^ Reduced case-finding and hence treatment coverage during the past two years likely increased transmission rates and WHO modelling forecasted worsening TB incidence and mortality in the coming few years.^6^

Indonesia has the second-highest TB burden in the world ^6^, and reported the highest number of confirmed COVID-19 cases and deaths in the Southeast Asia region.^1^ A recent nationwide analysis of patient-level data from the National TB Control Programme (NTCP) in Indonesia reported that during the years immediately preceding the COVID-19 pandemic (2017-2019) notified cases of drug-susceptible TB increased from 429 219 to 523 614 (from 167 to 196 cases per 100 000), with stable treatment success rates and mortality.^16^ Despite significant progress in rolling out national health insurance towards achieving universal health coverage ^17,18^, the quality of health care remains highly variable across its decentralised public health system.^18^ The 2018 Public Health Development Index (PHDI), a composite indicator constructed by the Ministry of Health expressing coverage and equity in health services, and health status ^19^, ranged from 35% in Paniai district, Papua province to 75% in Gianyar district, Bali province. No studies have yet examined the impact of the COVID-19 pandemic on TB services and outcomes in Indonesia at the subnational level in association with key health system indicators.

Reports to date, including both the WHO Global TB Report ^15^ and the Global Burden of Disease Study ^20^, applied extrapolated and modelled TB burden estimates at national levels. However, rigorous large-scale, sub-national analyses of patient databases in high-burden TB countries are required to more fully understand the impacts of the COVID-19 pandemic on TB control, services and disease outcomes in the context of metrics of health system resilience.^12^ In the present study, we sought to conduct a comprehensive nationwide analysis of the intertwined COVID-19 and TB epidemics in Indonesia. We specifically sought to understand the impacts on the NTCP, by examining TB case notification, treatment and mortality rates before (2016-2019) and during the pandemic (2020-2021), in association with metrics of healthcare system resilience at the district level, across all 514 districts and 34 provinces of Indonesia.

## Methods

### Study design and participants

We conducted a comprehensive analysis of NTCP data to assess the impact of the COVID-19 pandemic on TB case notification, treatment coverage, and reported mortality and possible associations with health system factors in all 514 districts in Indonesia. The study included aggregated data reported by primary health centres, government hospitals as well as private healthcare facilities through the national tuberculosis information system (*Sistem Informasi Tuberkulosis*, SITB).^21^ The SITB is a mandatory case notification system for healthcare facilities to inform their district health office to support tuberculosis surveillance, contact investigation, outbreak management, and infection control.^22^

### Data collection

The study outcomes were annual number of new reported TB cases, treatment, and reported deaths. TB cases were defined as all children and adults clinically or microbiologically diagnosed and reported as TB cases, based on clinical scoring, smear microscopy, culture and/or molecular diagnostics (typically GeneXpert).^23^ TB treatment was defined as the number of TB cases that had initiated treatment. TB reported deaths were defined as the number of patients dying from any cause by the end of treatment.^23^ District-level quarterly aggregated data of notified TB cases, number of TB cases who started treatment, and number of deaths among TB treatment cases between 2016 and 2021, and number of health facilities with TB smear microscopy service, and number of health facilities with a TB GeneXpert machine in 2020 and 2021 were collected from the NTCP at the Ministry of Health of Indonesia.^21^ Each district had a TB programme officer responsible for data completion, verification, and submission of programme data to the provincial and national registries. Data on annual population numbers between 2016 and 2021 were collected from the National Bureau of Statistics Database.^24^

For explanatory variables, aggregated data of the monthly number of COVID-19 cases, and number of COVID-19 deaths among confirmed cases by district, from 1 March 2020 to 31 December 2021, were collected from the government COVID-19 database managed by the National COVID-19 Task Force.^25^ District-level data on the cumulative number of doctors, nurses, midwives, and hospitals in 2020 and 2021 were collected from the Ministry of Health records. District-level data on per capita domestic expenditure, life expectancy at birth, and average length of formal education were collected from the Human Development Index 2020 Report.^26^ District-level data on Public Health Development Index (PHDI) were collected from the 2018 PHDI report.^19^

### Statistical analysis

For outcomes, district-level case notification rate (annual number of all reported TB cases per 100 000 population), treatment coverage (annual proportion of TB cases who started treatment), and mortality rate (annual number of TB reported deaths per 100 000 population) were calculated. The case notification rate and mortality rates were summarised and plotted over time to illustrate trends from 2016 to 2021. Due to data limitations, only the annual treatment coverage from 2019 to 2021 were analysed. CNR, mortality rate, and treatment coverage were compared between the pandemic phase (2020 and 2021) and the pre-pandemic phase (2019), and expressed as case notification rate ratio, mortality rate ratio, and treatment coverage ratio, with their 95% confidence intervals (CI). Proportions of districts significantly impacted based on decrease in case notification rate (ratio and 95% CI<1), decrease in treatment coverage (ratio and 95% CI<1), and increase in mortality rate (ratio and 95% CI>1) were calculated for each province. The use of 2019 data as the reference was considered sufficient based on trend analysis presented in Supplementary Figure 1.

For explanatory variables, district-level COVID-19 incidence rate, COVID-19 mortality rate, numbers per 100 000 population of facilities with a GeneXpert machine, facilities with TB microscopy smear service, primary health centres, doctors, nurses, and midwives, per capita domestic expenditure (USD per capita), and mean duration of formal education (years) were calculated and categorised into quartiles. Descriptive statistics included summaries of medians (IQR: interquartile range) and Mann-Whitney test to compare medians, proportions for districts and chi-squared tests to compare characteristics between districts with heterogeneity in impact of the COVID-19 pandemic.

We used bivariable and multivariable mixed effects logistic regression models to determine factors associated with a significant decrease in TB case notification rate, increase in TB mortality rate, and decrease in treatment coverage, expressed as odds ratio (OR) with 95% CI. Province was treated as the random effect variable to adjust for clustering of observations within provinces. We did null model analysis (no predictor was added) and the results show variance of 37% for case notification, 61% for treatment coverage, and 59% for mortality (p<0.001 each), thus justified the use of the mixed effects models. All independent variables with p-value <0.10 in bivariable analysis were included in the multivariable models. Final model selection was informed by likelihood ratio tests. We set statistical significance at p<0.05, and all tests were two sided. All analyses were done in Stata/IC 15.1 (StataCorp, College Station, TX, USA). This study is reported as per Strengthening the Reporting of Observational Studies in Epidemiology (STROBE) guidelines.^27^

### Ethics

This study was approved by the Health Research Ethics Committee of National Institute of Health Research and Development, Ministry of Health of Indonesia (LB.02.01/2/KE.486/2021). The requirement for patient consent was waived as this was a secondary analysis of aggregated routine programme data with no personal identifiers.

## Results

Table 1 summarises the general profiles of the 514 districts analysed in this study. The cumulative burden of COVID-19 incidence and mortality rate as per 31 December 2021 was 139 (IQR: 59-280) and 4 (1-9) per 100 000 population, respectively. Health facilities with a GeneXpert machine was median 0.5 (IQR: 0.2-0.9) per 100 000 population, health facility with TB microscopy smear service was median 4 (IQR: 3-6) per 100 000 population, and the number of doctors was median 8 (IQR: 5-18) per 100 000 population. The per capita domestic expenditure was median USD 675 (IQR: 574-777).

**Table 1.**
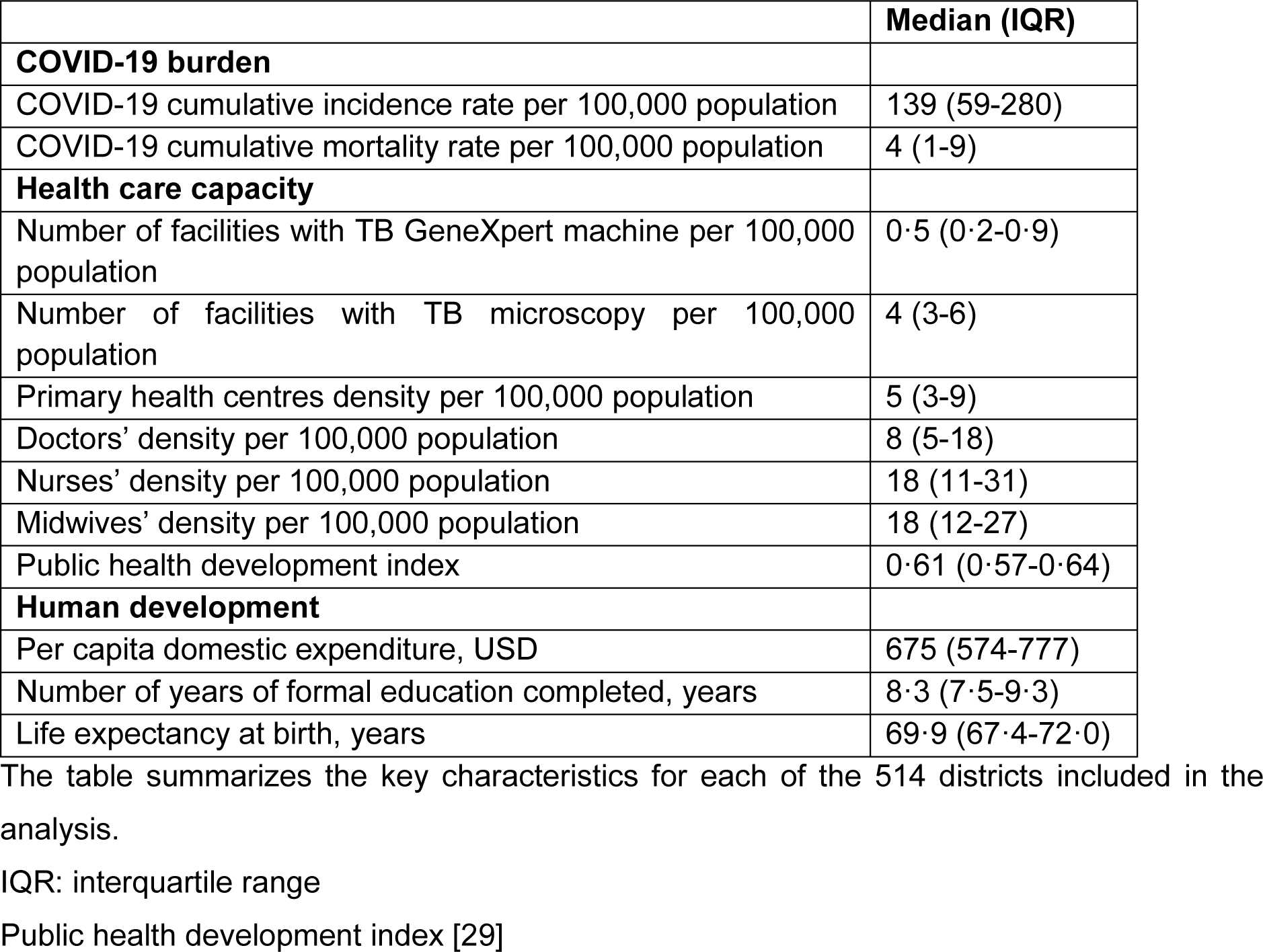
District profiles.

### Tuberculosis case notification rate and mortality rate before (2016-2019) and during the COVID-19 pandemic (2020-2021)

Table 2 summarises the number of reported TB cases and deaths, and changes in TB case notification rate and mortality rate from 2016 to 2021. The median case notification per quarter gradually increased from quarter 1 of 2016 through quarter 1 of 2020, sharply decreased in quarter 2 of 2020 (after the start of COVID-19 pandemic in Indonesia in early March 2020), and then increased from quarter 1 through 4 in 2021 (Supplementary Figure 2A). The median case notification rate in 2019 (before the COVID-19 pandemic), was 172 (1QR 129-244) per 100 000 population. The median case notification rate during the COVID-19 pandemic was 119 (IQR 87-170) in 2020 and 142 (IQR 99-204) in 2021. The total number of new TB cases decreased from 565 669 in 2019, to 393 323 (−31%) in 2020, and increased again to 473 006 (+20%) in 2021.

**Table 2.** Changes in tuberculosis case notification rate and mortality over time.

| Year | Total New TB cases | Case notification rate per 100,000 population |  |  | New TB deaths | Mortality rate per 100,000 population |  |  |
| --- | --- | --- | --- | --- | --- | --- | --- | --- |
|  |  | Median (IQR) | Median different (% change) | p value |  | Median (IQR) | Median different (% change) | p value |
| 2016 | 360,565 | 124 (88-174) | 0 | .. | 9,049 | 3.1 (1.5-5.8) | 0 | .. |
| 2017 | 436,180 | 142 (100-199) | +18 (14.5) | **0.0001** | 13,640 | 3.9 (1.6-6.5) | +0.8 (25.8) | **0.0190** |
| 2018 | 514,161 | 166 (116-227) | +24 (16.9) | **0.0001** | 12,442 | 4.2 (2.0-6.6) | +0.3 (7.7) | 0.2833 |
| 2019 | 565,669 | 172 (129-244) | +6 (3.6) | 0.0530 | 13,059 | 4.2 (2.0-7.4) | +0.0 (0.0) | 0.4148 |
| 2020 | 393,323 | 119 (87-170) | -53 (30.8) | **<0.0001** | 14,148 | 5.0 (3.1-7.5) | +0.8 (19.0) | **0.0023** |
| 2021 | 473,006 | 142 (99-204) | +23 (19.3) | **<0.0001** | 12,016 | 4.1 (2.5-6.8) | -0.9 (18.0) | **0.0009** |

Median different was calculated by comparing median in each year with the preceding year. *p* value was calculated based on Mann-Whitney two-sample test.

The decline in case notification rate was highly heterogeneous across districts (Figure 1 and Supplementary Table 1). The top five districts experiencing the largest case notification rate decline from 2019 to 2020 were Pasaman Barat in West Sumatra province (case notification rate ratio 0·15, 95% CI: 0·13-0·18), Kapuas Hulu in West Kalimantan province (0·16, 95% CI: 0·13-0·20), Yalimo in Papua province (0·19, 95% CI: 0·09-0·44), Sorong Selatan in West Papua province (0·21, 95% CI: 0·29-0·47), and Solok in Maluku province (0·22, 95% CI: 0·48-0·65) (Supplementary Table 1). Compared with the first pandemic year 2020, we observed an improvement in the second pandemic year 2021, i.e., fewer districts maintained a significant decline in TB case notification rate (80% in 2020 to 66% in 2021) (Figure 3A).

**Figure 1:**
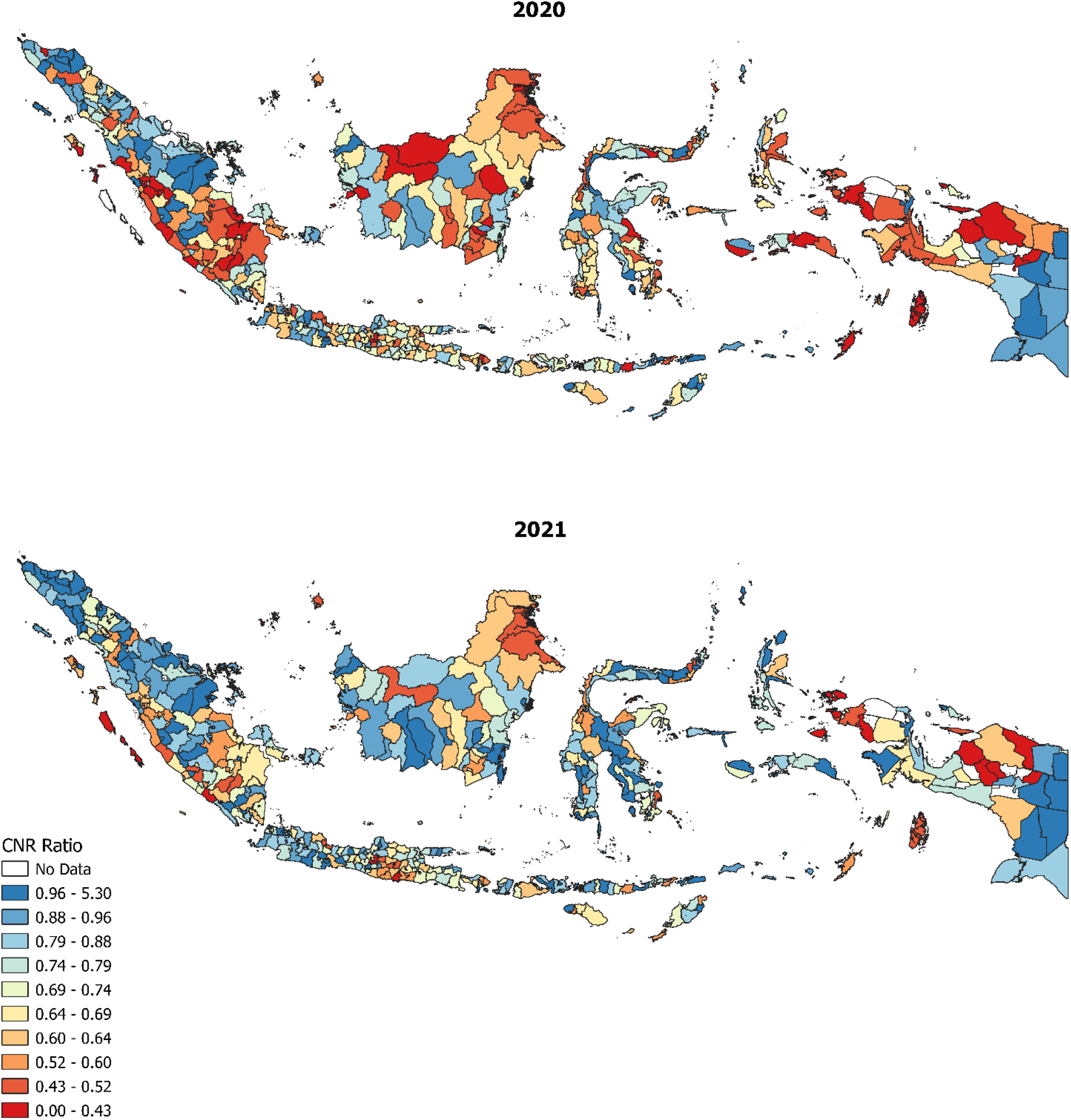
Maps showing case notification rate ratio in 2020 and 2021, compared to 2019 by district. A smaller CNR ratio (i.e., closer to 0) (red) indicates that fewer TB cases have been notified during the pandemic compared to before the pandemic, i.e., greatest negative impact of the COVID-19 pandemic on the national TB programme, whereas a greater CNR ratio (i.e., greater than 1) (blue) indicates that more TB cases have been notified during the pandemic compared to before the pandemic, i.e., smallest negative impact of the COVID-19 pandemic on the national TB programme.

The total number of reported deaths increased from 13,059 in 2019, to 14,148 (8%) in 2020, and decreased to 12,016 (−15%) in 2021. The median number of deaths per quarter sharply increased in quarter 1 of 2020 (start of COVID-19 pandemic), then gradually decreased from quarter 2 of 2020 through quarter 4 of 2021 (Supplementary Figure 2B). The median annual mortality rate sharply increased during the first COVID-19 pandemic year 2020. The median mortality rate was 4·2 (IQR 2·0-7·4) per 100 000 population in 2019, 5·0 (IQR 3·1-7·5) in 2020 and 4·1 (IQR 2·5-6·8) in 2021 (Table 2). Similar to the case notification rate, the significant increase in mortality rate was highly heterogeneous across districts (Figure 2 and Supplementary Table 2). The top five districts experiencing the largest increase in mortality rate from 2019 to 2020 were Barru in South Sulawesi province (mortality rate ratio 16·9, 95% CI: 3·9-73·9), Lebong in Bengkulu province (16·5, 95% CI: 3·7-72·9), Langkat in North Sumatra province (15·9, 95% CI: 6·7-37·7), Kota Manado (15·4, 95% CI: 3·4-69·2), and Kepulauan Talaud in North Sulawesi province (11·7, 95% CI: 2·4-58·3) (Supplementary Table 2). Compared with the first pandemic year 2020, we observed an improvement in the second pandemic year 2021, i.e., fewer districts maintained a significant increase in mortality (20% in 2020 to 13% in 2021) (Figure 3B).

**Figure 2:**
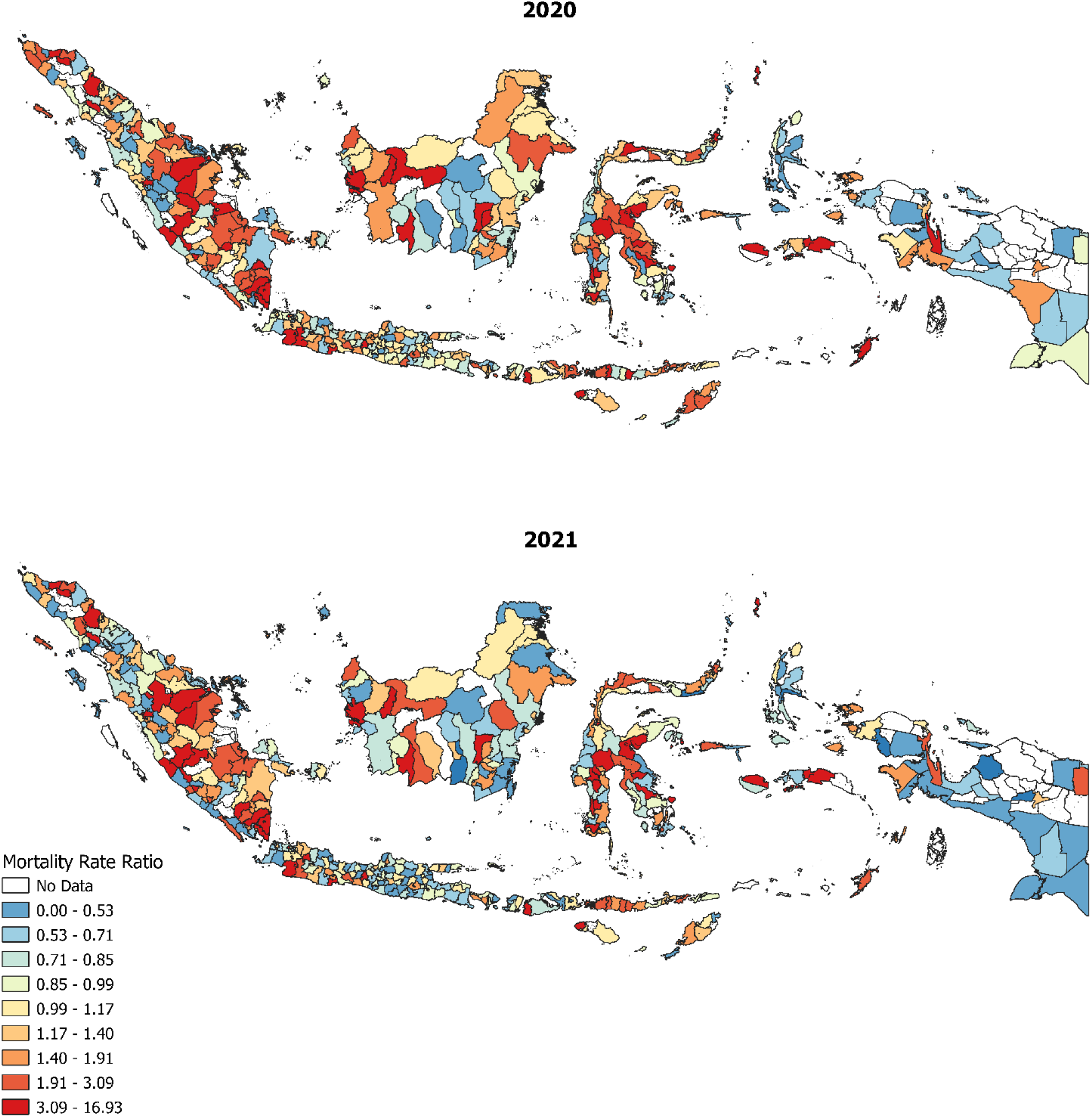
Maps showing mortality rate ratio in 2020 and 2021, compared to 2019 by district. A greater mortality rate ratio (i.e., greater than 1) (red) indicates more reported deaths during the pandemic compared to before the pandemic, i.e., greatest negative impact of the COVID-19 pandemic on the national TB programme, whereas a smaller mortality ratio (i.e., closer to 0) (blue) indicates fewer deaths during the pandemic compared to before the pandemic, i.e., smallest negative impact of the COVID-19 pandemic on the national TB programme.

**Figure 3:**
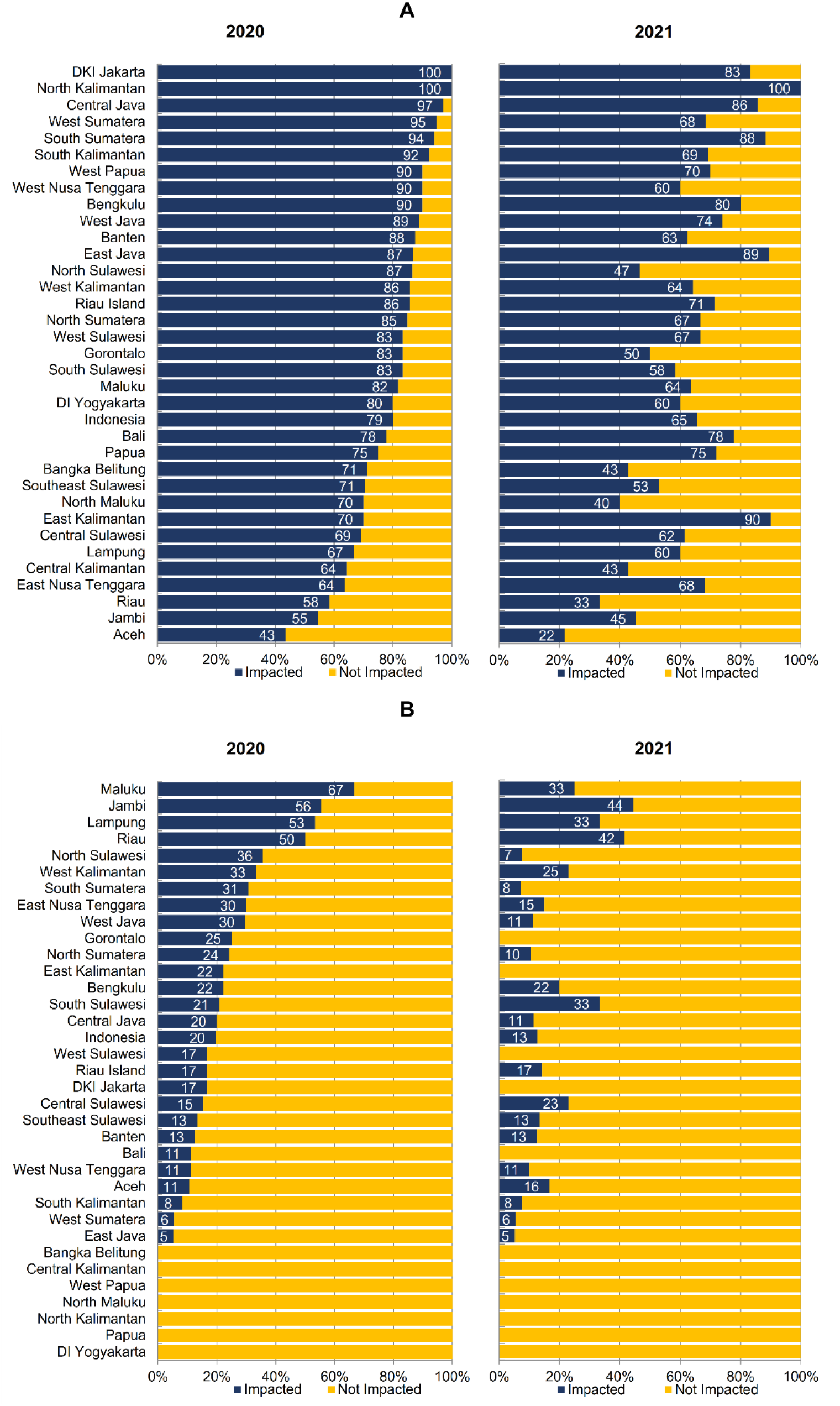
**Proportion of districts within their province impacted based on decrease of case notification rate (A) and increase of mortality rate (B) during the COVID-19 pandemic years 2020 and 2021, compared to the pre-pandemic year 2019.** Districts were categorised as impacted if they have a statistically significant decrease in case notification rate and significant increase in mortality rate in 2020 and 2021, compared to 2019, respectively.

### Treatment coverage of TB cases before (2019) and during the COVID-19 pandemic (2020-2021)

Due to incomplete data, data from only 503 of 514 districts in 2019-2021 were included in this analysis. The overall proportion of TB cases who started treatment decreased from 98% in 2019 to 91% in 2020 (−7%), and 84% (−7%) in 2021 (Table 3). The median proportion of TB cases who started treatment significantly decreased from 100% in 2019 to 95% in 2020, and 88% in 2021. The top five districts experiencing the largest decrease in treatment coverage from 2019 to 2020 were Keerom in Papua province (treatment coverage ratio 0·3, 95% CI: 0·2-0·4), Bombana in Southeast Sulawesi province (0·4, 95% CI: 0·3-0·4), Padang Pariaman in West Sumatra province (0·5, 95% CI: 0·4-0·5), Seluma in Bengkulu province (0·5, 95% CI: 0·3-0·7), and Nias in North Sumatra province (0·5, 95% CI: 0·4-0·6) (Supplementary Table 3). Compared with the first pandemic year 2020, we observed an improvement in the second pandemic year 2021, i.e., more districts had a significant increase in TB treatment coverage (24% in 2020 to 48% in 2021) (Figure 4).

**Table 3.** Tuberculosis treatment coverage in 503 districts in 2019-2021.

| Year | Total new TB cases | Total TB cases who started treatment | TB treatment coverage (%) |  |  |  |
| --- | --- | --- | --- | --- | --- | --- |
|  |  |  | Overall | Median (IQR) | Median difference (% change) | p value |
| 2019 | 564,503 | 554,869 | 98 | 100 (100-100) | 0 | .. |
| 2020 | 392,562 | 355,924 | 91 | 95 (89-100) | -5 (5.0) | **<0.0001** |
| 2021 | 472,065 | 397,117 | 84 | 88 (80-94) | -7 (7.4) | **<0.0001** |

Due to incomplete data, only data from 503 districts in 2019-2021 were included in this analysis. Median different was calculated by comparing median in each year with the preceding year. *p* value was calculated based on Mann-Whitney two-sample test.

**Figure 4:**
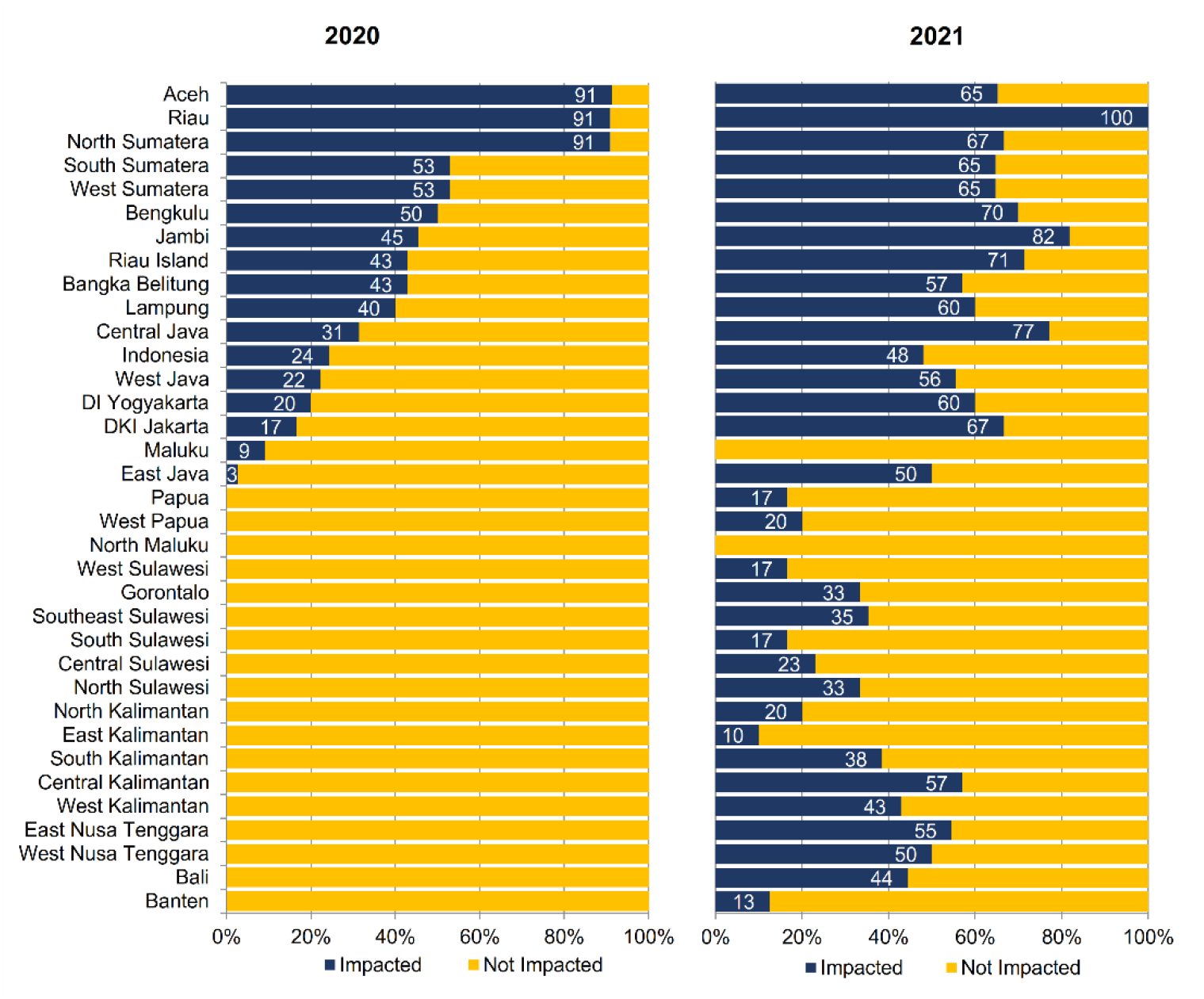
**Proportion of districts within their province impacted based on decrease in treatment coverage during the COVID-19 pandemic years 2020 and 2021, compared to the pre-pandemic year 2019.** Districts were categorised as impacted if they have a statistically significant decrease in treatment coverage in 2020 and 2021, compared to 2019.

### Factors associated with TB case notification rate, treatment coverage, and mortality rate

In the first year of the pandemic (2020), multivariable analysis at the district level (Table 4) found that a significant decline in TB case notification rate was associated with a higher COVID-19 incidence rate and a lower number of facilities with a GeneXpert machine per 100 000 population; a significant decrease in TB treatment coverage was associated with a lower number of doctors per 100 000 population; and a significant increase in mortality rate was associated with a lower number of primary health centres per 100 000 population, a lower per capita domestic expenditure, and a higher level of education.

**Table 4.**
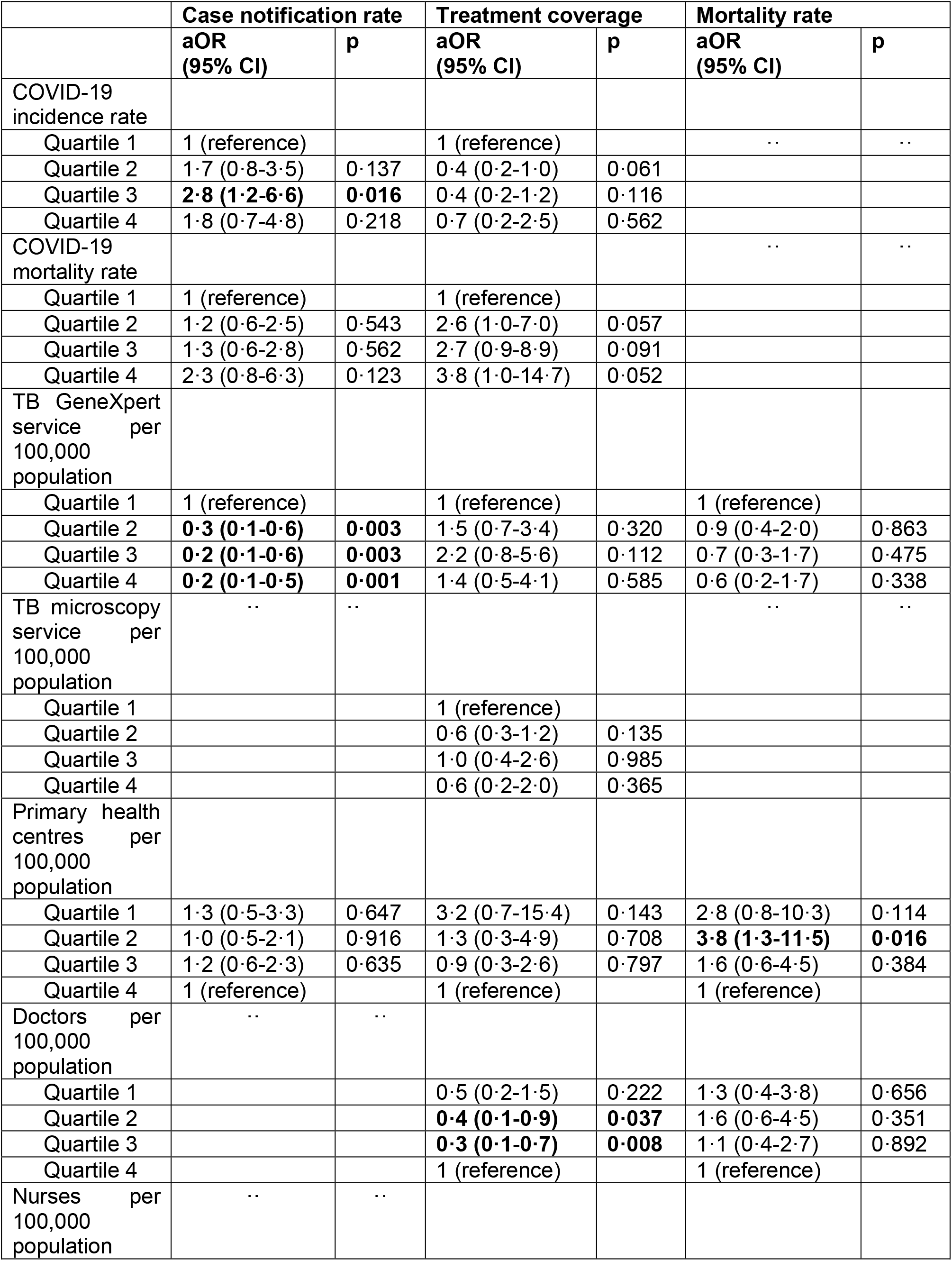
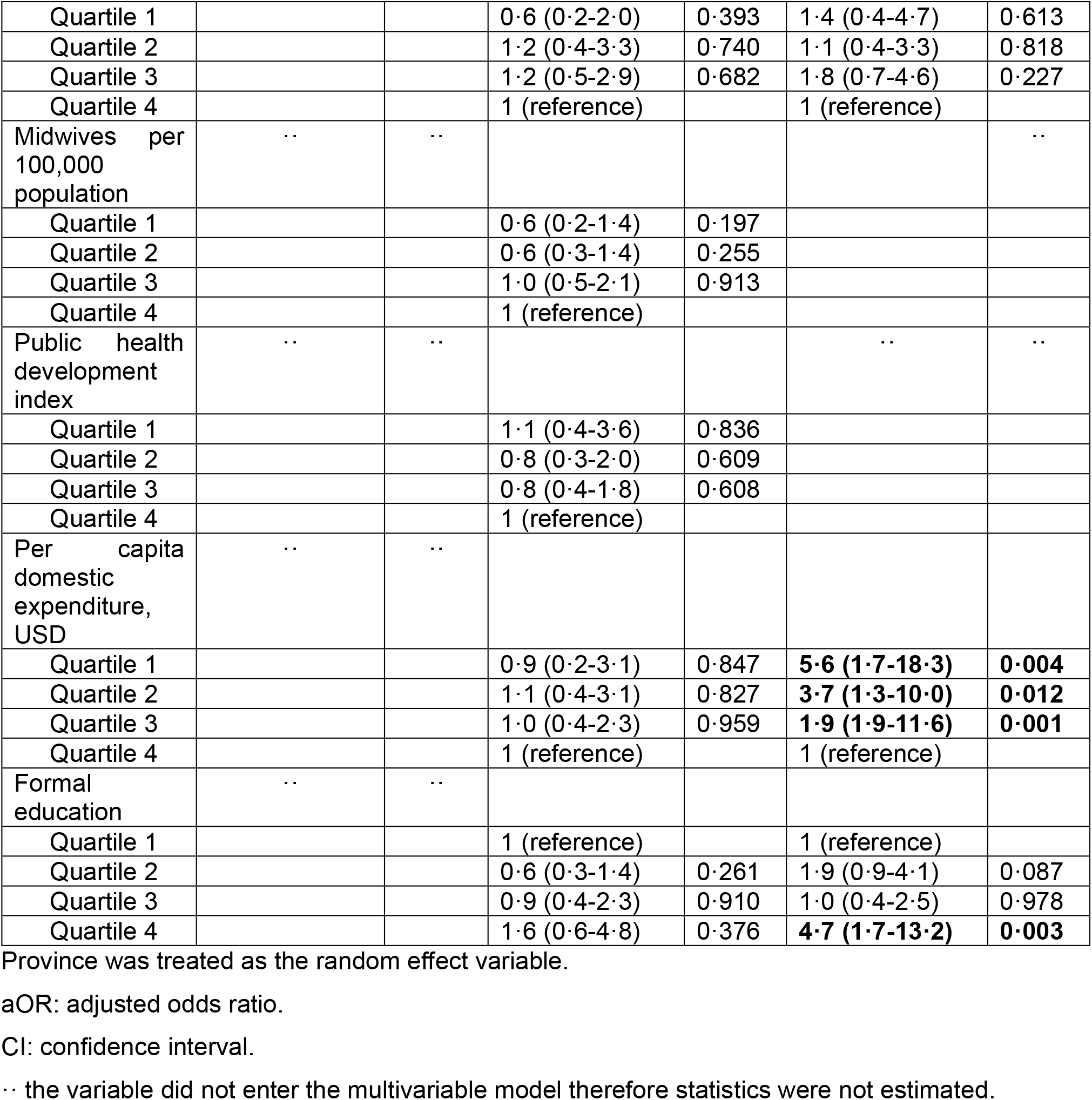
**Multivariable analysis of factors associated with decrease in tuberculosis case notification rate, decrease in treatment coverage, and increase in mortality rate in Indonesia 2020**

|  | Case notification rate |  | Treatment coverage |  | Mortality rate |  |
| --- | --- | --- | --- | --- | --- | --- |
|  | aOR<br>(95% CI) | p | aOR<br>(95% CI) | p | aOR<br>(95% CI) | p |
| COVID-19 incidence rate |  |  |  |  |  |  |
| Quartile 1 | 1 (reference) |  | 1 (reference) |  | .. | .. |
| Quartile 2 | 1.7 (0.8-3.5) | 0.137 | 0.4 (0.2-1.0) | 0.061 |  |  |
| Quartile 3 | **2.8 (1.2-6.6)** | **0.016** | 0.4 (0.2-1.2) | 0.116 |  |  |
| Quartile 4 | 1.8 (0.7-4.8) | 0.218 | 0.7 (0.2-2.5) | 0.562 |  |  |
| COVID-19 mortality rate |  |  |  |  | .. | .. |
| Quartile 1 | 1 (reference) |  | 1 (reference) |  |  |  |
| Quartile 2 | 1.2 (0.6-2.5) | 0.543 | 2.6 (1.0-7.0) | 0.057 |  |  |
| Quartile 3 | 1.3 (0.6-2.8) | 0.562 | 2.7 (0.9-8.9) | 0.091 |  |  |
| Quartile 4 | 2.3 (0.8-6.3) | 0.123 | 3.8 (1.0-14.7) | 0.052 |  |  |
| TB GeneXpert service per 100,000 population |  |  |  |  |  |  |
| Quartile 1 | 1 (reference) |  | 1 (reference) |  | 1 (reference) |  |
| Quartile 2 | **0.3 (0.1-0.6)** | **0.003** | 1.5 (0.7-3.4) | 0.320 | 0.9 (0.4-2.0) | 0.863 |
| Quartile 3 | **0.2 (0.1-0.6)** | **0.003** | 2.2 (0.8-5.6) | 0.112 | 0.7 (0.3-1.7) | 0.475 |
| Quartile 4 | **0.2 (0.1-0.5)** | **0.001** | 1.4 (0.5-4.1) | 0.585 | 0.6 (0.2-1.7) | 0.338 |
| TB microscopy service per 100,000 population | .. | .. |  |  | .. | .. |
| Quartile 1 |  |  | 1 (reference) |  |  |  |
| Quartile 2 |  |  | 0.6 (0.3-1.2) | 0.135 |  |  |
| Quartile 3 |  |  | 1.0 (0.4-2.6) | 0.985 |  |  |
| Quartile 4 |  |  | 0.6 (0.2-2.0) | 0.365 |  |  |
| Primary health centres per 100,000 population |  |  |  |  |  |  |
| Quartile 1 | 1.3 (0.5-3.3) | 0.647 | 3.2 (0.7-15.4) | 0.143 | 2.8 (0.8-10.3) | 0.114 |
| Quartile 2 | 1.0 (0.5-2.1) | 0.916 | 1.3 (0.3-4.9) | 0.708 | **3.8 (1.3-11.5)** | **0.016** |
| Quartile 3 | 1.2 (0.6-2.3) | 0.635 | 0.9 (0.3-2.6) | 0.797 | 1.6 (0.6-4.5) | 0.384 |
| Quartile 4 | 1 (reference) |  | 1 (reference) |  | 1 (reference) |  |
| Doctors per 100,000 population | .. | .. |  |  |  |  |
| Quartile 1 |  |  | 0.5 (0.2-1.5) | 0.222 | 1.3 (0.4-3.8) | 0.656 |
| Quartile 2 |  |  | **0.4 (0.1-0.9)** | **0.037** | 1.6 (0.6-4.5) | 0.351 |
| Quartile 3 |  |  | **0.3 (0.1-0.7)** | **0.008** | 1.1 (0.4-2.7) | 0.892 |
| Quartile 4 |  |  | 1 (reference) |  | 1 (reference) |  |
| Nurses per 100,000 population | .. | .. |  |  |  |  |
| Quartile 1 |  |  | 0.6 (0.2-2.0) | 0.393 | 1.4 (0.4-4.7) | 0.613 |
| Quartile 2 |  |  | 1.2 (0.4-3.3) | 0.740 | 1.1 (0.4-3.3) | 0.818 |
| Quartile 3 |  |  | 1.2 (0.5-2.9) | 0.682 | 1.8 (0.7-4.6) | 0.227 |
| Quartile 4 |  |  | 1 (reference) |  | 1 (reference) |  |
| Midwives per 100,000 population | .. | .. |  |  |  | .. |
| Quartile 1 |  |  | 0.6 (0.2-1.4) | 0.197 |  |  |
| Quartile 2 |  |  | 0.6 (0.3-1.4) | 0.255 |  |  |
| Quartile 3 |  |  | 1.0 (0.5-2.1) | 0.913 |  |  |
| Quartile 4 |  |  | 1 (reference) |  |  |  |
| Public health development index | .. | .. |  |  | .. | .. |
| Quartile 1 |  |  | 1.1 (0.4-3.6) | 0.836 |  |  |
| Quartile 2 |  |  | 0.8 (0.3-2.0) | 0.609 |  |  |
| Quartile 3 |  |  | 0.8 (0.4-1.8) | 0.608 |  |  |
| Quartile 4 |  |  | 1 (reference) |  |  |  |
| Per capita domestic expenditure, USD | .. | .. |  |  |  |  |
| Quartile 1 |  |  | 0.9 (0.2-3.1) | 0.847 | **5.6 (1.7-18.3)** | **0.004** |
| Quartile 2 |  |  | 1.1 (0.4-3.1) | 0.827 | **3.7 (1.3-10.0)** | **0.012** |
| Quartile 3 |  |  | 1.0 (0.4-2.3) | 0.959 | **1.9 (1.9-11.6)** | **0.001** |
| Quartile 4 |  |  | 1 (reference) |  | 1 (reference) |  |
| Formal education | .. | .. |  |  |  |  |
| Quartile 1 |  |  | 1 (reference) |  | 1 (reference) |  |
| Quartile 2 |  |  | 0.6 (0.3-1.4) | 0.261 | 1.9 (0.9-4.1) | 0.087 |
| Quartile 3 |  |  | 0.9 (0.4-2.3) | 0.910 | 1.0 (0.4-2.5) | 0.978 |
| Quartile 4 |  |  | 1.6 (0.6-4.8) | 0.376 | **4.7 (1.7-13.2)** | **0.003** |

Province was treated as the random effect variable.
aOR: adjusted odds ratio.
CI: confidence interval.
.. the variable did not enter the multivariable model therefore statistics were not estimated.

In the second year of the pandemic (2021), multivariable analysis at the district level (Supplementary Table 4) found no factors to be associated with the decrease in TB case notification rate; a significant decrease in TB treatment coverage was associated with a lower number of facilities with a GeneXpert machine and primary health centres per 100 000 population, and a higher level of education; and a significant increase in mortality rate was associated with a lower incidence of COVID-19 and a lower number of primary health centres per 100 000 population.

## Discussion

Our analysis identified a gradual increase in TB case notifications from 2016 through early 2020, corroborating a recently published analysis covering 2017 and 2019.^16^ After the onset of the COVID-19 pandemic in Indonesia in early March 2020, the TB case notification rate decreased by 31% for 2020, signifying one of the most dramatic drops among the high-burden TB countries reported by WHO.^6^ Likewise, there was a significant decrease in TB treatment coverage (7%) and an increase in reported deaths for any reason (8%). Although we did control for COVID-19 mortality in our regression models, the data did not allow us to ascertain cause of death, and estimate mortality attributable to TB, COVID-19, or to any interruptions in vital health services for other health conditions. Nonetheless, these findings affirm substantial impacts of the COVID-19 pandemic on national TB programmes, as reported for several of the TB high-burden, resource-limited countries in sub-Saharan Africa ^7–11^ and Asia.^12,13^ The findings further suggest that the second pandemic year saw signs of recovery in the public health system in many, but not all, districts, with attenuated impacts on TB notifications (80% of districts impacted in 2020 vs 66% in 2021) and/or reported deaths (20% of districts impacted in 2020 vs 13% in 2021), whereas there was continued decline in TB treatment coverage overall (24% of districts impacted in 2020 vs 48% in 2021). Extended reductions in treatment coverage may seriously escalate TB transmission in the coming years.^6^

Even without pandemic disruptions of TB case finding, diagnostic and treatment services, patient pathways to reach a TB diagnosis and access effective treatment are already complex, often involving long delays and visits to multiple health care providers.^5,28,29^ The social and economic shocks of the pandemic deeply exacerbated the fraught path to recovery, correlating with the vulnerability of vital health services delivery. Indeed, we observed substantial heterogeneity of the COVID-19 impacts between districts regarding all three TB study outcomes, and several local health system factors were identified that could offer, at least partially, explanations for the substantial differences observed between districts.

The study findings suggested that districts with a higher COVID-19 incidence and lower resources in the health system were experiencing the most detrimental impacts on TB notification, treatment coverage, and reported death. Specifically, this was true for districts with fewer per capita health facilities with TB GeneXpert diagnostic services, doctors and primary health care centres and per capita domestic expenditure. In Indonesia’s decentralised public health system, resource allocations are mostly under the responsibility of local governments at district level, and as such are highly dependent on local government resources, policy strategies and priorities. Considerable disparities in available financial resources between districts, with per capita domestic expenditure ranging from USD 290-1572, may have contributed to the heterogeneous impact on the overall performance of the TB programme. The pandemic forced local governments to reallocate already scarce health resources to the COVID-19 response ^30^, which, nationally, included an approximately 30% decrease in total TB financing in 2020 compared to 2019. Moreover, a recent review of the Indonesian healthcare system suggested that the existing medical work force was largely insufficient and not evenly distributed to deal with the COVID-19 pandemic.^31^ Therefore, our data highlight the need for both national and sub-national governments to make targeted investments in vital health infrastructure in districts who have shown to be most vulnerable to shocks like COVID-19.

Several additional factors, which our analysis was not able to capture, may have further exacerbated the pandemic impacts on TB case finding and diagnostic and treatment services. First, high rates of infection ^32^ as well as deaths ^33^ among frontline health care workers resulted in substantial absenteeism and task-shifting, especially during the early pandemic. Notably, a recent report estimated that Indonesian healthcare workers had a five times higher chance of death from COVID-19 than the general population.^33,34^ Second, the pandemic affected patient health-seeking behaviour as well as access to essential health services, because of fear of contracting COVID-19 and extra costs for personal protective equipment ^35^, and government-mandated local lockdowns.^36^ Third, emerging data suggest that biological interactions between both pathogens can result in shared dysregulation of immune responses and a dual risk for COVID-19 severity and TB disease progression or poor outcomes.^37,38^ The interplay between these highly prevalent infections warrants further research.

This study had several limitations. This study was based on routine district health office reports, with the potential for gaps in quality control as well as underreporting, which were likely exacerbated by the COVID-19 pandemic. A previous inventory study in 2016 estimated that only half of incident TB cases were detected and reported to the NTCP, with underreporting estimated at 31% for government facilities and 75% for private facilities.^39^ Private health facilities are key entry points for a large proportion of patients seeking health care in Indonesia, including for TB.^5,40^ Additionally, the SITB registers drug-susceptible TB cases, and we could not assess the burden of multidrug/rifampicin-resistant (MDR/RR) cases. Although Indonesia is listed as one of the 30 high-burden countries for drug-resistant TB, incidence estimates at the district-level are lacking and may be underreported, due to limited diagnostic capacities in many remote, under-resourced districts. We were also not able include recent information on district-level HIV and diabetes mellitus prevalence.^20^

In conclusion, Indonesia and other high-burden TB countries, will need to intensify efforts to identify the missed cases with undiagnosed TB, close to the most affected communities, in order to prevent more excess TB reported deaths. Our analysis affirms the notion that people whose access to quality health care services was already limited prior to the pandemic experienced the most acute and severe hardships during the pandemic. Poor resilience of the public healthcare system will, directly and indirectly, exacerbate the shocks and impacts of events like the COVID-19 pandemic. Preparing for the inevitable next pandemic requires fortifying those systems, while prioritising those locations that are the most vulnerable.

## Data Availability

After publication, the datasets used for this study will be made available to others on reasonable requests to the corresponding author and Sub-Directorate of Tuberculosis Ministry of Health of Indonesia, including a detailed research proposal, study objectives and statistical analysis plan.

## Contributors

RLH and IRFE were the principal investigators of this study. HS designed the analysis, did the analysis with assistance from EP, and had full access to all of the data in the study and take responsibility for the integrity of the data and the accuracy of the data analysis. HS, EP, DD, TTP, EL, SS, and SMD contributed to data collection and verification. IRFE and RLH contributed to funding acquisition. HS, IRFE and RLH drafted the paper. All authors critically revised the manuscript for important intellectual content and all authors gave final approval for the version to be published.

## Acknowledgments

We acknowledged the National Tuberculosis Control Program, Ministry of Health of Republic Indonesia and all health care workers involved in the care for the tuberculosis patients, as well as those involved in the field data collection.

## Reference

1 World Health Organization. WHO Coronavirus Disease (COVID-19) Dashboard. https://covid19.who.int. [cited 2023 Feb 10].

2 COVID-19 Excess Mortality Collaborators. Estimating excess mortality due to the COVID-19 pandemic: a systematic analysis of COVID-19-related mortality, 2020-21. Lancet 2022; 399: 1513–36.

3 World Health Organization. Second round of the national pulse survey on continuity of essential health services during the COVID-19 pandemic: January-March 2021. Geneva: World Health Organization; 2021.

4 Downey LE, Gadsden T, Vilas VDR, Peiris D, Jan S. The impact of COVID-19 on essential health service provision for endemic infectious diseases in the South-East Asia region: A systematic review. Lancet Reg Heal - Southeast Asia 2022; 1: 100011.

5 Lestari BW, McAllister S, Hadisoemarto PF, et al. Patient pathways and delays to diagnosis and treatment of tuberculosis in an urban setting in Indonesia. Lancet Reg Heal - West Pacific 2020; 5: 100059.

6 World Health Organization. Global Tuberculosis Report 2021. Geneva: World Health Organization; 2021.

7 Manhiça I, Augusto O, Sherr K, et al. COVID-19-related healthcare impacts: An uncontrolled, segmented time-series analysis of tuberculosis diagnosis services in Mozambique, 2017-2020. BMJ Glob Heal 2022; 7.

8 Lakoh S, Jiba DF, Baldeh M, et al. Impact of covid-19 on tuberculosis case detection and treatment outcomes in Sierra Leone. Trop Med Infect Dis 2021; 6.

9 Lungu PS, Kerkhoff AD, Muyoyeta M, et al. Interrupted time-series analysis of active case-finding for tuberculosis during the COVID-19 pandemic, Zambia. Bull World Health Organ 2022; 100: 205–15.

10 Arega B, Negesso A, Taye B, et al. Impact of COVID-19 pandemic on TB prevention and care in Addis Ababa, Ethiopia: A retrospective database study. BMJ Open 2022; 12: 1–6.

11 G. Caturegli, J. Materi, A. Lombardo, M. Milovanovic, N. Yende, E. Variava, J. E. Golub, N. A. Martinson CJH. Impact of COVID-19 on TB active case finding in Nigeria. Public Heal Action 2020; 10: 157–162.

12 Arentz M, Ma J, Zheng P, Vos T, Murray CJL, Kyu HH. The impact of the COVID-19 pandemic and associated suppression measures on the burden of tuberculosis in India. BMC Infect Dis 2022; 22: 1–8.

13 Fei H, Yinyin X, Hui C, et al. The impact of the COVID-19 epidemic on tuberculosis control in China. Lancet Reg Heal - West Pacific 2020; 3: 100032.

14 World Health Organization. Pulse survey on continuity of essential health services during COVID-19 pandemic: interim report 27 August 2020. Geneva: World Health Organization; 2020.

15 World Health Organization. Global Tuberculosis Report 2022. Geneva: World Health Organization; 2022.

16 Iskandar D, Suwantika AA, Pradipta IS, Postma MJ, Boven JFM Van. Clinical and economic burden of drug-susceptible tuberculosis in Indonesia: national trends 2017 – 19. Lancet Glob Heal 2022; 22: 1–9.

17 Mahendradhata Y, Trisnantoro L, Listyadewi S, et al. Health Systems in Transition Vol. 7 No. 1 2017. The Republic of Indonesia Health System Review. 2017.

18 Agustina R, Dartanto T, Sitompul R, et al. Universal health coverage in Indonesia: concept, progress, and challenges. Lancet 2019; 393: 75–102.

19 Ministry of Health of Indonesia. Public Health Development Index 2018. Jakarta: National Institute of Health Research and Development; 2019.

20 Ledesma JR, Ma J, Vongpradith A, et al. Global, regional, and national sex differences in the global burden of tuberculosis by HIV status, 1990–2019: results from the Global Burden of Disease Study 2019. Lancet Infect Dis 2022; 22: 222–41.

21 Ministry of Health of Indonesia. Sistem Informasi Tuberkulosis. http://www.sitb.id/sitb/app.

22 Ministry of Health of Indonesia. Health Minister Regulation on Tuberculosis Control. Jakarta: Ministry of Health of Indonesia; 2016.

23 Ministry of Health of Indonesia. National guideline for Tuberculosis management. Jakarta: Ministry of Health of Indonesia; 2020.

24 National Bureau of Statistics Indonesia. Data Kependudukan. https://www.bps.go.id/subject/12/kependudukan.html. [Cited 2022 Jan 31].

25 Indonesia COVID-19 National Task Force. National COVID-19 Database. https://data.covid19.go.id/user/login. [Cited 2022 Jan 31].

26 National Bureau of Statistics Indonesia. https://www.bps.go.id/dynamictable/2020/02/17/1771/indeks-pembangunan-manusia-menurut-kabupaten-kota-metode-baru-2010-2019.html. 2019. [Cited 2022 Jan 31]

27 von Elm E, Altman DG, Egger M, Pocock SJ, Gøtzsche PC, Vandenbroucke JP. The Strengthening the Reporting of Observational Studies in Epidemiology (STROBE) statement: guidelines for reporting observational studies. J Clin Epidemiol 2008; 61: 344–9.

28 Kapoor SK, Raman AV, Sachdeva KS, Satyanarayana S. How did the TB patients reach DOTS services in Delhi? a study of patient treatment seeking behavior. PLoS One 2012; 7: 1–6.

29 Murrison LB, Ananthakrishnan R, Sukumar S, et al. How do urban indian private practitioners diagnose and treat tuberculosis? A cross-sectional study in Chennai. PLoS One 2016; 11: 1–14.

30 Caren GJ, Iskandar D, Pitaloka DAE, Abdulah R, Suwantika AA. COVID-19 Pandemic Disruption on the Management of Tuberculosis Treatment in Indonesia. J Multidiscip Healthc 2022; 15: 175–83.

31 Mahendradhata Y, Andayani NLPE, Hasri ET, et al. The Capacity of the Indonesian Healthcare System to Respond to COVID-19. Front Public Heal 2021; 9: 1–9.

32 Sinto R, Utomo D, Suwarti, et al. Antibody Responses and Reactogenicity of a Heterologous, Full-Dose Messenger RNA-1273 Booster in Heavily SARS-CoV-2– Exposed CoronaVac-Vaccinated Health-Care Workers in Indonesia: A Real-World Observational Study. Am J Trop Med Hyg 2022; 1: 115–23.

33 Ekawati LL, Arif A, Hidayana I, et al. Mortality among healthcare workers in Indonesia during 18 months of COVID-19. PLOS Glob Public Heal 2022; 2(12): e0000893.

34 Pramana C, Indriana G, Setyopambudi K. Health Workers and Doctors Death During the Covid-19 Pandemic in Indonesia. Int J Med Rev Case Reports 2021; 5: 1.

35 Klinton JS, Heitkamp P, Rashid A, et al. One year of COVID-19 and its impact on private provider engagement for TB: A rapid assessment of intermediary NGOs in seven high TB burden countries. J Clin Tuberc Other Mycobact Dis 2021; 25: 100277.

36 Djaafara BA, Whittaker C, Watson OJ, et al. Using syndromic measures of mortality to capture the dynamics of COVID-19 in Java, Indonesia, in the context of vaccination rollout. BMC Med 2021; 19: 1–13.

37 Jassat W, Cohen C, Tempia S, et al. Risk factors for COVID-19-related in-hospital mortality in a high HIV and tuberculosis prevalence setting in South Africa: a cohort study. lancet HIV 2021; 8: e554–67.

38 Boulle A, Davies MA, Hussey H, et al. Risk Factors for Coronavirus Disease 2019 (COVID-19) Death in a Population Cohort Study from the Western Cape Province, South Africa. Clin Infect Dis 2021; 73: E2005–15.

39 Kristina L Tobing, Feri Ahmadi, Oster S Suriani, Dian Perwitasari, Dina Bisara Lolong, Jonathan Marbun, Lamria Pangaribuan and RK. Under reported of tuberculosis patients at private health care facilities in Indonesia. Sys Rev Pharm 2021; 12: 899–905.

40 Sunjaya DK, Paskaria C, Herawati DMD, Pramayanti M, Riani R, Parwati I. Initiating a district-based public–private mix to overcome tuberculosis missing cases in Indonesia: readiness to engage. BMC Health Serv Res 2022; 22: 1–11.

