## Supplementary for "Impact of the COVID-19 pandemic on tuberculosis control in Indonesia: a nationwide analysis of programme data and health system vulnerabilities"

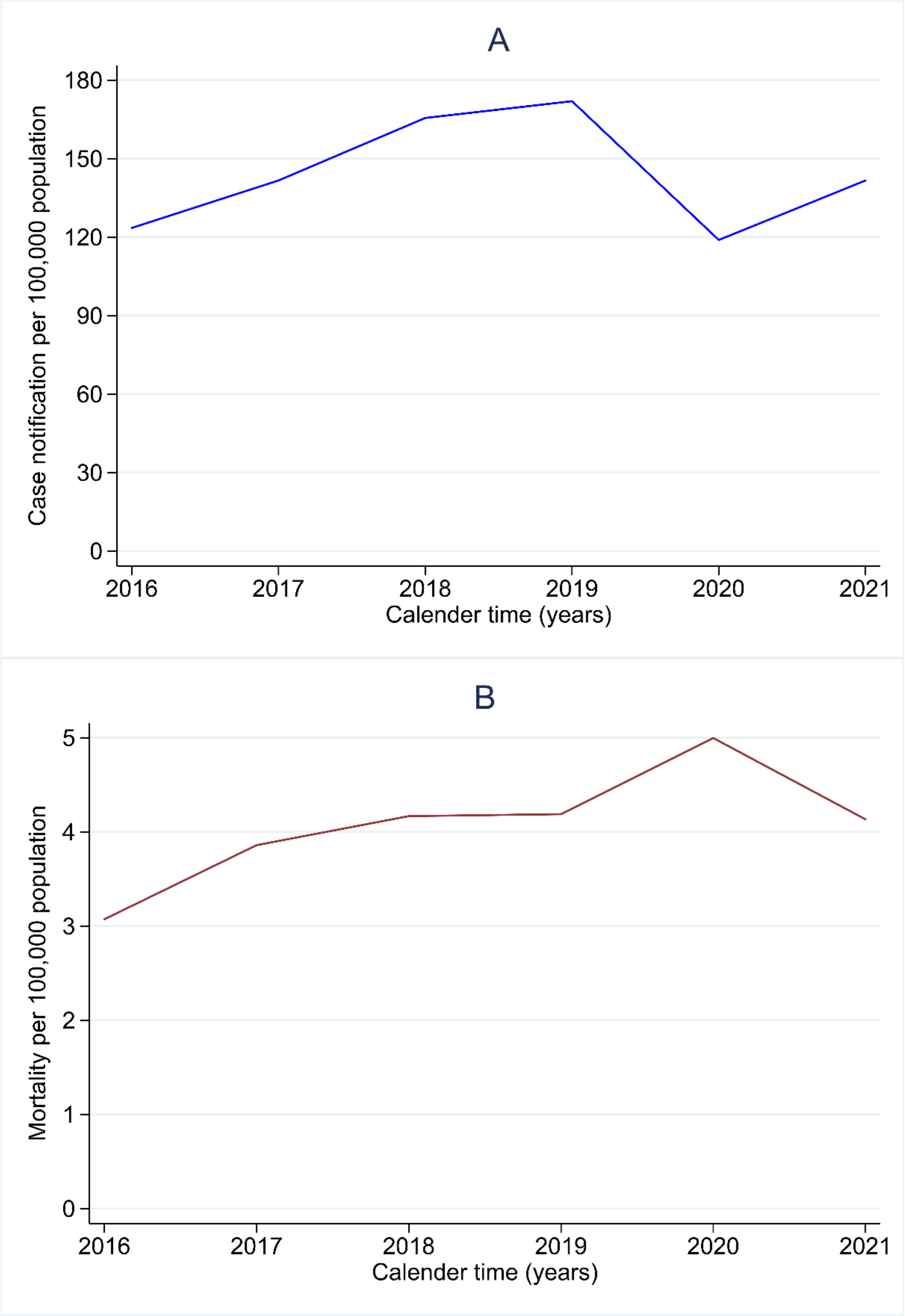

**Supplementary Figure 1: The median national tuberculosis case notification (A) and mortality (B) per 100,000 population over time.**

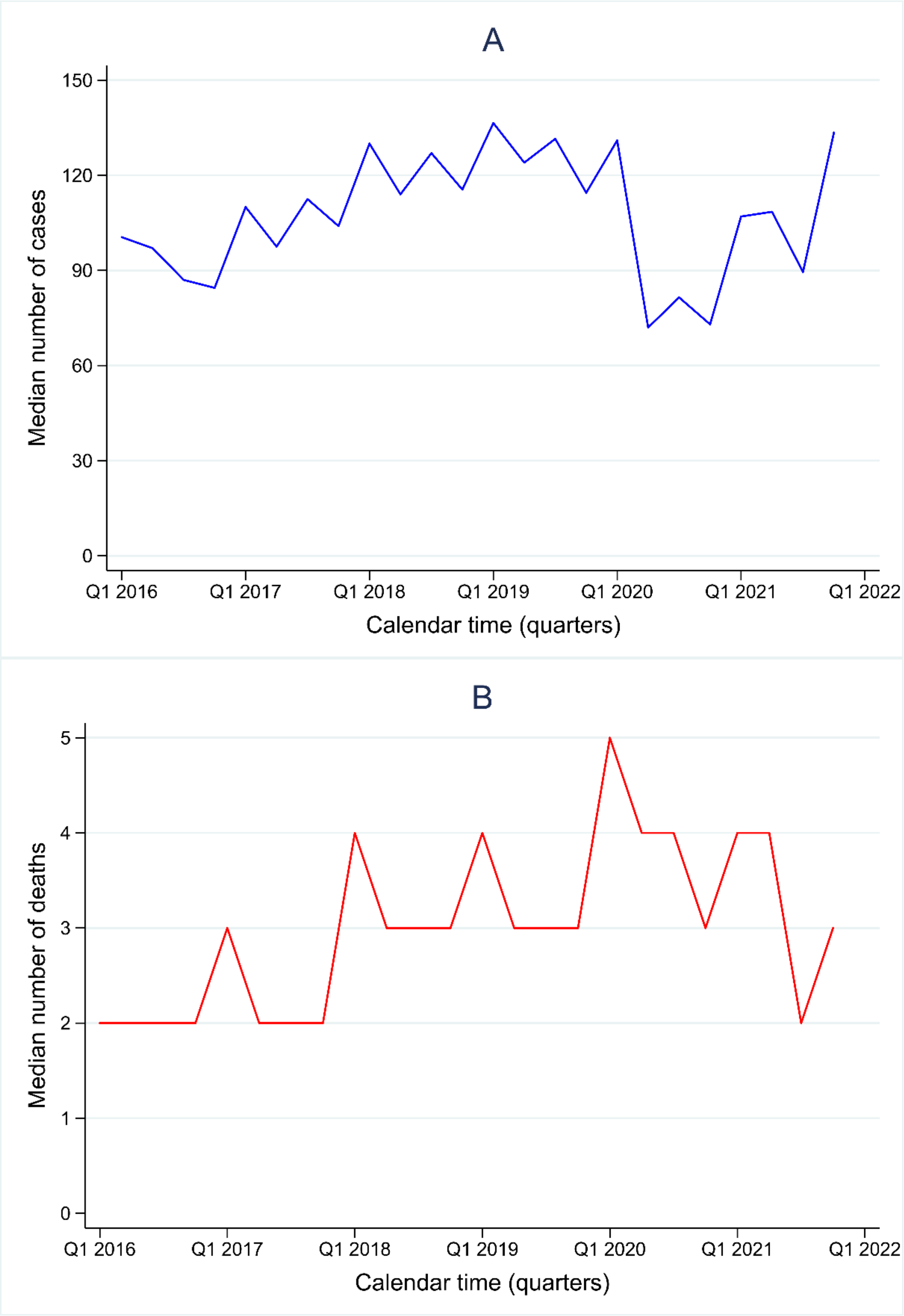

**Supplementary Figure 2: The median national tuberculosis case notification (A) and mortality (B) over time (quarter).**

**Supplementary Table 1. Number of annual populations, reported new TB reported cases, case notification rate (CNR), and CNR ratio in 2020 and 2021 (pandemic), compared to in 2019 (pre-pandemic)**

| District | Number of populations | | | Number of new TB reported cases | | | Case notification rate (CNR) per 100,000 populations | | | CNR ratio for 2020 vs 2019 (95% CI) | CNR ratio for 2021 vs 2019 (95% CI) |
|---|---|---|---|---|---|---|---|---|---|---|---|
| | 2019 | 2020 | 2021 | 2019 | 2020 | 2021 | 2019 | 2020 | 2021 | | |
| **Aceh** | | | | | | | | | | | |
| Simeulue | 93228 | 92865 | 93760 | 124 | 166 | 118 | 133 | 179 | 126 | 1·34 (1·07-1·69) | 0·95 (0·74-1·22) |
| Aceh Singkil | 124101 | 126514 | 128380 | 210 | 145 | 156 | 169 | 115 | 122 | 0·68 (0·55-0 ·84) | 0·72 (0·58-0·88) |
| Aceh Selatan | 238081 | 232414 | 234630 | 375 | 349 | 533 | 158 | 150 | 227 | 0·95 (0·82-1·10) | 1·44 (1·26-1·64) |
| Aceh Tenggara | 216495 | 220860 | 224120 | 130 | 126 | 347 | 60 | 57 | 155 | 0·95 (0· 74-1·21) | 2·58 (2·12-3·13) |
| Aceh Timur | 436081 | 422401 | 427030 | 567 | 613 | 639 | 130 | 145 | 150 | 1·12 (0·99 1·25) | 1·15 (1·03-1·29) |
| Aceh Tengah | 212494 | 215576 | 218680 | 179 | 193 | 232 | 84 | 90 | 106 | 1·06 (0·87-1·30) | 1·26 (1·04-1·53) |
| Aceh Barat | 210113 | 198736 | 200580 | 315 | 288 | 279 | 150 | 145 | 139 | 0·97 (0·82-1·13) | 0·93 (0·79-1·09) |
| Aceh Besar | 425216 | 405535 | 409530 | 404 | 363 | 375 | 95 | 90 | 92 | 0·94 (0·82-1·09) | 0·96 (0·84-1·11) |
| Pidie | 444976 | 435275 | 439400 | 814 | 673 | 773 | 183 | 155 | 176 | 0·85 (0·76-0·94) | 0·96 (0·87-1·06) |
| Bireuen | 471635 | 436418 | 439790 | 859 | 757 | 820 | 182 | 174 | 186 | 0·95 (0·86-1·05) | 1·02 (0·93-1·13) |
| Aceh Utara | 619407 | 602793 | 608110 | 909 | 970 | 763 | 147 | 161 | 126 | 1·10 (1·00-1·20) | 0·86 (0·78-0·94) |
| Aceh Barat Daya | 150393 | 150775 | 152660 | 213 | 286 | 372 | 142 | 190 | 244 | 1·34 (1·12-1·60) | 1·72 (1·46-2·03) |
| Gayo Lues | 94100 | 99532 | 101100 | 198 | 107 | 198 | 210 | 108 | 196 | 0·51 (0·41-0·64) | 0·93 (0·76-1·13) |
| Aceh Tamiang | 295011 | 294356 | 297520 | 426 | 298 | 471 | 144 | 101 | 158 | 0·70 (0·61-0·81) | 1·10 (0·96-1·25) |
| Nagan Raya | 167294 | 168392 | 170590 | 285 | 163 | 249 | 170 | 97 | 146 | 0·57 (0·47-0·69) | 0·86 (0·72-1·02) |
| Aceh Jaya | 92892 | 93159 | 94420 | 213 | 161 | 164 | 229 | 173 | 174 | 0·75 (0·61-0·92) | 0·76 (0·62-0·93) |
| Bener Meriah | 148175 | 161342 | 164520 | 71 | 109 | 95 | 48 | 68 | 58 | 1·41 (1·05-1·90) | 1·21 (0·89-1·64) |
| Pidie Jaya | 161215 | 158397 | 160330 | 199 | 78 | 202 | 123 | 49 | 126 | 0·40 (0·31-0·51) | 1·02 (0·84-1·24) |
| Kota Banda Aceh | 270321 | 252899 | 255030 | 846 | 510 | 786 | 313 | 202 | 308 | 0·64 (0·58-0·72) | 0·99 (0·89-1·09) |
| Kota Sabang | 34874 | 41197 | 42070 | 26 | 29 | 17 | 75 | 70 | 40 | 0·94 (0·56-1·60) | 0·54 (0·30-0·99) |
| Kota Langsa | 176811 | 185971 | 188880 | 412 | 298 | 360 | 233 | 160 | 191 | 0·69 (0·59-0·80) | 0·82 (0·71-0·94) |

| District | Number of populations | | | Number of new TB reported cases | | | Case notification rate (CNR) per 100,000 populations | | | CNR ratio for 2020 vs 2019 (95% CI) | CNR ratio for 2021 vs 2019 (95% CI) |
|---|---|---|---|---|---|---|---|---|---|---|---|
| | 2019 | 2020 | 2021 | 2019 | 2020 | 2021 | 2019 | 2020 | 2021 | | |
| Kota Lhokseumawe | 207202 | 188713 | 189940 | 593 | 317 | 672 | 286 | 168 | 354 | 0·59 (0·51-0·67) | 1·24 (1·11-1·38) |
| Kota Subulussalam | 81417 | 90751 | 92670 | 176 | 227 | 247 | 216 | 250 | 267 | 1·16 (0·95-1·41) | 1·23 (1·02-1·50) |
| **North Sumatera** | | | | | | | | | | | |
| Nias | 143319 | 143983 | 147794 | 300 | 205 | 436 | 209 | 142 | 295 | 0·68 (0·57-0·81) | 1·41 (1·22-1·63) |
| Mandailing Natal | 447287 | 451028 | 478062 | 881 | 854 | 800 | 197 | 189 | 167 | 0·96 (0·87-1·06) | 0·85 (0·77-0·93) |
| Tapanuli Selatan | 281931 | 283389 | 303685 | 656 | 452 | 402 | 233 | 159 | 132 | 0·69 (0·61-0·77) | 0·57 (0·50-0·64) |
| Tapanuli Tengah | 376667 | 382917 | 369300 | 776 | 573 | 574 | 206 | 150 | 155 | 0·73 (0·65-0·81) | 0·75 (0·68-0·84) |
| Tapanuli Utara | 301789 | 303688 | 315222 | 641 | 314 | 449 | 212 | 103 | 142 | 0·49 (0·43-0·56) | 0·67 (0·59-0·76) |
| Toba Samosir | 183712 | 184493 | 208754 | 457 | 261 | 286 | 249 | 141 | 137 | 0·57 (0·49-0·66) | 0·55 (0·48-0·64) |
| Labuhan Batu | 494178 | 501596 | 499982 | 1625 | 845 | 864 | 329 | 168 | 173 | 0·51 (0·47-0·56) | 0·53 (0·48-0·57) |
| Asahan | 729795 | 735026 | 777626 | 987 | 815 | 954 | 135 | 111 | 123 | 0·82 (0·75-0·90) | 0·91 (0·83-0·99) |
| Simalungun | 867922 | 871678 | 1003727 | 1733 | 1566 | 1444 | 200 | 180 | 144 | 0·90 (0·84-0·96) | 0·72 (0·67-0·77) |
| Dairi | 284304 | 285481 | 311665 | 577 | 485 | 482 | 203 | 170 | 155 | 0·84 (0·74-0·94) | 0·76 (0·68-0·86) |
| Karo | 415878 | 421997 | 409077 | 824 | 597 | 577 | 198 | 141 | 141 | 0·71 (0·64-0·79) | 0·71 (0·64-0·79) |
| Deli Serdang | 2195709 | 2234320 | 1941374 | 3415 | 2794 | 3448 | 156 | 125 | 178 | 0·80 (0·76-0·85) | 1·14 (1·09-1·20) |
| Langkat | 1041775 | 1048100 | 1034519 | 1833 | 1161 | 1297 | 176 | 111 | 125 | 0·63 (0·59-0·68) | 0·71 (0·66-0·76) |
| Nias Selatan | 319902 | 322520 | 366163 | 174 | 76 | 121 | 54 | 24 | 33 | 0·43 (0·33-0·56) | 0·61 (0·48-0·76) |
| Humbang Hasundutan | 190186 | 191776 | 199719 | 257 | 203 | 284 | 135 | 106 | 142 | 0·78 (0·65-0·94) | 1·05 (0·89-1·25) |
| Pakpak Bharat | 48935 | 49688 | 53315 | 125 | 95 | 90 | 255 | 191 | 169 | 0·75 (0·57-0·98) | 0·66 (0·50-0·86) |
| Samosir | 126188 | 126710 | 137696 | 247 | 172 | 180 | 196 | 136 | 131 | 0·69 (0·57-0·84) | 0·67 (0·55-0·81) |
| Serdang Bedagai | 616396 | 617772 | 662076 | 946 | 618 | 743 | 153 | 100 | 112 | 0·65 (0·59-0·72) | 0·73 (0·66-0·80) |
| Batu Bara | 416493 | 420103 | 413171 | 461 | 469 | 527 | 111 | 112 | 128 | 1·01 (0·89-1·15) | 1·15 (1·02-1·31) |
| Padang Lawas Utara | 272713 | 277423 | 263551 | 459 | 299 | 428 | 168 | 108 | 162 | 0·64 (0·55-0·74) | 0·97 (0·85-1·10) |
| Padang Lawas | 281239 | 286627 | 263719 | 377 | 347 | 568 | 134 | 121 | 215 | 0·90 (0·78-1·04) | 1·61 (1·41-1·83) |

| District | Number of populations | | | Number of new TB reported cases | | | Case notification rate (CNR) per 100,000 populations | | | CNR ratio for 2020 vs 2019 (95% CI) | CNR ratio for 2021 vs 2019 (95% CI) |
|---|---|---|---|---|---|---|---|---|---|---|---|
| | 2019 | 2020 | 2021 | 2019 | 2020 | 2021 | 2019 | 2020 | 2021 | | |
| Labuhan Batu Selatan | 338982 | 344819 | 316798 | 455 | 368 | 365 | 134 | 107 | 115 | 0·80 (0·69-0·91) | 0·86 (0·75-0·98) |
| Labuhan Batu Utara | 363816 | 366603 | 385869 | 583 | 518 | 595 | 160 | 141 | 154 | 0·88 (0·78-0·99) | 0·96 (0·86-1·08) |
| Nias Utara | 137967 | 138800 | 148790 | 155 | 94 | 91 | 112 | 68 | 61 | 0·60 (0·47-0·78) | 0·54 (0·42-0·70) |
| Nias Barat | 82154 | 82425 | 90585 | 94 | 49 | 54 | 114 | 59 | 60 | 0·52 (0·37-0·73) | 0·52 (0·38-0·72) |
| Kota Sibolga | 87626 | 87791 | 89932 | 327 | 186 | 440 | 373 | 212 | 489 | 0·57 (0·48-0·68) | 1·31 (1·14-1·51) |
| Kota Tanjung Balai | 175223 | 177005 | 177640 | 798 | 617 | 770 | 455 | 349 | 433 | 0·77 (0·69-0·85) | 0·95 (0·86-1·05) |
| Kota Pematang Siantar | 255317 | 257110 | 270768 | 801 | 483 | 693 | 314 | 188 | 256 | 0·60 (0·54-0·67) | 0·82 (0·74-0·90) |
| Kota Tebing Tinggi | 164402 | 166100 | 174969 | 236 | 226 | 341 | 144 | 136 | 195 | 0·95 (0·79-1·14) | 1·36 (1·15-1·60) |
| Kota Medan | 2279894 | 2295003 | 2460858 | 8186 | 5455 | 7329 | 359 | 238 | 298 | 0·66 (0·64-0·68) | 0·83 (0·80-0·86) |
| Kota Binjai | 276597 | 279302 | 295361 | 477 | 486 | 691 | 172 | 174 | 234 | 1·01 (0·89-1·14) | 1·36 (1·21-1·52) |
| Kota Padangsidimpuan | 221827 | 224483 | 227674 | 572 | 347 | 454 | 258 | 155 | 199 | 0·60 (0·53-0·68) | 0·77 (0·68-0·87) |
| Kota Gunungsitoli | 142426 | 143776 | 136707 | 217 | 129 | 84 | 152 | 90 | 61 | 0·59 (0·47-0·73) | 0·40 (0·32-0·51) |
| **West Sumatera** | | | | | | | | | | | |
| Kepulauan Mentawai | 92021 | 87623 | 88389 | 202 | 1 | 62 | 220 | 1 | 70 | 0·01 (0·00-0·01) | 0·32 (0·24-0·42) |
| Pesisir Selatan | 463923 | 504418 | 509618 | 1204 | 634 | 705 | 260 | 126 | 138 | 0·48 (0·44-0·53) | 0·53 (0·49-0·58) |
| Solok | 373414 | 391497 | 394237 | 415 | 95 | 244 | 111 | 24 | 62 | 0·22 (0·18-0·27) | 0·56 (0·48-0·65) |
| Sijunjung | 237376 | 235045 | 237313 | 331 | 114 | 218 | 139 | 49 | 92 | 0·35 (0·28-0·43) | 0·66 (0·56-0·78) |
| Tanah Datar | 348219 | 371704 | 373693 | 461 | 188 | 300 | 132 | 51 | 80 | 0·38 (0·32-0·45) | 0·61 (0·53-0·70) |
| Padang Pariaman | 415613 | 430626 | 433018 | 884 | 1311 | 1815 | 213 | 304 | 419 | 1·43 (1·31-1·56) | 1·97 (1·82-2·13) |
| Agam | 491282 | 529138 | 534202 | 757 | 503 | 503 | 154 | 95 | 94 | 0·62 (0·55-0·69) | 0·61 (0·55-0·68) |
| Lima Puluh Kota | 382817 | 383525 | 385634 | 513 | 308 | 333 | 134 | 80 | 86 | 0·60 (0·52-0·69) | 0·64 (0·56-0·74) |
| Pasaman | 281211 | 299851 | 303103 | 550 | 338 | 527 | 196 | 113 | 174 | 0·58 (0·50-0·66) | 0·89 (0·79-1·00) |
| Solok Selatan | 171075 | 182027 | 184854 | 279 | 196 | 257 | 163 | 108 | 139 | 0·66 (0·55-0·79) | 0·85 (0·72-1·01) |
| Dharmasraya | 247579 | 228591 | 231217 | 411 | 135 | 214 | 166 | 59 | 93 | 0·36 (0·30-0·43) | 0·56 (0·47-0·66) |

| District | Number of populations | | | Number of new TB reported cases | | | Case notification rate (CNR) per 100,000 populations | | | CNR ratio for 2020 vs 2019 (95% CI) | CNR ratio for 2021 vs 2019 (95% CI) |
|---|---|---|---|---|---|---|---|---|---|---|---|
| | 2019 | 2020 | 2021 | 2019 | 2020 | 2021 | 2019 | 2020 | 2021 | | |
| Pasaman Barat | 443722 | 431672 | 436313 | 815 | 122 | 653 | 184 | 28 | 150 | 0·15 (0·13-0·18) | 0·82 (0·74-0·90) |
| Kota Padang | 950871 | 909040 | 913448 | 3052 | 1644 | 2782 | 321 | 181 | 305 | 0·56 (0·53-0·60) | 0·95 (0·90-1·00) |
| Kota Solok | 71010 | 73438 | 74469 | 269 | 188 | 237 | 379 | 256 | 318 | 0·68 (0·56-0·81) | 0·84 (0·71-1·00) |
| Kota Sawah Lunto | 62524 | 65138 | 65687 | 101 | 8 | 68 | 162 | 12 | 104 | 0·08 (0·04-0·13) | 0·64 (0·47-0·87) |
| Kota Padang Panjang | 53693 | 56311 | 56971 | 176 | 98 | 190 | 328 | 174 | 334 | 0·53 (0·42-0·68) | 1·02 (0·83-1·25) |
| Kota Bukittinggi | 130773 | 121028 | 121588 | 487 | 316 | 432 | 372 | 261 | 355 | 0·70 (0·61-0·81) | 0·95 (0·84-1·09) |
| Kota Payakumbuh | 135573 | 139576 | 141184 | 266 | 183 | 227 | 196 | 131 | 161 | 0·67 (0·55-0·81) | 0·82 (0·69-0·98) |
| Kota Pariaman | 88501 | 94224 | 95294 | 289 | 93 | 238 | 327 | 99 | 250 | 0·30 (0·24-0·38) | 0·77 (0·64-0·91) |
| **Riau** | | | | | | | | | | | |
| Kuantan Singingi | 327316 | 334943 | 339894 | 348 | 272 | 303 | 106 | 81 | 89 | 0·76 (0·65-0·89) | 0·84 (0·72-0·98) |
| Indragiri Hulu | 441789 | 444548 | 453241 | 361 | 423 | 488 | 82 | 95 | 108 | 1·16 (1·01-1·34) | 1·32 (1·15-1·51) |
| Indragiri Hilir | 740598 | 654909 | 658025 | 475 | 591 | 629 | 64 | 90 | 96 | 1·41 (1·25-1·59) | 1·49 (1·32-1·68) |
| Pelalawan | 483622 | 390046 | 399264 | 656 | 576 | 486 | 136 | 148 | 122 | 1·09 (0·97-1·22) | 0·90 (0·80-1·01) |
| Siak | 489996 | 457940 | 466683 | 480 | 370 | 349 | 98 | 81 | 75 | 0·83 (0·72-0·94) | 0·76 (0·67-0·88) |
| Kampar | 871117 | 841332 | 857752 | 985 | 841 | 856 | 113 | 100 | 100 | 0·88 (0·81-0·97) | 0·88 (0·81-0·97) |
| Rokan Hulu | 692120 | 561385 | 570952 | 1025 | 848 | 795 | 148 | 151 | 139 | 1·02 (0·93-1·12) | 0·94 (0·86-1·03) |
| Bengkalis | 573003 | 565569 | 573504 | 809 | 739 | 805 | 141 | 131 | 140 | 0·93 (0·84-1·02) | 0·99 (0·90-1·10) |
| Rokan Hilir | 714497 | 637161 | 646791 | 1302 | 1018 | 1064 | 182 | 160 | 165 | 0·88 (0·81-0·95) | 0·90 (0·83-0·98) |
| Kepulauan Meranti | 185516 | 206116 | 209460 | 201 | 171 | 214 | 108 | 83 | 102 | 0·77 (0·62-0·94) | 0·94 (0·78-1·14) |
| Kota Pekanbaru | 1143359 | 983356 | 994585 | 3653 | 2825 | 3378 | 319 | 287 | 340 | 0·90 (0·86-0·94) | 1·06 (1·01-1·11) |
| Kota Dumai | 308812 | 316782 | 323452 | 704 | 627 | 685 | 228 | 198 | 212 | 0·87 (0·78-0·97) | 0·93 (0·84-1·03) |
| **Jambi** | | | | | | | | | | | |
| Kerinci | 238682 | 250300 | 251900 | 135 | 206 | 151 | 57 | 82 | 60 | 1·46 (1·17-1·81) | 1·06 (0·84-1·34) |
| Merangin | 388928 | 354100 | 355700 | 737 | 381 | 550 | 189 | 108 | 155 | 0·57 (0·50-0·64) | 0·82 (0·73-0·91) |

| District | Number of populations | | | Number of new TB reported cases | | | Case notification rate (CNR) per 100,000 populations | | | CNR ratio for 2020 vs 2019 (95% CI) | CNR ratio for 2021 vs 2019 (95% CI) |
|---|---|---|---|---|---|---|---|---|---|---|---|
| | 2019 | 2020 | 2021 | 2019 | 2020 | 2021 | 2019 | 2020 | 2021 | | |
| Sarolangun | 301908 | 290100 | 293600 | 527 | 461 | 463 | 175 | 159 | 158 | 0·91 (0·80-1·03) | 0·90 (0·80-1·02) |
| Batang Hari | 272879 | 301700 | 306700 | 418 | 257 | 381 | 153 | 85 | 124 | 0·56 (0·48-0·65) | 0·81 (0·71-0·93) |
| Muaro Jambi | 443364 | 402000 | 406700 | 455 | 198 | 375 | 103 | 49 | 92 | 0·48 (0·41-0·57) | 0·90 (0·78-1·03) |
| Tanjung Jabung Timur | 219985 | 229800 | 231800 | 228 | 166 | 141 | 104 | 72 | 61 | 0·70 (0·57-0·85) | 0·59 (0·48-0·72) |
| Tanjung Jabung Barat | 333932 | 317500 | 320600 | 545 | 290 | 319 | 163 | 91 | 100 | 0·56 (0·49-0·64) | 0·61 (0·53-0·70) |
| Tebo | 354485 | 337700 | 340900 | 377 | 240 | 263 | 106 | 71 | 77 | 0·67 (0·57-0·78) | 0·73 (0·62-0·85) |
| Bungo | 374770 | 362400 | 367200 | 291 | 357 | 455 | 78 | 99 | 124 | 1·27 (1·09-1·48) | 1·60 (1·38-1·85) |
| Kota Jambi | 604736 | 606200 | 612200 | 1055 | 980 | 1229 | 174 | 162 | 201 | 0·93 (0·85-1·01) | 1·15 (1·06-1·25) |
| Kota Sungai Penuh | 90910 | 96600 | 97800 | 52 | 49 | 72 | 57 | 51 | 74 | 0·89 (0·60-1·31) | 1·29 (0·90-1·84) |
| **South Sumatera** | | | | | | | | | | | |
| Ogan Komering Ulu | 368756 | 367603 | 371106 | 1150 | 398 | 562 | 312 | 108 | 151 | 0·35 (0·31-0·39) | 0·49 (0·44-0·54) |
| Ogan Komering Ilir | 832151 | 769348 | 772742 | 1711 | 688 | 1068 | 206 | 89 | 138 | 0·44 (0·40-0·47) | 0·67 (0·62-0·73) |
| Muara Enim | 637556 | 612900 | 617846 | 1732 | 1041 | 1155 | 272 | 170 | 187 | 0·63 (0·58-0·67) | 0·69 (0·64-0·74) |
| Lahat | 409348 | 430071 | 434939 | 712 | 387 | 586 | 174 | 90 | 135 | 0·52 (0·46-0·58) | 0·78 (0·69-0·86) |
| Musi Rawas | 405175 | 395570 | 398732 | 775 | 515 | 624 | 191 | 130 | 156 | 0·68 (0·61-0·76) | 0·82 (0·74-0·91) |
| Musi Banyuasin | 649085 | 622206 | 627070 | 1588 | 774 | 912 | 245 | 124 | 145 | 0·51 (0·47-0·55) | 0·59 (0·55-0·64) |
| Banyu Asin | 857097 | 836914 | 843871 | 2311 | 866 | 1538 | 270 | 103 | 182 | 0·38 (0·36-0·41) | 0·68 (0·63-0·72) |
| Ogan Komering Ulu Selatan | 363004 | 408981 | 416616 | 365 | 186 | 263 | 101 | 45 | 63 | 0·45 (0·38-0·54) | 0·63 (0·54-0·73) |
| Ogan Komering Ulu Timur | 677080 | 649853 | 653062 | 1287 | 497 | 627 | 190 | 76 | 96 | 0·40 (0·36-0·44) | 0·51 (0·46-0·55) |
| Ogan Ilir | 429595 | 416549 | 419401 | 1035 | 499 | 649 | 241 | 120 | 155 | 0·50 (0·45-0·55) | 0·64 (0·58-0·71) |
| Empat Lawang | 250465 | 333622 | 343839 | 398 | 293 | 282 | 159 | 88 | 82 | 0·55 (0·48-0·64) | 0·52 (0·44-0·60) |
| Penukal Abab | 190062 | 194900 | 197290 | 410 | 282 | 252 | 216 | 145 | 128 | 0·67 (0·58-0·78) | 0·59 (0·51-0·69) |
| Musi Rawas Utara | 192540 | 188861 | 190420 | 277 | 274 | 355 | 144 | 145 | 186 | 1·01 (0·85-1·19) | 1·30 (1·11-1·52) |
| Kota Palembang | 1674243 | 1668848 | 1686073 | 4589 | 3154 | 5730 | 274 | 189 | 340 | 0·69 (0·66-0·72) | 1·24 (1·19-1·29) |

| District | Number of populations | | | Number of new TB reported cases | | | Case notification rate (CNR) per 100,000 populations | | | CNR ratio for 2020 vs 2019 (95% CI) | CNR ratio for 2021 vs 2019 (95% CI) |
|---|---|---|---|---|---|---|---|---|---|---|---|
| | 2019 | 2020 | 2021 | 2019 | 2020 | 2021 | 2019 | 2020 | 2021 | | |
| Kota Prabumulih | 188669 | 193196 | 195748 | 1057 | 412 | 584 | 560 | 213 | 298 | 0·38 (0·34-0·42) | 0·53 (0·48-0·59) |
| Kota Pagar Alam | 139192 | 143844 | 145266 | 256 | 140 | 157 | 184 | 97 | 108 | 0·53 (0·43-0·65) | 0·59 (0·48-0·72) |
| Kota Lubuklinggau | 233178 | 234166 | 236828 | 975 | 292 | 643 | 418 | 125 | 272 | 0·30 (0·26-0·34) | 0·65 (0·59-0·72) |
| **Bengkulu** | | | | | | | | | | | |
| Bengkulu Selatan | 158410 | 166250 | 167989 | 207 | 163 | 151 | 131 | 98 | 90 | 0·75 (0·61-0·92) | 0·69 (0·56-0·85) |
| Rejang Lebong | 260900 | 276640 | 278793 | 411 | 213 | 191 | 158 | 77 | 69 | 0·49 (0·42-0·57) | 0·44 (0·37-0·51) |
| Bengkulu Utara | 310000 | 296520 | 299395 | 271 | 128 | 179 | 87 | 43 | 60 | 0·49 (0·40-0·61) | 0·68 (0·57-0·83) |
| Kaur | 121210 | 126550 | 127953 | 121 | 63 | 36 | 100 | 50 | 28 | 0·50 (0·37-0·67) | 0·28 (0·20-0·40) |
| Seluma | 193800 | 207880 | 210505 | 114 | 47 | 85 | 59 | 23 | 40 | 0·38 (0·28-0·53) | 0·69 (0·52-0·91) |
| Mukomuko | 193880 | 190500 | 193196 | 542 | 154 | 281 | 280 | 81 | 145 | 0·29 (0·24-0·34) | 0·52 (0·45-0·60) |
| Lebong | 116610 | 106290 | 106767 | 176 | 158 | 156 | 151 | 149 | 146 | 0·99 (0·79-1·22) | 0·97 (0·78-1·20) |
| Kepahiang | 137190 | 149740 | 151640 | 310 | 269 | 436 | 226 | 180 | 288 | 0·80 (0·68-0·94) | 1·27 (1·10-1·47) |
| Bengkulu Tengah | 114700 | 116710 | 118100 | 134 | 71 | 88 | 117 | 61 | 75 | 0·52 (0·39-0·69) | 0·64 (0·49-0·83) |
| Kota Bengkulu | 385140 | 373590 | 378604 | 986 | 559 | 757 | 256 | 150 | 200 | 0·58 (0·53-0·65) | 0·78 (0·71-0·86) |
| **Lampung** | | | | | | | | | | | |
| Lampung Barat | 302828 | 304874 | 302749 | 278 | 204 | 273 | 92 | 67 | 90 | 0·73 (0·61-0·87) | 0·98 (0·83-1·16) |
| Tanggamus | 598299 | 603706 | 645807 | 847 | 788 | 627 | 142 | 131 | 97 | 0·92 (0·84-1·02) | 0·69 (0·62-0·76) |
| Lampung Selatan | 1011286 | 1019789 | 1071727 | 2268 | 1203 | 1454 | 224 | 118 | 136 | 0·53 (0·49-0·56) | 0·61 (0·57-0·65) |
| Lampung Timur | 1044320 | 1051994 | 1118115 | 1367 | 937 | 1024 | 131 | 89 | 92 | 0·68 (0·63-0·74) | 0·70 (0·65-0·76) |
| Lampung Tengah | 1281310 | 1290407 | 1477395 | 2761 | 1999 | 1992 | 215 | 155 | 135 | 0·72 (0·68-0·76) | 0·63 (0·59-0·66) |
| Lampung Utara | 616897 | 618818 | 634117 | 982 | 626 | 1088 | 159 | 101 | 172 | 0·64 (0·58-0·70) | 1·08 (0·99-1·17) |
| Way Kanan | 450109 | 453921 | 476871 | 728 | 578 | 568 | 162 | 127 | 119 | 0·79 (0·71-0·88) | 0·74 (0·66-0·82) |
| Tulangbawang | 450902 | 455891 | 430630 | 735 | 590 | 620 | 163 | 129 | 144 | 0·79 (0·71-0·88) | 0·88 (0·79-0·98) |
| Pesawaran | 444380 | 448410 | 481708 | 536 | 416 | 467 | 121 | 93 | 97 | 0·77 (0·68-0·87) | 0·80 (0·71-0·91) |

| District | Number of populations | | | Number of new TB reported cases | | | Case notification rate (CNR) per 100,000 populations | | | CNR ratio for 2020 vs 2019 (95% CI) | CNR ratio for 2021 vs 2019 (95% CI) |
|---|---|---|---|---|---|---|---|---|---|---|---|
| | 2019 | 2020 | 2021 | 2019 | 2020 | 2021 | 2019 | 2020 | 2021 | | |
| Pringsewu | 400187 | 403115 | 406823 | 650 | 691 | 767 | 162 | 171 | 189 | 1·06 (0·95-1·17) | 1·16 (1·05-1·29) |
| Mesuji | 200198 | 200999 | 229772 | 302 | 266 | 237 | 151 | 132 | 103 | 0·88 (0·74-1·03) | 0·68 (0·58-0·81) |
| Tulang Bawang Barat | 273215 | 274905 | 287707 | 416 | 398 | 391 | 152 | 145 | 136 | 0·95 (0·83-1·09) | 0·89 (0·78-1·02) |
| Pesisir Barat | 154895 | 155883 | 163641 | 161 | 126 | 138 | 104 | 81 | 84 | 0·78 (0·62-0·98) | 0·81 (0·65-1·02) |
| Kota Bandar Lampung | 1051500 | 1068982 | 1184949 | 3571 | 2649 | 3066 | 340 | 248 | 259 | 0·73 (0·69-0·77) | 0·76 (0·73-0·80) |
| Kota Metro | 167411 | 169507 | 169781 | 520 | 499 | 528 | 311 | 294 | 311 | 0·95 (0·84-1·07) | 1·00 (0·89-1·13) |
| **Bangka Belitung** | | | | | | | | | | | |
| Bangka | 337337 | 343821 | 337286 | 515 | 401 | 420 | 153 | 117 | 125 | 0·76 (0·67-0·87) | 0·82 (0·72-0·93) |
| Belitung | 189824 | 193493 | 189752 | 281 | 238 | 239 | 148 | 123 | 126 | 0·83 (0·70-0·99) | 0·85 (0·72-1·01) |
| Bangka Barat | 213163 | 217332 | 213282 | 203 | 154 | 174 | 95 | 71 | 82 | 0·74 (0·60-0·92) | 0·86 (0·70-1·05) |
| Bangka Tengah | 196196 | 200016 | 196060 | 254 | 155 | 190 | 129 | 77 | 97 | 0·60 (0·49-0·73) | 0·75 (0·62-0·90) |
| Bangka Selatan | 209973 | 213966 | 209815 | 234 | 127 | 153 | 111 | 59 | 73 | 0·53 (0·43-0·66) | 0·65 (0·53-0·80) |
| Belitung Timur | 129572 | 132069 | 129411 | 160 | 149 | 128 | 123 | 113 | 99 | 0·91 (0·73-1·14) | 0·80 (0·64-1·01) |
| Kota Pangkal Pinang | 212727 | 216893 | 212639 | 484 | 529 | 433 | 228 | 244 | 204 | 1·07 (0·95-1·21) | 0·90 (0·79-1·02) |
| **Riau Island** | | | | | | | | | | | |
| Karimun | 232800 | 253457 | 259450 | 504 | 342 | 392 | 216 | 135 | 151 | 0·62 (0·54-0·71) | 0·70 (0·61-0·80) |
| Bintan | 159400 | 159518 | 162560 | 273 | 211 | 248 | 171 | 132 | 153 | 0·77 (0·65-0·92) | 0·89 (0·75-1·06) |
| Natuna | 77770 | 81495 | 83360 | 109 | 65 | 60 | 140 | 80 | 72 | 0·57 (0·42-0·77) | 0·51 (0·38-0·70) |
| Lingga | 89780 | 98633 | 100660 | 124 | 109 | 112 | 138 | 111 | 111 | 0·80 (0·62-1·03) | 0·81 (0·62-1·04) |
| Kepulauan Anambas | 42310 | 47402 | 48740 | 89 | 59 | 37 | 210 | 124 | 76 | 0·59 (0·43-0·82) | 0·36 (0·25-0·52) |
| Kota Batam | 1376010 | 1196396 | 1230100 | 3645 | 2701 | 3034 | 265 | 226 | 247 | 0·85 (0·81-0·90) | 0·93 (0·89-0·98) |
| Kota Tanjung Pinang | 211580 | 227663 | 233370 | 744 | 478 | 581 | 352 | 210 | 249 | 0·60 (0·53-0·67) | 0·71 (0·64-0·79) |
| **DKI Jakarta** | | | | | | | | | | | |
| Kepulauan Seribu | 24300 | 27750 | 28240 | 73 | 48 | 51 | 300 | 173 | 181 | 0·58 (0·40-0·82) | 0·60 (0·42-0·86) |

| District | Number of populations | | | Number of new TB reported cases | | | Case notification rate (CNR) per 100,000 populations | | | CNR ratio for 2020 vs 2019 (95% CI) | CNR ratio for 2021 vs 2019 (95% CI) |
|---|---|---|---|---|---|---|---|---|---|---|---|
| | 2019 | 2020 | 2021 | 2019 | 2020 | 2021 | 2019 | 2020 | 2021 | | |
| Kota Jakarta Selatan | 2264700 | 2226810 | 2233855 | 8116 | 5440 | 6226 | 358 | 244 | 279 | 0·68 (0·66-0·71) | 0·78 (0·75-0·80) |
| Kota Jakarta Timur | 2937860 | 3037140 | 3056300 | 12764 | 8173 | 10763 | 434 | 269 | 352 | 0·62 (0·60-0·64) | 0·81 (0·79-0·83) |
| Kota Jakarta Pusat | 928110 | 1056900 | 1066460 | 7406 | 5205 | 6387 | 798 | 492 | 599 | 0·62 (0·60-0·64) | 0·75 (0·73-0·78) |
| Kota Jakarta Barat | 2589930 | 2434510 | 2440073 | 8374 | 4903 | 6758 | 323 | 201 | 277 | 0·62 (0·60-0·65) | 0·86 (0·83-0·88) |
| Kota Jakarta Utara | 1812910 | 1778980 | 1784753 | 5333 | 3620 | 5097 | 294 | 203 | 286 | 0·69 (0·66-0·72) | 0·97 (0·93-1·01) |
| **West Java** | | | | | | | | | | | |
| Bogor | 5965410 | 6088233 | 5489540 | 16769 | 11434 | 13466 | 281 | 188 | 245 | 0·67 (0·65-0·68) | 0·87 (0·85-0·89) |
| Cianjur | 2466272 | 2470219 | 2761480 | 6459 | 4014 | 5364 | 262 | 162 | 194 | 0·62 (0·60-0·65) | 0·74 (0·72-0·77) |
| Sukabumi | 2263072 | 2264328 | 2506680 | 5358 | 5015 | 5471 | 237 | 221 | 218 | 0·94 (0·90-0·97) | 0·92 (0·89-0·96) |
| Bandung | 3775279 | 3831505 | 3666160 | 7967 | 6284 | 6507 | 211 | 164 | 177 | 0·78 (0·75-0·80) | 0·84 (0·81-0·87) |
| Garut | 2622425 | 2636637 | 2604790 | 5095 | 4529 | 5551 | 194 | 172 | 213 | 0·88 (0·85-0·92) | 1·10 (1·06-1·14) |
| Tasikmalaya | 1754128 | 1755710 | 1883730 | 3016 | 1989 | 2451 | 172 | 113 | 130 | 0·66 (0·62-0·70) | 0·76 (0·72-0·80) |
| Ciamis | 1195176 | 1201685 | 1237730 | 1501 | 1569 | 1771 | 126 | 131 | 143 | 1·04 (0·97-1·12) | 1·14 (1·06-1·22) |
| Kuningan | 1080804 | 1087105 | 1180390 | 2463 | 1964 | 1975 | 228 | 181 | 167 | 0·79 (0·75-0·84) | 0·73 (0·69-0·78) |
| Cirebon | 2192903 | 2209633 | 2290970 | 7879 | 3498 | 3782 | 359 | 158 | 165 | 0·44 (0·42-0·46) | 0·46 (0·44-0·48) |
| Majalengka | 1205034 | 1210709 | 1318970 | 2317 | 1899 | 1999 | 192 | 157 | 152 | 0·82 (0·77-0·87) | 0·79 (0·74-0·84) |
| Sumedang | 1152400 | 1154428 | 1159350 | 2093 | 1780 | 1778 | 182 | 154 | 153 | 0·85 (0·80-0·90) | 0·84 (0·79-0·90) |
| Indramayu | 1728469 | 1737624 | 1851380 | 3210 | 1679 | 2097 | 186 | 97 | 113 | 0·52 (0·49-0·55) | 0·61 (0·58-0·64) |
| Subang | 1595825 | 1612576 | 1608590 | 3411 | 3353 | 3180 | 214 | 208 | 198 | 0·97 (0·93-1·02) | 0·93 (0·88-0·97) |
| Purwakarta | 962893 | 971889 | 191147 | 2119 | 1936 | 3027 | 220 | 199 | 1584 | 0·91 (0·85-0·96) | 7·20 (6·86-7·55) |
| Karawang | 2353915 | 2370488 | 2468580 | 6920 | 5171 | 6043 | 294 | 218 | 245 | 0·74 (0·72-0·77) | 0·83 (0·80-0·86) |
| Bekasi | 3763886 | 3899017 | 3157960 | 9728 | 4896 | 5441 | 258 | 126 | 172 | 0·49 (0·47-0·50) | 0·67 (0·65-0·69) |
| Bandung Barat | 1699896 | 1714982 | 1814230 | 2197 | 1547 | 1985 | 129 | 90 | 109 | 0·70 (0·65-0·74) | 0·85 (0·80-0·90) |
| Pangandaran | 399284 | 401493 | 427610 | 311 | 432 | 528 | 78 | 108 | 123 | 1·38 (1·19-1·60) | 1·59 (1·38-1·82) |

| District | Number of populations | | | Number of new TB reported cases | | | Case notification rate (CNR) per 100,000 populations | | | CNR ratio for 2020 vs 2019 (95% CI) | CNR ratio for 2021 vs 2019 (95% CI) |
|---|---|---|---|---|---|---|---|---|---|---|---|
| | 2019 | 2020 | 2021 | 2019 | 2020 | 2021 | 2019 | 2020 | 2021 | | |
| Kota Bogor | 1112081 | 1126927 | 1052360 | 3834 | 2378 | 4897 | 345 | 211 | 465 | 0·61 (0·58-0·64) | 1·35 (1·29-1·41) |
| Kota Sukabumi | 328680 | 330691 | 350800 | 2103 | 1250 | 1656 | 640 | 378 | 472 | 0·59 (0·55-0·63) | 0·74 (0·69-0·79) |
| Kota Bandung | 2507888 | 2510103 | 2452940 | 13038 | 9227 | 10749 | 520 | 368 | 438 | 0·71 (0·69-0·73) | 0·84 (0·82-0·86) |
| Kota Cirebon | 319312 | 322322 | 336860 | 1862 | 1525 | 2358 | 583 | 473 | 700 | 0·81 (0·76-0·87) | 1·20 (1·13-1·28) |
| Kota Bekasi | 3003923 | 3075690 | 2564940 | 11016 | 5876 | 7018 | 367 | 191 | 274 | 0·52 (0·51-0·54) | 0·75 (0·72-0·77) |
| Kota Depok | 2406826 | 2484186 | 2085940 | 6686 | 3427 | 4609 | 278 | 138 | 221 | 0·50 (0·48-0·52) | 0·80 (0·77-0·83) |
| Kota Cimahi | 614304 | 620393 | 571630 | 2557 | 2022 | 2078 | 416 | 326 | 364 | 0·78 (0·74-0·83) | 0·87 (0·82-0·93) |
| Kota Tasikmalaya | 663517 | 663986 | 723920 | 1671 | 1233 | 1729 | 252 | 186 | 239 | 0·74 (0·69-0·79) | 0·95 (0·89-1·01) |
| Kota Banjar | 183110 | 183299 | 203420 | 834 | 318 | 382 | 455 | 173 | 188 | 0·38 (0·34-0·43) | 0·41 (0·37-0·46) |
| **Central Java** | | | | | | | | | | | |
| Cilacap | 1727098 | 1944857 | 1963824 | 4077 | 2749 | 2851 | 236 | 141 | 145 | 0·60 (0·57-0·63) | 0·62 (0·59-0·64) |
| Banyumas | 1693006 | 1776918 | 1789630 | 4271 | 3608 | 4426 | 252 | 203 | 247 | 0·81 (0·77-0·84) | 0·98 (0·94-1·02) |
| Purbalingga | 933989 | 998561 | 1007794 | 1376 | 1016 | 1167 | 147 | 102 | 116 | 0·69 (0·64-0·75) | 0·79 (0·73-0·85) |
| Banjarnegara | 923192 | 1017767 | 1026866 | 1258 | 911 | 937 | 136 | 90 | 91 | 0·66 (0·60-0·71) | 0·67 (0·62-0·73) |
| Kebumen | 1197982 | 1350438 | 1361913 | 2491 | 1904 | 2225 | 208 | 141 | 163 | 0·68 (0·64-0·72) | 0·79 (0·74-0·83) |
| Purworejo | 718316 | 769880 | 773588 | 704 | 577 | 610 | 98 | 75 | 79 | 0·77 (0·69-0·85) | 0·81 (0·72-0·90) |
| Wonosobo | 790504 | 879124 | 886613 | 1857 | 1102 | 1387 | 235 | 125 | 156 | 0·53 (0·50-0·57) | 0·67 (0·62-0·71) |
| Magelang | 1290591 | 1299859 | 1305512 | 751 | 509 | 549 | 58 | 39 | 42 | 0·67 (0·60-0·75) | 0·72 (0·65-0·81) |
| Boyolali | 984807 | 1062713 | 1070247 | 1452 | 427 | 561 | 147 | 40 | 52 | 0·27 (0·25-0·30) | 0·36 (0·32-0·39) |
| Klaten | 1174986 | 1260506 | 1267272 | 1282 | 681 | 1094 | 109 | 54 | 86 | 0·50 (0·45-0·54) | 0·79 (0·73-0·86) |
| Sukoharjo | 891912 | 907587 | 911603 | 749 | 639 | 734 | 84 | 70 | 81 | 0·84 (0·75-0·93) | 0·96 (0·87-1·06) |
| Wonogiri | 959492 | 1043177 | 1049292 | 1077 | 683 | 684 | 112 | 65 | 65 | 0·58 (0·53-0·64) | 0·58 (0·53-0·64) |
| Karanganyar | 886519 | 931963 | 938808 | 571 | 414 | 352 | 64 | 44 | 37 | 0·69 (0·61-0·78) | 0·58 (0·51-0·66) |
| Sragen | 890518 | 976951 | 983641 | 960 | 570 | 523 | 108 | 58 | 53 | 0·54 (0·49-0·60) | 0·49 (0·44-0·55) |

| District | Number of populations | | | Number of new TB reported cases | | | Case notification rate (CNR) per 100,000 populations | | | CNR ratio for 2020 vs 2019 (95% CI) | CNR ratio for 2021 vs 2019 (95% CI) |
|---|---|---|---|---|---|---|---|---|---|---|---|
| | **2019** | **2020** | **2021** | **2019** | **2020** | **2021** | **2019** | **2020** | **2021** | | |
| Grobogan | 1377788 | 1453526 | 1460873 | 1094 | 908 | 991 | 79 | 62 | 68 | 0·79 (0·72-0·86) | 0·85 (0·78-0·93) |
| Blora | 865013 | 884333 | 886147 | 1366 | 975 | 935 | 158 | 110 | 106 | 0·70 (0·64-0·76) | 0·67 (0·62-0·73) |
| Rembang | 638188 | 645333 | 647766 | 972 | 578 | 655 | 152 | 90 | 101 | 0·59 (0·53-0·65) | 0·66 (0·60-0·73) |
| Pati | 1259590 | 1324188 | 1330983 | 1770 | 1615 | 1685 | 141 | 122 | 127 | 0·87 (0·81-0·93) | 0·90 (0·84-0·96) |
| Kudus | 871311 | 849184 | 852443 | 1610 | 1239 | 1899 | 185 | 146 | 223 | 0·79 (0·73-0·85) | 1·21 (1·13-1·29) |
| Jepara | 1257912 | 1184947 | 1188510 | 1182 | 942 | 938 | 94 | 79 | 79 | 0·85 (0·78-0·92) | 0·84 (0·77-0·91) |
| Demak | 1162805 | 1203956 | 1212377 | 1488 | 1432 | 1113 | 128 | 119 | 92 | 0·93 (0·86-1·00) | 0·72 (0·66-0·78) |
| Semarang | 1053786 | 1053094 | 1059844 | 993 | 632 | 663 | 94 | 60 | 63 | 0·64 (0·58-0·70) | 0·66 (0·60-0·73) |
| Temanggung | 772018 | 790174 | 794403 | 650 | 483 | 543 | 84 | 61 | 68 | 0·73 (0·65-0·82) | 0·81 (0·72-0·91) |
| Kendal | 971086 | 1018505 | 1025020 | 1507 | 1386 | 1406 | 155 | 136 | 137 | 0·88 (0·82-0·94) | 0·88 (0·82-0·95) |
| Batang | 768583 | 801718 | 807005 | 1204 | 862 | 912 | 157 | 108 | 113 | 0·69 (0·63-0·75) | 0·72 (0·66-0·79) |
| Pekalongan | 897711 | 968821 | 976504 | 1255 | 1101 | 1426 | 140 | 114 | 146 | 0·81 (0·75-0·88) | 1·05 (0·97-1·13) |
| Pemalang | 1302813 | 1471489 | 1484209 | 1899 | 1445 | 1790 | 146 | 98 | 121 | 0·67 (0·63-0·72) | 0·83 (0·78-0·88) |
| Tegal | 1440698 | 1596996 | 1608611 | 3855 | 3022 | 3075 | 268 | 189 | 191 | 0·71 (0·67-0·74) | 0·71 (0·68-0·75) |
| Brebes | 1809096 | 1978759 | 1992685 | 3028 | 2057 | 2825 | 167 | 104 | 142 | 0·62 (0·59-0·66) | 0·85 (0·80-0·89) |
| Kota Magelang | 122111 | 121526 | 121610 | 1070 | 883 | 951 | 876 | 727 | 782 | 0·83 (0·76-0·91) | 0·89 (0·82-0·97) |
| Kota Surakarta | 519587 | 522364 | 522728 | 1902 | 1303 | 1523 | 366 | 249 | 291 | 0·68 (0·64-0·73) | 0·80 (0·74-0·85) |
| Kota Salatiga | 194084 | 192322 | 193525 | 855 | 503 | 588 | 441 | 262 | 304 | 0·59 (0·53-0·66) | 0·69 (0·62-0·77) |
| Kota Semarang | 1814110 | 1653524 | 1656564 | 5403 | 2876 | 3878 | 298 | 174 | 234 | 0·58 (0·56-0·61) | 0·79 (0·75-0·82) |
| Kota Pekalongan | 307097 | 307150 | 308310 | 826 | 839 | 886 | 269 | 273 | 287 | 1·02 (0·92-1·12) | 1·07 (0·97-1·17) |
| Kota Tegal | 249905 | 273825 | 275781 | 2463 | 2245 | 2512 | 986 | 820 | 911 | 0·83 (0·79-0·88) | 0·92 (0·87-0·98) |
| **Di Yogyakarta** | | | | | | | | | | | |
| Kulon Progo | 432058 | 437373 | 442724 | 323 | 240 | 248 | 75 | 55 | 56 | 0·73 (0·62-0·87) | 0·75 (0·64-0·88) |
| Bantul | 1022788 | 1036489 | 1050308 | 1072 | 752 | 793 | 105 | 73 | 76 | 0·69 (0·63-0·76) | 0·72 (0·66-0·79) |

| District | Number of populations | | | Number of new TB reported cases | | | Case notification rate (CNR) per 100,000 populations | | | CNR ratio for 2020 vs 2019 (95% CI) | CNR ratio for 2021 vs 2019 (95% CI) |
|---|---|---|---|---|---|---|---|---|---|---|---|
| | 2019 | 2020 | 2021 | 2019 | 2020 | 2021 | 2019 | 2020 | 2021 | | |
| Gunung Kidul | 749229 | 758316 | 767464 | 438 | 328 | 286 | 58 | 43 | 37 | 0·74 (0·64-0·85) | 0·64 (0·55-0·74) |
| Sleman | 1231246 | 1248258 | 1256429 | 1208 | 1177 | 1255 | 98 | 94 | 100 | 0·96 (0·89-1·04) | 1·02 (0·94-1·10) |
| Kota Yogyakarta | 433267 | 438761 | 444295 | 1109 | 962 | 1048 | 256 | 219 | 236 | 0·86 (0·79-0·93) | 0·92 (0·85-1·00) |
| **East Java** | | | | | | | | | | | |
| Pacitan | 555304 | 586110 | 589108 | 362 | 292 | 220 | 65 | 50 | 37 | 0·76 (0·66-0·89) | 0·57 (0·49-0·68) |
| Ponorogo | 871370 | 949318 | 955839 | 1158 | 992 | 762 | 133 | 104 | 80 | 0·79 (0·72-0·86) | 0·60 (0·55-0·66) |
| Trenggalek | 696295 | 731125 | 734888 | 551 | 405 | 250 | 79 | 55 | 34 | 0·70 (0·62-0·80) | 0·43 (0·37-0·50) |
| Tulungagung | 1039284 | 1089775 | 1096588 | 1283 | 963 | 808 | 123 | 88 | 74 | 0·72 (0·66-0·78) | 0·60 (0·55-0·65) |
| Blitar | 1160677 | 1223745 | 1231013 | 856 | 580 | 485 | 74 | 47 | 39 | 0·64 (0·58-0·71) | 0·53 (0·48-0·60) |
| Kediri | 1574272 | 1635294 | 1644400 | 2013 | 1645 | 1374 | 128 | 101 | 84 | 0·79 (0·74-0·84) | 0·65 (0·61-0·70) |
| Malang | 2606204 | 2654448 | 2668296 | 2794 | 2084 | 1985 | 107 | 79 | 74 | 0·73 (0·69-0·77) | 0·69 (0·66-0·73) |
| Lumajang | 1042395 | 1119251 | 1127094 | 1924 | 1316 | 1467 | 185 | 118 | 130 | 0·64 (0·59-0·68) | 0·71 (0·66-0·75) |
| Jember | 2450668 | 2536729 | 2550360 | 4355 | 3626 | 3562 | 178 | 143 | 140 | 0·80 (0·77-0·84) | 0·79 (0·75-0·82) |
| Banyuwangi | 1613991 | 1708114 | 1718462 | 2776 | 2094 | 2144 | 172 | 123 | 125 | 0·71 (0·67-0·75) | 0·73 (0·69-0·77) |
| Bondowoso | 775715 | 776151 | 778525 | 1442 | 878 | 1035 | 186 | 113 | 133 | 0·61 (0·56-0·66) | 0·72 (0·66-0·77) |
| Situbondo | 682978 | 685967 | 688337 | 1249 | 1079 | 1109 | 183 | 157 | 161 | 0·86 (0·79-0·93) | 0·88 (0·81-0·96) |
| Probolinggo | 1168503 | 1152537 | 1155894 | 1785 | 1182 | 1305 | 153 | 103 | 113 | 0·67 (0·62-0·72) | 0·74 (0·69-0·79) |
| Pasuruan | 1627396 | 1605969 | 1611805 | 3516 | 1945 | 2164 | 216 | 121 | 134 | 0·56 (0·53-0·59) | 0·62 (0·59-0·66) |
| Sidoarjo | 2249476 | 2082801 | 2091930 | 4009 | 2599 | 3231 | 178 | 125 | 154 | 0·70 (0·67-0·74) | 0·87 (0·83-0·91) |
| Mojokerto | 1117688 | 1119209 | 1125522 | 1563 | 989 | 1147 | 140 | 88 | 102 | 0·63 (0·58-0·68) | 0·73 (0·68-0·79) |
| Jombang | 1263814 | 1318062 | 1325914 | 1730 | 1299 | 1369 | 137 | 99 | 103 | 0·72 (0·67-0·77) | 0·75 (0·70-0·81) |
| Nganjuk | 1054611 | 1103902 | 1109683 | 1094 | 744 | 738 | 104 | 67 | 67 | 0·65 (0·59-0·71) | 0·64 (0·58-0·70) |
| Madiun | 682684 | 744350 | 750143 | 1234 | 673 | 648 | 181 | 90 | 86 | 0·50 (0·46-0·55) | 0·48 (0·44-0·52) |
| Magetan | 628977 | 670812 | 674133 | 766 | 506 | 427 | 122 | 75 | 63 | 0·62 (0·55-0·69) | 0·52 (0·46-0·58) |

| District | Number of populations | | | Number of new TB reported cases | | | Case notification rate (CNR) per 100,000 populations | | | CNR ratio for 2020 vs 2019 (95% CI) | CNR ratio for 2021 vs 2019 (95% CI) |
|---|---|---|---|---|---|---|---|---|---|---|---|
| | 2019 | 2020 | 2021 | 2019 | 2020 | 2021 | 2019 | 2020 | 2021 | | |
| Ngawi | 830108 | 870057 | 873346 | 1045 | 736 | 692 | 126 | 85 | 79 | 0·67 (0·61-0·74) | 0·63 (0·57-0·69) |
| Bojonegoro | 1249692 | 1301635 | 1307602 | 1844 | 1463 | 1455 | 148 | 112 | 111 | 0·76 (0·71-0·82) | 0·75 (0·70-0·81) |
| Tuban | 1172790 | 1198012 | 1203127 | 2007 | 1411 | 1333 | 171 | 118 | 111 | 0·69 (0·64-0·74) | 0·65 (0·60-0·69) |
| Lamongan | 1189106 | 1344165 | 1356027 | 2244 | 1627 | 1924 | 189 | 121 | 142 | 0·64 (0·60-0·68) | 0·75 (0·71-0·80) |
| Gresik | 1312881 | 1311215 | 1320570 | 2642 | 1620 | 2023 | 201 | 124 | 153 | 0·61 (0·58-0·65) | 0·76 (0·72-0·81) |
| Bangkalan | 986672 | 1060377 | 1071712 | 1443 | 1142 | 1192 | 146 | 108 | 111 | 0·74 (0·68-0·80) | 0·76 (0·70-0·82) |
| Sampang | 978875 | 969694 | 976020 | 1106 | 768 | 969 | 113 | 79 | 99 | 0·70 (0·64-0·77) | 0·88 (0·81-0·96) |
| Pamekasan | 879992 | 850057 | 853507 | 1118 | 1025 | 1053 | 127 | 121 | 123 | 0·95 (0·87-1·03) | 0·97 (0·89-1·06) |
| Sumenep | 1088910 | 1124436 | 1129822 | 1864 | 1606 | 1625 | 171 | 143 | 144 | 0·83 (0·78-0·89) | 0·84 (0·79-0·90) |
| Kota Kediri | 287409 | 286796 | 287962 | 865 | 738 | 768 | 301 | 257 | 267 | 0·86 (0·78-0·94) | 0·89 (0·80-0·98) |
| Kota Blitar | 141876 | 149149 | 150371 | 280 | 291 | 257 | 197 | 195 | 171 | 0·99 (0·84-1·16) | 0·87 (0·73-1·03) |
| Kota Malang | 870682 | 843810 | 844933 | 2197 | 1400 | 1593 | 252 | 166 | 189 | 0·66 (0·62-0·70) | 0·75 (0·70-0·80) |
| Kota Probolinggo | 237208 | 239649 | 241202 | 790 | 419 | 457 | 333 | 175 | 189 | 0·53 (0·47-0·59) | 0·57 (0·51-0·64) |
| Kota Pasuruan | 200422 | 208006 | 209528 | 580 | 591 | 706 | 289 | 284 | 337 | 0·98 (0·88-1·10) | 1·16 (1·04-1·30) |
| Kota Mojokerto | 129014 | 132434 | 133272 | 453 | 540 | 629 | 351 | 408 | 472 | 1·16 (1·03-1·32) | 1·34 (1·19-1·52) |
| Kota Madiun | 177007 | 195175 | 196917 | 734 | 655 | 599 | 415 | 336 | 304 | 0·81 (0·73-0·90) | 0·73 (0·66-0·82) |
| Kota Surabaya | 2896195 | 2874314 | 2880284 | 8805 | 4456 | 6182 | 304 | 155 | 215 | 0·51 (0·49-0·53) | 0·71 (0·68-0·73) |
| Kota Batu | 207490 | 213046 | 214653 | 239 | 261 | 187 | 115 | 123 | 87 | 1·06 (0·89-1·27) | 0·76 (0·62-0·92) |
| **Banten** | | | | | | | | | | | |
| Pandeglang | 1211909 | 1272687 | 1288314 | 2331 | 2184 | 2224 | 192 | 172 | 173 | 0·89 (0·84-0·95) | 0·90 (0·85-0·95) |
| Lebak | 1302608 | 1386793 | 1407857 | 2977 | 2023 | 2561 | 229 | 146 | 182 | 0·64 (0·60-0·68) | 0·80 (0·76-0·84) |
| Tangerang | 3800787 | 3245619 | 3293533 | 7859 | 5937 | 7546 | 207 | 183 | 229 | 0·89 (0·86-0·91) | 1·11 (1·07-1·14) |
| Serang | 1508397 | 1622630 | 1647790 | 4019 | 2731 | 3371 | 266 | 168 | 205 | 0·63 (0·60-0·66) | 0·77 (0·73-0·80) |
| Kota Tangerang | 2229901 | 1895486 | 1911914 | 6174 | 4479 | 6017 | 277 | 236 | 315 | 0·85 (0·82-0·89) | 1·14 (1·10-1·18) |

| District | Number of populations | | | Number of new TB reported cases | | | Case notification rate (CNR) per 100,000 populations | | | CNR ratio for 2020 vs 2019 (95% CI) | CNR ratio for 2021 vs 2019 (95% CI) |
|---|---|---|---|---|---|---|---|---|---|---|---|
| | 2019 | 2020 | 2021 | 2019 | 2020 | 2021 | 2019 | 2020 | 2021 | | |
| Kota Cilegon | 437205 | 434896 | 441761 | 1292 | 1200 | 1162 | 296 | 276 | 263 | 0·93 (0·86-1·01) | 0·89 (0·82-0·96) |
| Kota Serang | 688603 | 692101 | 704618 | 2299 | 1315 | 1540 | 334 | 190 | 219 | 0·57 (0·53-0·61) | 0·66 (0·61-0·70) |
| Kota Tangerang Selatan | 1747906 | 1354350 | 1365688 | 4064 | 2782 | 3491 | 233 | 205 | 256 | 0·88 (0·84-0·93) | 1·10 (1·05-1·15) |
| **Bali** | | | | | | | | | | | |
| Jembrana | 278700 | 317100 | 321900 | 194 | 128 | 148 | 70 | 40 | 46 | 0·58 (0·47-0·72) | 0·66 (0·53-0·82) |
| Tabanan | 446700 | 461600 | 465300 | 277 | 182 | 190 | 62 | 39 | 41 | 0·64 (0·53-0·77) | 0·66 (0·55-0·79) |
| Badung | 678900 | 548200 | 549300 | 587 | 379 | 385 | 86 | 69 | 70 | 0·80 (0·70-0·91) | 0·81 (0·71-0·92) |
| Gianyar | 514300 | 515300 | 519500 | 347 | 224 | 250 | 67 | 43 | 48 | 0·64 (0·55-0·76) | 0·71 (0·61-0·84) |
| Klungkung | 179100 | 206900 | 210100 | 105 | 131 | 127 | 59 | 63 | 60 | 1·08 (0·84-1·40) | 1·03 (0·61-0·84) |
| Bangli | 227600 | 258700 | 262500 | 88 | 27 | 53 | 39 | 10 | 20 | 0·27 (0·18-0·40) | 0·52 (0·37-0·73) |
| Karangasem | 417000 | 492400 | 500800 | 322 | 193 | 223 | 77 | 39 | 45 | 0·51 (0·43-0·60) | 0·58 (0·49-0·68) |
| Buleleng | 661900 | 791800 | 806600 | 663 | 528 | 691 | 100 | 67 | 86 | 0·67 (0·59-0·75) | 0·86 (0·77-0·95) |
| Kota Denpasar | 957800 | 725300 | 726600 | 1622 | 1215 | 1306 | 169 | 168 | 180 | 0·99 (0·92-1·07) | 1·06 (0·99-1·14) |
| **West Nusa Tenggara** | | | | | | | | | | | |
| Lombok Barat | 694985 | 704586 | 731800 | 1090 | 742 | 926 | 157 | 105 | 127 | 0·67 (0·61-0·74) | 0·81 (0·74-0·88) |
| Lombok Tengah | 947488 | 955411 | 1049700 | 1096 | 909 | 976 | 116 | 95 | 93 | 0·82 (0·75-0·90) | 0·80 (0·74-0·88) |
| Lombok Timur | 1200612 | 1208594 | 1343900 | 1501 | 1273 | 1566 | 125 | 105 | 117 | 0·84 (0·78-0·91) | 0·93 (0·87-1·00) |
| Sumbawa | 457671 | 461502 | 519800 | 704 | 447 | 504 | 154 | 97 | 97 | 0·63 (0·56-0·71) | 0·63 (0·56-0·71) |
| Dompu | 252288 | 255569 | 238200 | 379 | 297 | 365 | 150 | 116 | 153 | 0·77 (0·66-0·90) | 1·02 (0·88-1·18) |
| Bima | 488577 | 493198 | 520400 | 717 | 525 | 636 | 147 | 106 | 122 | 0·73 (0·65-0·81) | 0·83 (0·75-0·93) |
| Sumbawa Barat | 148606 | 152437 | 148500 | 276 | 203 | 204 | 186 | 133 | 137 | 0·72 (0·60-0·86) | 0·74 (0·62-0·89) |
| Lombok Utara | 220412 | 222212 | 251500 | 320 | 295 | 251 | 145 | 133 | 100 | 0·91 (0·78-1·07) | 0·69 (0·58-0·81) |
| Kota Mataram | 486715 | 495681 | 432000 | 927 | 850 | 941 | 190 | 171 | 218 | 0·90 (0·82-0·99) | 1·14 (1·04-1·25) |
| Kota Bima | 173031 | 176432 | 156200 | 262 | 210 | 319 | 151 | 119 | 204 | 0·79 (0·66-0·94) | 1·35 (1·15-1·59) |

| District | Number of populations | | | Number of new TB reported cases | | | Case notification rate (CNR) per 100,000 populations | | | CNR ratio for 2020 vs 2019 (95% CI) | CNR ratio for 2021 vs 2019 (95% CI) |
|---|---|---|---|---|---|---|---|---|---|---|---|
| | 2019 | 2020 | 2021 | 2019 | 2020 | 2021 | 2019 | 2020 | 2021 | | |
| **East Nusa Tenggara** | | | | | | | | | | | |
| Sumba Barat | 129710 | 131600 | 148252 | 406 | 270 | 276 | 313 | 205 | 186 | 0·66 (0·56-0·76) | 0·60 (0·51-0·69) |
| Sumba Timur | 258486 | 261503 | 246618 | 394 | 245 | 257 | 152 | 94 | 104 | 0·62 (0·52-0·72) | 0·68 (0·58-0·80) |
| Kupang | 403582 | 421618 | 372101 | 491 | 332 | 317 | 122 | 79 | 85 | 0·65 (0·56-0·74) | 0·70 (0·61-0·81) |
| Timor Tengah Selatan | 467990 | 469673 | 457406 | 528 | 417 | 416 | 113 | 89 | 91 | 0·79 (0·69-0·89) | 0·81 (0·71-0·92) |
| Timor Tengah Utara | 254171 | 256299 | 262698 | 235 | 247 | 191 | 92 | 96 | 73 | 1·04 (0·87-1·25) | 0·79 (0·65-0·95) |
| Belu | 220115 | 223176 | 220764 | 693 | 453 | 381 | 315 | 203 | 173 | 0·65 (0·57-0·73) | 0·55 (0·48-0·62) |
| Alor | 205599 | 206806 | 213994 | 246 | 274 | 289 | 120 | 132 | 135 | 1·11 (0·93-1·32) | 1·13 (0·95-1·34) |
| Lembata | 143074 | 145685 | 137631 | 201 | 94 | 113 | 140 | 65 | 82 | 0·46 (0·36-0·58) | 0·58 (0·47-0·73) |
| Flores Timur | 255916 | 257785 | 281001 | 227 | 229 | 188 | 89 | 89 | 67 | 1·00 (0·83-1·20) | 0·75 (0·62-0·91) |
| Sikka | 320401 | 321790 | 324252 | 460 | 394 | 499 | 144 | 122 | 154 | 0·85 (0·75-0·98) | 1·07 (0·94-1·22) |
| Ende | 273929 | 274599 | 272078 | 663 | 237 | 346 | 242 | 86 | 127 | 0·36 (0·31-0·41) | 0·53 (0·46-0·60) |
| Ngada | 163217 | 165314 | 167396 | 171 | 116 | 143 | 105 | 70 | 85 | 0·67 (0·53-0·85) | 0·82 (0·65-1·02) |
| Manggarai | 338424 | 342908 | 315041 | 359 | 242 | 230 | 106 | 71 | 73 | 0·67 (0·57-0·78) | 0·69 (0·58-0·81) |
| Rote Ndao | 172104 | 178805 | 145972 | 120 | 100 | 58 | 70 | 56 | 40 | 0·80 (0·62-1·05) | 0·57 (0·42-0·78) |
| Manggarai Barat | 274689 | 280412 | 259566 | 326 | 263 | 273 | 119 | 94 | 105 | 0·79 (0·67-0·93) | 0·89 (0·75-1·04) |
| Sumba Tengah | 72800 | 73820 | 87630 | 114 | 77 | 92 | 157 | 104 | 105 | 0·67 (0·50-0·89) | 0·67 (0·51-0·88) |
| Sumba Barat Daya | 344720 | 350923 | 305689 | 472 | 550 | 832 | 137 | 157 | 272 | 1·15 (1·01-1·29) | 1·99 (1·78-2·22) |
| Nagekeo | 145826 | 147189 | 162463 | 162 | 151 | 126 | 111 | 103 | 78 | 0·92 (0·74-1·15) | 0·70 (0·55-0·88) |
| Manggarai Timur | 287207 | 289836 | 277914 | 205 | 143 | 191 | 71 | 49 | 69 | 0·69 (0·56-0·85) | 0·96 (0·79-1·17) |
| Sabu Raijua | 97379 | 100684 | 90837 | 85 | 86 | 51 | 87 | 85 | 56 | 0·98 (0·73-1·32) | 0·64 (0·46-0·91) |
| Malaka | 191892 | 194776 | 185809 | 370 | 379 | 376 | 193 | 195 | 202 | 1·01 (0·87-1·16) | 1·05 (0·91-1·21) |
| Kota Kupang | 434972 | 446193 | 452626 | 667 | 567 | 613 | 153 | 127 | 135 | 0·83 (0·74-0·93) | 0·88 (0·79-0·99) |
| **West Kalimantan** | | | | | | | | | | | |

| District | Number of populations | | | Number of new TB reported cases | | | Case notification rate (CNR) per 100,000 populations | | | CNR ratio for 2020 vs 2019 (95% CI) | CNR ratio for 2021 vs 2019 (95% CI) |
|---|---|---|---|---|---|---|---|---|---|---|---|
| | 2019 | 2020 | 2021 | 2019 | 2020 | 2021 | 2019 | 2020 | 2021 | | |
| Sambas | 535725 | 629905 | 637811 | 1039 | 858 | 1171 | 194 | 136 | 184 | 0·70 (0·64-0·77) | 0·95 (0·87-1·03) |
| Bengkayang | 255261 | 286366 | 290943 | 331 | 416 | 451 | 130 | 145 | 155 | 1·12 (0·97-1·29) | 1·20 (1·04-1·38) |
| Landak | 377305 | 397610 | 401103 | 561 | 383 | 391 | 149 | 96 | 97 | 0·65 (0·57-0·74) | 0·66 (0·58-0·75) |
| Mempawah | 264225 | 301560 | 305673 | 360 | 285 | 274 | 136 | 95 | 90 | 0·69 (0·59-0·81) | 0·66 (0·56-0·77) |
| Sanggau | 470224 | 484836 | 488527 | 836 | 700 | 651 | 178 | 144 | 133 | 0·81 (0·73-0·90) | 0·75 (0·68-0·83) |
| Ketapang | 512783 | 570657 | 579927 | 660 | 611 | 666 | 129 | 107 | 115 | 0·83 (0·75-0·93) | 0·89 (0·80-0·99) |
| Sintang | 418785 | 421306 | 423674 | 1086 | 396 | 508 | 259 | 94 | 120 | 0·36 (0·32-0·40) | 0·46 (0·42-0·51) |
| Kapuas Hulu | 263207 | 252609 | 253740 | 459 | 69 | 377 | 174 | 27 | 149 | 0·16 (0·13-0·20) | 0·85 (0·74-0·98) |
| Sekadau | 201578 | 211559 | 212878 | 313 | 185 | 255 | 155 | 87 | 120 | 0·56 (0·47-0·67) | 0·77 (0·65-0·91) |
| Melawi | 208417 | 228270 | 231242 | 468 | 335 | 453 | 225 | 147 | 196 | 0·65 (0·57-0·75) | 0·87 (0·77-0·99) |
| Kayong Utara | 112715 | 126571 | 128550 | 194 | 81 | 129 | 172 | 64 | 100 | 0·37 (0·29-0·48) | 0·58 (0·47-0·73) |
| Kubu Raya | 579331 | 609392 | 615125 | 610 | 496 | 580 | 105 | 81 | 94 | 0·77 (0·69-0·87) | 0·90 (0·80-1·00) |
| Kota Pontianak | 646661 | 658685 | 663713 | 1728 | 1466 | 1908 | 267 | 223 | 287 | 0·83 (0·78-0·89) | 1·08 (1·01-1·15) |
| Kota Singkawang | 222910 | 235064 | 237891 | 884 | 859 | 932 | 397 | 365 | 392 | 0·92 (0·84-1·01) | 0·99 (0·90-1·08) |
| **Central Kalimantan** | | | | | | | | | | | |
| Kotawaringin Barat | 312911 | 270388 | 272531 | 526 | 320 | 393 | 168 | 118 | 144 | 0·70 (0·61-0·81) | 0·86 (0·75-0·98) |
| Kotawaringin Timur | 466366 | 428895 | 432283 | 555 | 474 | 508 | 119 | 111 | 118 | 0·93 (0·82-1·05) | 0·99 (0·88-1·11) |
| Kapuas | 358820 | 410446 | 416181 | 375 | 257 | 295 | 105 | 63 | 71 | 0·60 (0·51-0·70) | 0·68 (0·58-0·79) |
| Barito Selatan | 136796 | 131140 | 131606 | 211 | 149 | 187 | 154 | 114 | 142 | 0·74 (0·60-0·91) | 0·92 (0·76-1·12) |
| Barito Utara | 130713 | 154812 | 157231 | 252 | 146 | 159 | 193 | 94 | 101 | 0·49 (0·40-0·60) | 0·53 (0·43-0·64) |
| Sukamara | 64342 | 63464 | 64941 | 98 | 81 | 87 | 152 | 128 | 134 | 0·84 (0·62-1·12) | 0·88 (0·66-1·17) |
| Lamandau | 82680 | 97611 | 100535 | 154 | 91 | 115 | 186 | 93 | 114 | 0·50 (0·39-0·65) | 0·61 (0·48-0·78) |
| Seruyan | 205880 | 162906 | 164378 | 208 | 150 | 353 | 101 | 92 | 215 | 0·91 (0·74-1·12) | 2·13 (1·80-2·51) |
| Katingan | 169997 | 162222 | 163099 | 208 | 131 | 188 | 122 | 81 | 115 | 0·66 (0·53-0·82) | 0·94 (0·77-1·15) |

| District | Number of populations | | | Number of new TB reported cases | | | Case notification rate (CNR) per 100,000 populations | | | CNR ratio for 2020 vs 2019 (95% CI) | CNR ratio for 2021 vs 2019 (95% CI) |
|---|---|---|---|---|---|---|---|---|---|---|---|
| | 2019 | 2020 | 2021 | 2019 | 2020 | 2021 | 2019 | 2020 | 2021 | | |
| Pulang Pisau | 127910 | 134499 | 135336 | 116 | 59 | 75 | 91 | 44 | 55 | 0·48 (0·36-0·66) | 0·61 (0·46-0·81) |
| Gunung Mas | 119910 | 135373 | 138407 | 178 | 154 | 132 | 148 | 114 | 95 | 0·77 (0·62-0·95) | 0·64 (0·51-0·80) |
| Barito Timur | 126874 | 113229 | 114243 | 100 | 81 | 131 | 79 | 72 | 115 | 0·91 (0·68-1·22) | 1·46 (1·12-1·88) |
| Murung Raya | 120785 | 111527 | 112445 | 244 | 205 | 202 | 202 | 184 | 180 | 0·91 (0·76-1·10) | 0·89 (0·74-1·07) |
| Kota Palangka Raya | 291785 | 293457 | 298954 | 623 | 279 | 619 | 214 | 95 | 207 | 0·45 (0·39-0·51) | 0·97 (0·87-1·08) |
| **South Kalimantan** | | | | | | | | | | | |
| Tanah Laut | 343890 | 348966 | 354340 | 448 | 218 | 309 | 130 | 62 | 87 | 0·48 (0·41-0·56) | 0·67 (0·58-0·77) |
| Kotabaru | 342217 | 325622 | 329483 | 365 | 268 | 369 | 107 | 82 | 112 | 0·77 (0·66-0·90) | 1·05 (0·91-1·21) |
| Banjar | 588066 | 565635 | 572109 | 1271 | 608 | 657 | 216 | 107 | 115 | 0·50 (0·45-0·55) | 0·53 (0·48-0·58) |
| Barito Kuala | 313595 | 313021 | 316963 | 269 | 205 | 235 | 86 | 65 | 74 | 0·76 (0·64-0·92) | 0·86 (0·73-1·03) |
| Tapin | 191372 | 189475 | 191801 | 282 | 125 | 172 | 147 | 66 | 90 | 0·45 (0·36-0·55) | 0·61 (0·50-0·73) |
| Hulu Sungai Selatan | 237702 | 228006 | 229960 | 421 | 174 | 290 | 177 | 76 | 126 | 0·43 (0·36-0·51) | 0·71 (0·61-0·83) |
| Hulu Sungai Tengah | 272419 | 258721 | 260754 | 412 | 370 | 436 | 151 | 143 | 167 | 0·95 (0·82-1·09) | 1·11 (0·97-1·26) |
| Hulu Sungai Utara | 237573 | 226727 | 228831 | 329 | 206 | 302 | 138 | 91 | 132 | 0·66 (0·55-0·78) | 0·95 (0·82-1·11) |
| Tabalong | 254322 | 253305 | 256903 | 360 | 206 | 265 | 142 | 81 | 103 | 0·58 (0·49-0·68) | 0·73 (0·62-0·85) |
| Tanah Bumbu | 360187 | 322646 | 328146 | 341 | 161 | 254 | 95 | 50 | 77 | 0·53 (0·44-0·63) | 0·82 (0·70-0·96) |
| Balangan | 131428 | 130355 | 132213 | 184 | 69 | 149 | 140 | 53 | 113 | 0·38 (0·29-0·49) | 0·81 (0·65-1·00) |
| Kota Banjarmasin | 708606 | 657663 | 662320 | 2155 | 886 | 1239 | 304 | 135 | 187 | 0·44 (0·41-0·48) | 0·62 (0·57-0·66) |
| Kota Banjar Baru | 262719 | 253442 | 258753 | 468 | 269 | 305 | 178 | 106 | 118 | 0·60 (0·51-0·69) | 0·66 (0·57-0·76) |
| **East Kalimantan** | | | | | | | | | | | |
| Paser | 285894 | 275452 | 277602 | 462 | 368 | 346 | 162 | 134 | 125 | 0·83 (0·72-0·95) | 0·77 (0·67-0·89) |
| Kutai Barat | 148020 | 172288 | 173982 | 359 | 177 | 310 | 243 | 103 | 178 | 0·42 (0·36-0·50) | 0·74 (0·63-0·85) |
| Kutai Kartanegara | 786122 | 729382 | 733626 | 956 | 601 | 773 | 122 | 82 | 105 | 0·68 (0·61-0·75) | 0·87 (0·79-0·95) |
| Kutai Timur | 376111 | 434459 | 449161 | 631 | 447 | 476 | 168 | 103 | 106 | 0·61 (0·54-0·69) | 0·63 (0·56-0·71) |

| District | Number of populations | | | Number of new TB reported cases | | | Case notification rate (CNR) per 100,000 populations | | | CNR ratio for 2020 vs 2019 (95% CI) | CNR ratio for 2021 vs 2019 (95% CI) |
|---|---|---|---|---|---|---|---|---|---|---|---|
| | 2019 | 2020 | 2021 | 2019 | 2020 | 2021 | 2019 | 2020 | 2021 | | |
| Berau | 232287 | 248035 | 252648 | 498 | 234 | 281 | 214 | 94 | 111 | 0·44 (0·38-0·51) | 0·52 (0·45-0·60) |
| Penajam Paser Utara | 160912 | 178681 | 180657 | 223 | 214 | 193 | 139 | 120 | 107 | 0·86 (0·72-1·04) | 0·77 (0·64-0·93) |
| Mahakam Hulu | 26375 | 32513 | 32969 | 48 | 39 | 39 | 182 | 120 | 118 | 0·66 (0·43-1·00) | 0·65 (0·43-0·99) |
| Kota Balikpapan | 655178 | 688318 | 695287 | 1797 | 952 | 1387 | 274 | 138 | 199 | 0·50 (0·47-0·54) | 0·73 (0·68-0·78) |
| Kota Samarinda | 872768 | 827994 | 831460 | 1864 | 1726 | 1979 | 214 | 208 | 238 | 0·98 (0·91-1·04) | 1·11 (1·05-1·19) |
| Kota Bontang | 177722 | 178917 | 180843 | 899 | 581 | 547 | 506 | 325 | 302 | 0·64 (0·58-0·71) | 0·60 (0·54-0·66) |
| **North Kalimantan** | | | | | | | | | | | |
| Malinau | 84609 | 82500 | 83800 | 285 | 169 | 172 | 337 | 205 | 205 | 0·61 (0·50-0·73) | 0·61 (0·51-0·73) |
| Bulungan | 133166 | 151800 | 154500 | 355 | 180 | 198 | 267 | 119 | 128 | 0·45 (0·37-0·53) | 0·48 (0·41-0·57) |
| Nunukan | 26607 | 25600 | 26400 | 424 | 186 | 254 | 1594 | 727 | 962 | 0·46 (0·39-0·54) | 0·60 (0·52-0·70) |
| Tana Tidung | 196918 | 199100 | 203200 | 47 | 17 | 24 | 24 | 9 | 12 | 0·36 (0·21-0·61) | 0·50 (0·31-0·80) |
| Kota Tarakan | 254262 | 242800 | 245700 | 677 | 473 | 518 | 266 | 195 | 211 | 0·73 (0·65-0·82) | 0·79 (0·71-0·89) |
| **North Sulawesi** | | | | | | | | | | | |
| Bolaang Mongondow | 247811 | 248751 | 250478 | 521 | 536 | 615 | 210 | 215 | 246 | 1·03 (0·91-1·16) | 1·17 (1·04-1·31) |
| Minahasa | 341176 | 347290 | 348673 | 716 | 451 | 677 | 210 | 130 | 194 | 0·62 (0·55-0·70) | 0·93 (0·83-1·03) |
| Kepulauan Sangihe | 131163 | 139262 | 139684 | 257 | 138 | 231 | 196 | 99 | 165 | 0·51 (0·41-0·62) | 0·84 (0·71-1·01) |
| Kepulauan Talaud | 92475 | 94521 | 94983 | 202 | 197 | 214 | 218 | 208 | 225 | 0·95 (0·78-1·16) | 1·03 (0·85-1·25) |
| Minahasa Selatan | 210695 | 236463 | 238746 | 505 | 259 | 290 | 240 | 110 | 121 | 0·46 (0·39-0·53) | 0·51 (0·44-0·58) |
| Minahasa Utara | 203624 | 224993 | 226915 | 512 | 450 | 431 | 251 | 200 | 190 | 0·80 (0·70-0·90) | 0·76 (0·66-0·86) |
| Bolaang Mongondow Utara | 80313 | 83112 | 83743 | 196 | 127 | 236 | 244 | 153 | 282 | 0·63 (0·50-0·78) | 1·16 (0·96-1·40) |
| Kep, Siau Tagulandang Biaro | 66403 | 71817 | 72135 | 165 | 119 | 110 | 248 | 166 | 152 | 0·67 (0·53-0·84) | 0·61 (0·48-0·78) |
| Minahasa Tenggara | 106899 | 116323 | 117079 | 193 | 163 | 245 | 181 | 140 | 209 | 0·78 (0·63-0·96) | 1·16 (0·96-1·40) |
| Bolaang Mongondow Selatan | 66071 | 69791 | 70529 | 228 | 118 | 130 | 345 | 169 | 184 | 0·49 (0·39-0·61) | 0·53 (0·43-0·66) |
| Bolaang Mongondow Timur | 72408 | 88241 | 89981 | 213 | 141 | 163 | 294 | 160 | 181 | 0·54 (0·44-0·67) | 0·62 (0·50-0·75) |

| District | Number of populations | | | Number of new TB reported cases | | | Case notification rate (CNR) per 100,000 populations | | | CNR ratio for 2020 vs 2019 (95% CI) | CNR ratio for 2021 vs 2019 (95% CI) |
|---|---|---|---|---|---|---|---|---|---|---|---|
| | 2019 | 2020 | 2021 | 2019 | 2020 | 2021 | 2019 | 2020 | 2021 | | |
| Kota Manado | 433635 | 451916 | 453182 | 2688 | 1471 | 1958 | 620 | 326 | 432 | 0·53 (0·49-0·56) | 0·70 (0·66-0·74) |
| Kota Bitung | 219004 | 225134 | 227177 | 759 | 543 | 604 | 347 | 241 | 266 | 0·70 (0·62-0·78) | 0·77 (0·69-0·85) |
| Kota Tomohon | 106917 | 100587 | 100853 | 347 | 229 | 366 | 325 | 228 | 363 | 0·70 (0·59-0·83) | 1·12 (0·97-1·29) |
| Kota Kotamobagu | 128387 | 123722 | 124473 | 350 | 256 | 439 | 273 | 207 | 353 | 0·76 (0·65-0·89) | 1·29 (1·12-1·49) |
| **Central Sulawesi** | | | | | | | | | | | |
| Banggai Kepulauan | 118401 | 120142 | 121680 | 263 | 159 | 195 | 222 | 132 | 160 | 0·60 (0·49-0·72) | 0·72 (0·60-0·87) |
| Banggai | 376808 | 362275 | 366220 | 1094 | 835 | 743 | 290 | 230 | 203 | 0·79 (0·73-0·87) | 0·70 (0·64-0·77) |
| Morowali | 121296 | 161727 | 167910 | 451 | 226 | 394 | 372 | 140 | 235 | 0·38 (0·32-0·44) | 0·63 (0·55-0·72) |
| Poso | 256393 | 244875 | 248350 | 277 | 253 | 263 | 108 | 103 | 106 | 0·96 (0·81-1·13) | 0·98 (0·83-1·16) |
| Donggala | 304110 | 300436 | 302910 | 539 | 269 | 306 | 177 | 90 | 101 | 0·51 (0·44-0·58) | 0·57 (0·50-0·65) |
| Toli-Toli | 235800 | 225154 | 226800 | 497 | 299 | 321 | 211 | 133 | 142 | 0·63 (0·55-0·73) | 0·67 (0·58-0·77) |
| Buol | 162179 | 145254 | 146630 | 226 | 153 | 224 | 139 | 105 | 153 | 0·76 (0·62-0·93) | 1·10 (0·91-1·32) |
| Parigi Moutong | 490915 | 440015 | 443170 | 677 | 630 | 505 | 138 | 143 | 114 | 1·04 (0·93-1·16) | 0·83 (0·74-0·93) |
| Tojo Una-Una | 153991 | 163829 | 166340 | 279 | 231 | 169 | 181 | 141 | 102 | 0·78 (0·65-0·93) | 0·56 (0·46-0·68) |
| Sigi | 239421 | 257585 | 261680 | 433 | 353 | 275 | 181 | 137 | 105 | 0·76 (0·66-0·87) | 0·58 (0·50-0·67) |
| Banggai Laut | 75003 | 70435 | 70870 | 119 | 147 | 167 | 159 | 209 | 236 | 1·32 (1·03-1·67) | 1·49 (1·18-1·88) |
| Morowali Utara | 128323 | 120789 | 122240 | 188 | 147 | 169 | 147 | 122 | 138 | 0·83 (0·67-1·03) | 0·94 (0·77-1·16) |
| Kota Palu | 391383 | 373218 | 377030 | 779 | 656 | 1030 | 199 | 176 | 273 | 0·88 (0·80-0·98) | 1·37 (1·25-1·51) |
| **South Sulawesi** | | | | | | | | | | | |
| Kepulauan Selayar | 135624 | 136871 | 136118 | 220 | 197 | 247 | 162 | 144 | 181 | 0·89 (0·73-1·08) | 1·12 (0·93-1·34) |
| Bulukumba | 420603 | 423012 | 421959 | 652 | 403 | 582 | 155 | 95 | 138 | 0·62 (0·54-0·69) | 0·89 (0·80-0·99) |
| Bantaeng | 187626 | 188495 | 189202 | 347 | 367 | 466 | 185 | 195 | 246 | 1·05 (0·91-1·22) | 1·33 (1·16-1·53) |
| Jeneponto | 363792 | 365610 | 367160 | 602 | 403 | 590 | 165 | 110 | 161 | 0·67 (0·59-0·76) | 0·97 (0·87-1·09) |
| Takalar | 298688 | 301424 | 298717 | 696 | 449 | 728 | 233 | 149 | 244 | 0·64 (0·57-0·72) | 1·05 (0·94-1·16) |

| District | Number of populations | | | Number of new TB reported cases | | | Case notification rate (CNR) per 100,000 populations | | | CNR ratio for 2020 vs 2019 (95% CI) | CNR ratio for 2021 vs 2019 (95% CI) |
|---|---|---|---|---|---|---|---|---|---|---|---|
| | 2019 | 2020 | 2021 | 2019 | 2020 | 2021 | 2019 | 2020 | 2021 | | |
| Gowa | 772684 | 784511 | 780138 | 1805 | 875 | 1223 | 234 | 112 | 157 | 0·48 (0·44-0·52) | 0·67 (0·62-0·72) |
| Sinjai | 244125 | 245389 | 245501 | 535 | 340 | 397 | 219 | 139 | 162 | 0·63 (0·55-0·72) | 0·74 (0·65-0·84) |
| Maros | 353121 | 356195 | 357320 | 673 | 433 | 542 | 191 | 122 | 152 | 0·64 (0·57-0·72) | 0·80 (0·71-0·89) |
| Pangkajene Dan Kepulauan | 335514 | 338219 | 339575 | 811 | 611 | 707 | 242 | 181 | 208 | 0·75 (0·67-0·83) | 0·86 (0·78-0·95) |
| Barru | 174323 | 174989 | 175023 | 265 | 200 | 263 | 152 | 114 | 150 | 0·75 (0·63-0·90) | 0·99 (0·83-1·17) |
| Bone | 758589 | 762073 | 757741 | 1282 | 851 | 1067 | 169 | 112 | 141 | 0·66 (0·61-0·72) | 0·83 (0·77-0·90) |
| Soppeng | 226991 | 227208 | 229539 | 379 | 249 | 212 | 167 | 110 | 92 | 0·66 (0·56-0·77) | 0·55 (0·47-0·65) |
| Wajo | 397814 | 398784 | 406091 | 877 | 575 | 773 | 220 | 144 | 190 | 0·65 (0·59-0·73) | 0·86 (0·78-0·95) |
| Sidenreng Rappang | 301972 | 304826 | 302918 | 584 | 375 | 520 | 193 | 123 | 172 | 0·64 (0·56-0·72) | 0·89 (0·79-1·00) |
| Pinrang | 377119 | 379402 | 381114 | 580 | 438 | 688 | 154 | 115 | 181 | 0·75 (0·66-0·85) | 1·17 (1·05-1·31) |
| Enrekang | 206387 | 207800 | 209974 | 207 | 159 | 203 | 100 | 77 | 97 | 0·76 (0·62-0·94) | 0·96 (0·79-1·17) |
| Luwu | 362027 | 364680 | 369924 | 622 | 415 | 496 | 172 | 114 | 134 | 0·66 (0·59-0·75) | 0·78 (0·69-0·88) |
| Tana Toraja | 234002 | 235103 | 239516 | 295 | 162 | 204 | 126 | 69 | 85 | 0·55 (0·45-0·66) | 0·68 (0·57-0·81) |
| Luwu Utara | 312883 | 315202 | 318064 | 609 | 328 | 383 | 195 | 104 | 120 | 0·54 (0·47-0·61) | 0·62 (0·55-0·70) |
| Luwu Timur | 299673 | 305407 | 303479 | 444 | 345 | 449 | 148 | 113 | 148 | 0·76 (0·66-0·88) | 1·00 (0·88-1·14) |
| Toraja Utara | 231214 | 232394 | 237259 | 230 | 222 | 270 | 99 | 96 | 114 | 0·96 (0·80-1·15) | 1·14 (0·96-1·36) |
| Kota Makassar | 1526677 | 1545373 | 1555088 | 6731 | 4849 | 5934 | 441 | 314 | 382 | 0·71 (0·69-0·74) | 0·87 (0·84-0·90) |
| Kota Parepare | 145178 | 146714 | 147090 | 460 | 427 | 399 | 317 | 291 | 271 | 0·92 (0·81-1·05) | 0·86 (0·75-0·98) |
| Kota Palopo | 184614 | 188323 | 187671 | 454 | 387 | 606 | 246 | 205 | 323 | 0·84 (0·73-0·96) | 1·31 (1·16-1·48) |
| **Southeast Sulawesi** | | | | | | | | | | | |
| Buton | 102641 | 103869 | 117040 | 239 | 155 | 194 | 233 | 149 | 166 | 0·64 (0·52-0·78) | 0·71 (0·59-0·86) |
| Muna | 224099 | 227289 | 218956 | 406 | 257 | 285 | 181 | 113 | 130 | 0·62 (0·53-0·73) | 0·72 (0·62-0·84) |
| Konawe | 254695 | 260411 | 261116 | 362 | 169 | 437 | 142 | 65 | 167 | 0·46 (0·38-0·55) | 1·18 (1·02-1·35) |
| Kolaka | 261664 | 266069 | 241366 | 377 | 320 | 379 | 144 | 120 | 157 | 0·84 (0·72-0·97) | 1·09 (0·95-1·26) |

| District | Number of populations | | | Number of new TB reported cases | | | Case notification rate (CNR) per 100,000 populations | | | CNR ratio for 2020 vs 2019 (95% CI) | CNR ratio for 2021 vs 2019 (95% CI) |
|---|---|---|---|---|---|---|---|---|---|---|---|
| | 2019 | 2020 | 2021 | 2019 | 2020 | 2021 | 2019 | 2020 | 2021 | | |
| Konawe Selatan | 314785 | 319291 | 312674 | 475 | 330 | 357 | 151 | 103 | 114 | 0·69 (0·60-0·79) | 0·76 (0·66-0·87) |
| Bombana | 184570 | 189269 | 151910 | 451 | 574 | 412 | 244 | 303 | 271 | 1·24 (1·10-1·40) | 1·11 (0·97-1·27) |
| Wakatobi | 95892 | 96111 | 113122 | 131 | 46 | 90 | 137 | 48 | 80 | 0·35 (0·25-0·48) | 0·58 (0·45-0·76) |
| Kolaka Utara | 150831 | 153669 | 139234 | 165 | 155 | 176 | 109 | 101 | 126 | 0·92 (0·74-1·15) | 1·16 (0·93-1·43) |
| Buton Utara | 64072 | 64993 | 67714 | 81 | 40 | 39 | 126 | 62 | 58 | 0·49 (0·34-0·71) | 0·46 (0·31-0·66) |
| Konawe Utara | 63814 | 65183 | 68950 | 140 | 100 | 128 | 219 | 153 | 186 | 0·70 (0·54-0·90) | 0·85 (0·67-1·08) |
| Kolaka Timur | 133324 | 135569 | 120966 | 173 | 118 | 114 | 130 | 87 | 94 | 0·67 (0·53-0·85) | 0·73 (0·57-0·92) |
| Konawe Kepulauan | 34219 | 34666 | 37639 | 45 | 71 | 62 | 132 | 205 | 165 | 1·56 (1·08-2·26) | 1·25 (0·85-1·84) |
| Muna Barat | 81624 | 82785 | 84777 | 140 | 88 | 112 | 172 | 106 | 132 | 0·62 (0·48-0·81) | 0·77 (0·60-0·99) |
| Buton Tengah | 93091 | 94207 | 116599 | 194 | 159 | 198 | 208 | 169 | 170 | 0·81 (0·66-1·00) | 0·82 (0·67-0·99) |
| Buton Selatan | 80784 | 81752 | 95472 | 101 | 83 | 74 | 125 | 102 | 78 | 0·81 (0·61-1·09) | 0·62 (0·46-0·83) |
| Kota Kendari | 392830 | 404232 | 350267 | 712 | 510 | 1177 | 181 | 126 | 336 | 0·70 (0·62-0·78) | 1·85 (1·69-2·03) |
| Kota Baubau | 171802 | 176224 | 161354 | 332 | 342 | 523 | 193 | 194 | 324 | 1·00 (0·86-1·17) | 1·68 (1·46-1·92) |
| **Gorontalo** | | | | | | | | | | | |
| Boalemo | 167024 | 145868 | 147038 | 377 | 283 | 363 | 226 | 194 | 247 | 0·86 (0·74-1·00) | 1·09 (0·95-1·26) |
| Gorontalo | 378527 | 393107 | 395635 | 1417 | 631 | 1305 | 374 | 161 | 330 | 0·43 (0·39-0·47) | 0·88 (0·82-0·95) |
| Pohuwato | 161373 | 146432 | 147689 | 446 | 312 | 309 | 276 | 213 | 209 | 0·77 (0·67-0·89) | 0·76 (0·66-0·87) |
| Bone Bolango | 161236 | 162778 | 164277 | 633 | 495 | 753 | 393 | 304 | 458 | 0·78 (0·69-0·87) | 1·17 (1·05-1·30) |
| Gorontalo Utara | 115072 | 124957 | 126521 | 349 | 262 | 303 | 303 | 210 | 239 | 0·69 (0·59-0·81) | 0·79 (0·68-0·92) |
| Kota Gorontalo | 219399 | 198539 | 199788 | 798 | 426 | 799 | 364 | 215 | 400 | 0·59 (0·53-0·66) | 1·10 (1·00-1·21) |
| **West Sulawesi** | | | | | | | | | | | |
| Majene | 173884 | 174407 | 175790 | 512 | 439 | 482 | 294 | 252 | 274 | 0·86 (0·75-0·97) | 0·93 (0·82-1·05) |
| Polewali Mandar | 442576 | 478534 | 483920 | 913 | 655 | 868 | 206 | 137 | 179 | 0·66 (0·60-0·73) | 0·87 (0·79-0·95) |
| Mamasa | 161971 | 163383 | 164800 | 169 | 101 | 113 | 104 | 62 | 69 | 0·59 (0·46-0·76) | 0·66 (0·52-0·83) |

| District | Number of populations | | | Number of new TB reported cases | | | Case notification rate (CNR) per 100,000 populations | | | CNR ratio for 2020 vs 2019 (95% CI) | CNR ratio for 2021 vs 2019 (95% CI) |
|---|---|---|---|---|---|---|---|---|---|---|---|
| | 2019 | 2020 | 2021 | 2019 | 2020 | 2021 | 2019 | 2020 | 2021 | | |
| Mamuju | 293326 | 278764 | 281850 | 647 | 459 | 521 | 221 | 165 | 185 | 0·75 (0·66-0·84) | 0·84 (0·75-0·94) |
| Pasangkayu | 174471 | 188861 | 193100 | 237 | 174 | 176 | 136 | 92 | 91 | 0·68 (0·56-0·82) | 0·67 (0·55-0·81) |
| Mamuju Tengah | 134028 | 135280 | 137380 | 258 | 138 | 222 | 192 | 1020 | 162 | 5·30 (4·41-6·37) | 0·84 (0·70-1·00) |
| **Maluku** | | | | | | | | | | | |
| Maluku Tenggara Barat | 113012 | 123572 | 124075 | 350 | 85 | 222 | 310 | 69 | 179 | 0·22 (0·18-0·28) | 0·58 (0·49-0·68) |
| Maluku Tenggara | 99790 | 121511 | 122640 | 382 | 249 | 326 | 383 | 205 | 266 | 0·54 (0·46-0·63) | 0·69 (0·60-0·80) |
| Maluku Tengah | 373378 | 423094 | 424730 | 747 | 285 | 644 | 200 | 67 | 152 | 0·34 (0·30-0·38) | 0·76 (0·68-0·84) |
| Buru | 143688 | 135238 | 136393 | 140 | 121 | 154 | 97 | 89 | 113 | 0·92 (0·72-1·17) | 1·16 (0·92-1·46) |
| Kepulauan Aru | 96114 | 102237 | 102916 | 421 | 134 | 220 | 438 | 131 | 214 | 0·30 (0·25-0·36) | 0·49 (0·42-0·57) |
| Seram Bagian Barat | 171586 | 212393 | 214733 | 206 | 191 | 206 | 120 | 90 | 96 | 0·75 (0·62-0·91) | 0·80 (0·66-0·97) |
| Seram Bagian Timur | 114677 | 137972 | 140271 | 156 | 87 | 208 | 136 | 63 | 148 | 0·46 (0·36-0·60) | 1·09 (0·89-1·34) |
| Maluku Barat Daya | 73103 | 81928 | 82187 | 122 | 127 | 127 | 167 | 155 | 155 | 0·93 (0·72-1·19) | 0·93 (0·72-1·19) |
| Buru Selatan | 63328 | 75410 | 76715 | 81 | 27 | 70 | 128 | 36 | 91 | 0·28 (0·19-0·42) | 0·71 (0·52-0·98) |
| Kota Ambon | 478616 | 347288 | 347644 | 1556 | 773 | 1663 | 325 | 223 | 478 | 0·69 (0·63-0·75) | 1·47 (1·37-1·58) |
| Kota Tual | 75578 | 88280 | 90322 | 216 | 196 | 176 | 286 | 222 | 195 | 0·78 (0·64-0·94) | 0·68 (0·56-0·83) |
| **North Maluku** | | | | | | | | | | | |
| Halmahera Barat | 118287 | 132349 | 134630 | 201 | 185 | 186 | 170 | 140 | 138 | 0·82 (0·67-1·00) | 0·81 (0·67-0·99) |
| Halmahera Tengah | 55728 | 56802 | 57809 | 81 | 42 | 99 | 145 | 74 | 171 | 0·51 (0·35-0·73) | 1·18 (0·88-1·58) |
| Kepulauan Sula | 102886 | 104082 | 105293 | 111 | 89 | 118 | 108 | 86 | 112 | 0·79 (0·60-1·05) | 1·04 (0·80-1·35) |
| Halmahera Selatan | 235090 | 248395 | 251690 | 419 | 290 | 339 | 178 | 117 | 135 | 0·66 (0·56-0·76) | 0·76 (0·66-0·87) |
| Halmahera Utara | 193851 | 197638 | 199936 | 368 | 235 | 388 | 190 | 119 | 194 | 0·63 (0·53-0·74) | 1·02 (0·89-1·18) |
| Halmahera Timur | 95005 | 91707 | 92954 | 137 | 62 | 85 | 144 | 68 | 91 | 0·47 (0·35-0·63) | 0·63 (0·48-0·83) |
| Pulau Morotai | 67284 | 74436 | 76102 | 89 | 71 | 166 | 132 | 95 | 218 | 0·72 (0·53-0·98) | 1·65 (1·28-2·13) |
| Pulau Taliabu | 53018 | 58047 | 58744 | 37 | 23 | 35 | 70 | 40 | 60 | 0·57 (0·34-0·95) | 0·85 (0·54-1·35) |

| District | Number of populations | | | Number of new TB reported cases | | | Case notification rate (CNR) per 100,000 populations | | | CNR ratio for 2020 vs 2019 (95% CI) | CNR ratio for 2021 vs 2019 (95% CI) |
|---|---|---|---|---|---|---|---|---|---|---|---|
| | 2019 | 2020 | 2021 | 2019 | 2020 | 2021 | 2019 | 2020 | 2021 | | |
| Kota Ternate | 233208 | 205001 | 205870 | 530 | 450 | 674 | 227 | 220 | 327 | 0·97 (0·85-1·10) | 1·44 (1·29-1·61) |
| Kota Tidore Kepulauan | 101414 | 114480 | 116149 | 249 | 149 | 221 | 246 | 130 | 190 | 0·53 (0·43-0·65) | 0·78 (0·65-0·93) |
| **West Papua** | | | | | | | | | | | |
| Fakfak | 78686 | 85197 | 85817 | 227 | 158 | 253 | 288 | 185 | 295 | 0·64 (0·53-0·79) | 1·02 (0·85-1·22) |
| Kaimana | 60216 | 62256 | 62957 | 211 | 111 | 144 | 350 | 178 | 229 | 0·51 (0·41-0·64) | 0·65 (0·53-0·81) |
| Teluk Wondama | 32521 | 41644 | 42609 | 109 | 71 | 125 | 335 | 170 | 293 | 0·51 (0·38-0·68) | 0·88 (0·68-1·13) |
| Teluk Bintuni | 64406 | 87083 | 89418 | 302 | 202 | 276 | 469 | 232 | 309 | 0·50 (0·42-0·59) | 0·66 (0·56-0·77) |
| Manokwari | 175178 | 192663 | 194905 | 758 | 519 | 699 | 433 | 269 | 359 | 0·62 (0·56-0·70) | 0·83 (0·75-0·92) |
| Sorong Selatan | 46922 | 52469 | 53167 | 196 | 45 | 82 | 418 | 86 | 154 | 0·21 (0·15-0·28) | 0·37 (0·29-0·47) |
| Sorong | 88927 | 118679 | 121963 | 434 | 142 | 269 | 488 | 120 | 221 | 0·25 (0·21-0·29) | 0·45 (0·39-0·52) |
| Raja Ampat | 48493 | 64141 | 65403 | 110 | 67 | 53 | 227 | 104 | 81 | 0·46 (0·34-0·62) | 0·36 (0·26-0·49) |
| Tambrauw | 13879 | 28379 | 31385 | 0 | 6 | 2 | 0 | 21 | 6 | NA | NA |
| Maybrat | 40899 | 42991 | 43364 | 3 | 0 | 0 | 7 | 0 | 0 | NA | NA |
| Manokwari Selatan | 24220 | 35949 | 37149 | 25 | 39 | 51 | 103 | 108 | 137 | 1·05 (0·64-1·74) | 1·33 (0·83-2·14) |
| Pegunungan Arfak | 30976 | 38207 | 38936 | 0 | 5 | 1 | 0 | 13 | 3 | NA | NA |
| Kota Sorong | 254294 | 284410 | 289767 | 592 | 334 | 474 | 233 | 117 | 164 | 0·50 (0·44-0·58) | 0·70 (0·62-0·79) |
| **Papua** | | | | | | | | | | | |
| Merauke | 227411 | 230932 | 231696 | 1010 | 937 | 818 | 444 | 406 | 353 | 0·91 (0·84-1·00) | 0·80 (0·73-0·87) |
| Jayawijaya | 217887 | 269553 | 273291 | 386 | 199 | 406 | 177 | 74 | 149 | 0·42 (0·35-0·49) | 0·84 (0·73-0·96) |
| Jayapura | 131802 | 166171 | 168476 | 844 | 633 | 950 | 640 | 381 | 564 | 0·60 (0·54-0·66) | 0·88 (0·80-0·97) |
| Nabire | 150308 | 169136 | 170914 | 1285 | 991 | 1133 | 855 | 586 | 663 | 0·69 (0·63-0·74) | 0·78 (0·72-0·84) |
| Kepulauan Yapen | 101204 | 112676 | 114210 | 549 | 236 | 280 | 542 | 209 | 245 | 0·39 (0·33-0·45) | 0·45 (0·39-0·52) |
| Biak Numfor | 152401 | 134650 | 135231 | 789 | 497 | 541 | 518 | 369 | 400 | 0·71 (0·64-0·80) | 0·77 (0·69-0·86) |
| Paniai | 177410 | 220410 | 223467 | 443 | 383 | 378 | 250 | 174 | 169 | 0·70 (0·61-0·80) | 0·68 (0·59-0·78) |

| District | Number of populations | | | Number of new TB reported cases | | | Case notification rate (CNR) per 100,000 populations | | | CNR ratio for 2020 vs 2019 (95% CI) | CNR ratio for 2021 vs 2019 (95% CI) |
|---|---|---|---|---|---|---|---|---|---|---|---|
| | 2019 | 2020 | 2021 | 2019 | 2020 | 2021 | 2019 | 2020 | 2021 | | |
| Puncak Jaya | 129300 | 224527 | 227641 | 46 | 50 | 18 | 36 | 22 | 8 | 0·63 (0·42-0·93) | 0·22 (0·14-0·37) |
| Mimika | 219689 | 311969 | 316295 | 1792 | 1524 | 1910 | 816 | 489 | 604 | 0·60 (0·56-0·64) | 0·74 (0·69-0·79) |
| Boven Digoel | 69211 | 64285 | 64716 | 360 | 302 | 343 | 520 | 470 | 530 | 0·90 (0·78-1·05) | 1·02 (0·88-1·18) |
| Mappi | 103292 | 108295 | 109579 | 1067 | 1086 | 1158 | 1033 | 1003 | 1057 | 0·97 (0·89-1·06) | 1·02 (0·94-1·11) |
| Asmat | 97490 | 110105 | 111632 | 399 | 384 | 275 | 409 | 349 | 246 | 0·85 (0·74-0·98) | 0·60 (0·52-0·70) |
| Yahukimo | 190887 | 350880 | 355746 | 97 | 176 | 192 | 51 | 50 | 54 | 0·99 (0·77-1·26) | 1·06 (0·83-1·36) |
| Pegunungan Bintang | 75788 | 77873 | 78178 | 62 | 60 | 68 | 82 | 77 | 87 | 0·94 (0·66-1·34) | 1·06 (0·75-1·50) |
| Tolikara | 139111 | 236986 | 240272 | 37 | 0 | 1 | 27 | 0 | 0 | NA | 0·02 (0·01-0·04) |
| Sarmi | 40515 | 41415 | 41849 | 103 | 56 | 40 | 254 | 135 | 96 | 0·53 (0·39-0·73) | 0·38 (0·26-0·53) |
| Keerom | 57100 | 61623 | 62157 | 108 | 145 | 120 | 189 | 235 | 193 | 1·24 (0·97-1·60) | 1·02 (0·79-1·32) |
| Waropen | 31514 | 33943 | 34414 | 64 | 29 | 27 | 203 | 85 | 78 | 0·42 (0·27-0·64) | 0·39 (0·25-0·60) |
| Supiori | 20710 | 22547 | 22860 | 65 | 26 | 71 | 314 | 115 | 311 | 0·37 (0·24-0·57) | 0·99 (0·71-1·39) |
| Mamberamo Raya | 24086 | 36483 | 36989 | 41 | 23 | 40 | 170 | 63 | 108 | 0·37 (0·23-0·60) | 0·64 (0·41-0·98) |
| Nduga | 98595 | 106533 | 107921 | 0 | 0 | 0 | 0 | 0 | 0 | NA | NA |
| Lanny Jaya | 178995 | 196339 | 198686 | 12 | 12 | 12 | 7 | 6 | 6 | 0·91 (0·41-2·03) | 0·90 (0·40-2·00) |
| Mamberamo Tengah | 48201 | 50685 | 51160 | 0 | 2 | 2 | 0 | 4 | 4 | NA | NA |
| Yalimo | 62605 | 101973 | 103387 | 19 | 6 | 6 | 30 | 6 | 6 | 0·19 (0·09-0·44) | 0·19 (0·08-0·43) |
| Puncak | 113204 | 114741 | 115474 | 16 | 15 | 6 | 14 | 13 | 5 | 0·93 (0·46-1·87) | 0·37 (0·15-0·90) |
| Dogiyai | 97902 | 116206 | 117818 | 18 | 10 | 17 | 18 | 9 | 14 | 0·47 (0·22-1·00) | 0·79 (0·41-1·52) |
| Intan Jaya | 49293 | 135043 | 136916 | 14 | 0 | 0 | 28 | 0 | 0 | NA | NA |
| Deiyai | 73199 | 99091 | 100466 | 0 | 0 | 0 | 0 | 0 | 0 | NA | NA |
| Kota Jayapura | 300192 | 398478 | 404004 | 2137 | 1536 | 2203 | 712 | 385 | 545 | 0·54 (0·51-0·58) | 0·77 (0·72-0·81) |

**Supplementary table 2. Number of annual populations, reported new TB reported cases, case notification rate (CNR), and CNR ratio in 2020 and 2021 (pandemic), compared to in 2019 (pre-pandemic)**

| District | Number of populations | | | Number of reported mortalities | | | Mortality rate per 100,000 population | | | Mortality rate ratio for 2020 vs 2019 (95% CI) | Mortality rate ratio for 2021 vs 2019 (95% CI) |
|---|---|---|---|---|---|---|---|---|---|---|---|
| | 2019 | 2020 | 2021 | 2019 | 2020 | 2021 | 2019 | 2020 | 2021 | | |
| **Aceh** | | | | | | | | | | | |
| Simeulue | 93228 | 92865 | 93760 | 1 | 2 | 3 | 1 | 2 | 3 | 2·01 (0·19 - 21·11) | 2·98 (0·35 - 25·72) |
| Aceh Singkil | 124101 | 126514 | 128380 | 5 | 7 | 0 | 4 | 6 | 0 | 1·37 (0·44 - 4·31) | NA |
| Aceh Selatan | 238081 | 232414 | 234630 | 14 | 13 | 13 | 6 | 6 | 6 | 0·95 (0·45 - 2·02) | 0·94 (0·44 - 2) |
| Aceh Tenggara | 216495 | 220860 | 224120 | 1 | 0 | 2 | 0 | 0 | 1 | NA | 1·93 (0·18 - 20·41) |
| Aceh Timur | 436081 | 422401 | 427030 | 27 | 17 | 18 | 6 | 4 | 4 | 0·65 (0·36 - 1·19) | 0·68 (0·38 - 1·23) |
| Aceh Tengah | 212494 | 215576 | 218680 | 0 | 20 | 5 | 0 | 9 | 2 | NA | NA |
| Aceh Barat | 210113 | 198736 | 200580 | 6 | 8 | 7 | 3 | 4 | 3 | 1·41 (0·49 - 4·04) | 1·22 (0·41 - 3·63) |
| Aceh Besar | 425216 | 405535 | 409530 | 14 | 22 | 14 | 3 | 5 | 3 | 1·65 (0·85 - 3·2) | 1·04 (0·5 - 2·18) |
| Pidie | 444976 | 435275 | 439400 | 11 | 21 | 18 | 2 | 5 | 4 | 1·95 (0·95 - 3·99) | 1·66 (0·79 - 3·48) |
| Bireuen | 471635 | 436418 | 439790 | 9 | 47 | 29 | 2 | 11 | 7 | 5·64 (3 - 10·61) | 3·46 (1·71 - 6·97) |
| Aceh Utara | 619407 | 602793 | 608110 | 7 | 21 | 18 | 1 | 3 | 3 | 3·08 (1·37 - 6·94) | 2·62 (1·13 - 6·07) |
| Aceh Barat Daya | 150393 | 150775 | 152660 | 14 | 19 | 5 | 9 | 13 | 3 | 1·35 (0·68 - 2·69) | 0·35 (0·13 - 0·93) |
| Gayo Lues | 94100 | 99532 | 101100 | 0 | 0 | 3 | 0 | 0 | 3 | NA | NA |
| Aceh Tamiang | 295011 | 294356 | 297520 | 12 | 12 | 21 | 4 | 4 | 7 | 1 (0·45 - 2·23) | 1·74 (0·86 - 3·5) |
| Nagan Raya | 167294 | 168392 | 170590 | 7 | 3 | 3 | 4 | 2 | 2 | 0·43 (0·11 - 1·58) | 0·42 (0·11 - 1·56) |
| Aceh Jaya | 92892 | 93159 | 94420 | 2 | 4 | 1 | 2 | 4 | 1 | 1·99 (0·38 - 10·53) | 0·49 (0·05 - 5·16) |
| Bener Meriah | 148175 | 161342 | 164520 | 1 | 4 | 5 | 1 | 2 | 3 | 3·67 (0·48 - 28·37) | 4·5 (0·64 - 31·88) |
| Pidie Jaya | 161215 | 158397 | 160330 | 9 | 1 | 1 | 6 | 1 | 1 | 0·11 (0·02 - 0·62) | 0·11 (0·02 - 0·61) |
| Kota Banda Aceh | 270321 | 252899 | 255030 | 0 | 14 | 19 | 0 | 6 | 7 | NA | NA |
| Kota Sabang | 34874 | 41197 | 42070 | 6 | 1 | 0 | 17 | 2 | 0 | 0·14 (0·02 - 0·86) | NA |
| Kota Langsa | 176811 | 185971 | 188880 | 11 | 7 | 10 | 6 | 4 | 5 | 0·61 (0·24 - 1·55) | 0·85 (0·36 - 2) |
| Kota Lhokseumawe | 207202 | 188713 | 189940 | 1 | 5 | 9 | 0 | 3 | 5 | 5·49 (0·81 - 37) | 9·82 (1·84 - 52·52) |
| Kota Subulussalam | 81417 | 90751 | 92670 | 1 | 1 | 1 | 1 | 1 | 1 | 0·9 (0·06 - 14·32) | 0·88 (0·06 - 14·02) |

| District | Number of populations | | | Number of reported mortalities | | | Mortality rate per 100,000 population | | | Mortality rate ratio for 2020 vs 2019 (95% CI) | Mortality rate ratio for 2021 vs 2019 (95% CI) |
|---|---|---|---|---|---|---|---|---|---|---|---|
| | 2019 | 2020 | 2021 | 2019 | 2020 | 2021 | 2019 | 2020 | 2021 | | |
| **North Sumatera** | | | | | | | | | | | |
| Nias | 143319 | 143983 | 147794 | 0 | 1 | 10 | 0 | 1 | 7 | NA | NA |
| Mandailing Natal | 447287 | 451028 | 478062 | 22 | 18 | 19 | 5 | 4 | 4 | 0·81 (0·44 - 1·51) | 0·81 (0·44 - 1·49) |
| Tapanuli Selatan | 281931 | 283389 | 303685 | 17 | 20 | 24 | 6 | 7 | 8 | 1·17 (0·61 - 2·23) | 1·31 (0·71 - 2·43) |
| Tapanuli Tengah | 376667 | 382917 | 369300 | 0 | 16 | 26 | 0 | 4 | 7 | NA | NA |
| Tapanuli Utara | 301789 | 303688 | 315222 | 9 | 16 | 15 | 3 | 5 | 5 | 1·77 (0·79 - 3·95) | 1·6 (0·7 - 3·62) |
| Toba Samosir | 183712 | 184493 | 208754 | 18 | 9 | 8 | 10 | 5 | 4 | 0·5 (0·23 - 1·09) | 0·39 (0·18 - 0·87) |
| Labuhan Batu | 494178 | 501596 | 499982 | 25 | 31 | 33 | 5 | 6 | 7 | 1·22 (0·72 - 2·07) | 1·3 (0·78 - 2·19) |
| Asahan | 729795 | 735026 | 777626 | 30 | 26 | 19 | 4 | 4 | 2 | 0·86 (0·51 - 1·45) | 0·59 (0·34 - 1·05) |
| Simalungun | 867922 | 871678 | 1003727 | 49 | 76 | 43 | 6 | 9 | 4 | 1·54 (1·08 - 2·21) | 0·76 (0·5 - 1·14) |
| Dairi | 284304 | 285481 | 311665 | 4 | 25 | 23 | 1 | 9 | 7 | 6·22 (2·48 - 15·64) | 5·25 (2·03 - 13·55) |
| Karo | 415878 | 421997 | 409077 | 17 | 13 | 12 | 4 | 3 | 3 | 0·75 (0·37 - 1·55) | 0·72 (0·34 - 1·5) |
| Deli Serdang | 2195709 | 2234320 | 1941374 | 99 | 105 | 109 | 5 | 5 | 6 | 1·04 (0·79 - 1·37) | 1·25 (0·95 - 1·63) |
| Langkat | 1041775 | 1048100 | 1034519 | 3 | 48 | 22 | 0 | 5 | 2 | 15·9 (6·71 - 37·71) | 7·38 (2·65 - 20·61) |
| Nias Selatan | 319902 | 322520 | 366163 | 11 | 4 | 6 | 3 | 1 | 2 | 0·36 (0·12 - 1·08) | 0·48 (0·18 - 1·26) |
| Humbang Hasundutan | 190186 | 191776 | 199719 | 0 | 14 | 11 | 0 | 7 | 6 | NA | NA |
| Pakpak Bharat | 48935 | 49688 | 53315 | 4 | 4 | 1 | 8 | 8 | 2 | 0·98 (0·25 - 3·94) | 0·23 (0·03 - 1·7) |
| Samosir | 126188 | 126710 | 137696 | 15 | 14 | 10 | 12 | 11 | 7 | 0·93 (0·45 - 1·93) | 0·61 (0·28 - 1·35) |
| Serdang Bedagai | 616396 | 617772 | 662076 | 25 | 20 | 25 | 4 | 3 | 4 | 0·8 (0·44 - 1·44) | 0·93 (0·53 - 1·62) |
| Batu Bara | 416493 | 420103 | 413171 | 18 | 43 | 15 | 4 | 10 | 4 | 2·37 (1·39 - 4·04) | 0·84 (0·42 - 1·67) |
| Padang Lawas Utara | 272713 | 277423 | 263551 | 18 | 13 | 7 | 7 | 5 | 3 | 0·71 (0·35 - 1·44) | 0·4 (0·17 - 0·94) |
| Padang Lawas | 281239 | 286627 | 263719 | 23 | 9 | 13 | 8 | 3 | 5 | 0·38 (0·18 - 0·81) | 0·6 (0·31 - 1·18) |
| Labuhan Batu Selatan | 338982 | 344819 | 316798 | 7 | 18 | 8 | 2 | 5 | 3 | 2·53 (1·09 - 5·87) | 1·22 (0·44 - 3·37) |
| Labuhan Batu Utara | 363816 | 366603 | 385869 | 34 | 28 | 25 | 9 | 8 | 6 | 0·82 (0·5 - 1·35) | 0·69 (0·41 - 1·16) |
| Nias Utara | 137967 | 138800 | 148790 | 10 | 3 | 5 | 7 | 2 | 3 | 0·3 (0·09 - 1) | 0·46 (0·16 - 1·32) |

| District | Number of populations | | | Number of reported mortalities | | | Mortality rate per 100,000 population | | | Mortality rate ratio for 2020 vs 2019 (95% CI) | Mortality rate ratio for 2021 vs 2019 (95% CI) |
|---|---|---|---|---|---|---|---|---|---|---|---|
| | 2019 | 2020 | 2021 | 2019 | 2020 | 2021 | 2019 | 2020 | 2021 | | |
| Nias Barat | 82154 | 82425 | 90585 | 5 | 2 | 2 | 6 | 2 | 2 | 0·4 (0·08 - 1·94) | 0·36 (0·08 - 1·75) |
| Kota Sibolga | 87626 | 87791 | 89932 | 2 | 9 | 10 | 2 | 10 | 11 | 4·49 (1·11 - 18·16) | 4·87 (1·24 - 19·18) |
| Kota Tanjung Balai | 175223 | 177005 | 177640 | 24 | 11 | 17 | 14 | 6 | 10 | 0·45 (0·23 - 0·91) | 0·7 (0·38 - 1·3) |
| Kota Pematang Siantar | 255317 | 257110 | 270768 | 0 | 4 | 6 | 0 | 2 | 2 | NA | NA |
| Kota Tebing Tinggi | 164402 | 166100 | 174969 | 5 | 6 | 10 | 3 | 4 | 6 | 1·19 (0·36 - 3·89) | 1·88 (0·65 - 5·4) |
| Kota Medan | 2279894 | 2295003 | 2460858 | 112 | 124 | 132 | 5 | 5 | 5 | 1·1 (0·85 - 1·42) | 1·09 (0·85 - 1·4) |
| Kota Binjai | 276597 | 279302 | 295361 | 10 | 12 | 15 | 4 | 4 | 5 | 1·19 (0·51 - 2·75) | 1·4 (0·63 - 3·11) |
| Kota Padangsidimpuan | 221827 | 224483 | 227674 | 4 | 12 | 10 | 2 | 5 | 4 | 2·96 (1·01 - 8·71) | 2·44 (0·79 - 7·48) |
| Kota Gunungsitoli | 142426 | 143776 | 136707 | 15 | 10 | 2 | 11 | 7 | 1 | 0·66 (0·3 - 1·46) | 0·14 (0·04 - 0·49) |
| **West Sumatera** | | | | | | | | | | | |
| Kepulauan Mentawai | 92021 | 87623 | 88389 | 0 | 0 | 2 | 0 | 0 | 2 | NA | NA |
| Pesisir Selatan | 463923 | 504418 | 509618 | 34 | 30 | 34 | 7 | 6 | 7 | 0·81 (0·5 - 1·32) | 0·91 (0·57 - 1·46) |
| Solok | 373414 | 391497 | 394237 | 4 | 2 | 6 | 1 | 1 | 2 | 0·48 (0·09 - 2·51) | 1·42 (0·4 - 5) |
| Sijunjung | 237376 | 235045 | 237313 | 13 | 2 | 6 | 5 | 1 | 3 | 0·16 (0·04 - 0·57) | 0·46 (0·18 - 1·19) |
| Tanah Datar | 348219 | 371704 | 373693 | 5 | 28 | 19 | 1 | 8 | 5 | 5·25 (2·24 - 12·28) | 3·54 (1·41 - 8·91) |
| Padang Pariaman | 415613 | 430626 | 433018 | 38 | 45 | 40 | 9 | 10 | 9 | 1·14 (0·74 - 1·76) | 1·01 (0·65 - 1·58) |
| Agam | 491282 | 529138 | 534202 | 19 | 26 | 13 | 4 | 5 | 2 | 1·27 (0·7 - 2·29) | 0·63 (0·31 - 1·27) |
| Lima Puluh Kota | 382817 | 383525 | 385634 | 40 | 19 | 15 | 10 | 5 | 4 | 0·47 (0·28 - 0·81) | 0·37 (0·21 - 0·66) |
| Pasaman | 281211 | 299851 | 303103 | 22 | 23 | 20 | 8 | 8 | 7 | 0·98 (0·55 - 1·76) | 0·84 (0·46 - 1·54) |
| Solok Selatan | 171075 | 182027 | 184854 | 6 | 2 | 2 | 4 | 1 | 1 | 0·31 (0·07 - 1·42) | 0·31 (0·07 - 1·4) |
| Dharmasraya | 247579 | 228591 | 231217 | 3 | 2 | 4 | 1 | 1 | 2 | 0·72 (0·12 - 4·29) | 1·43 (0·32 - 6·33) |
| Pasaman Barat | 443722 | 431672 | 436313 | 17 | 17 | 26 | 4 | 4 | 6 | 1·03 (0·52 - 2·01) | 1·56 (0·85 - 2·85) |
| Kota Padang | 950871 | 909040 | 913448 | 78 | 91 | 48 | 8 | 10 | 5 | 1·22 (0·9 - 1·65) | 0·64 (0·45 - 0·92) |
| Kota Solok | 71010 | 73438 | 74469 | 2 | 6 | 6 | 3 | 8 | 8 | 2·9 (0·63 - 13·36) | 2·86 (0·62 - 13·2) |
| Kota Sawah Lunto | 62524 | 65138 | 65687 | 3 | 1 | 2 | 5 | 2 | 3 | 0·32 (0·04 - 2·73) | 0·63 (0·11 - 3·74) |

| District | Number of populations | | | Number of reported mortalities | | | Mortality rate per 100,000 population | | | Mortality rate ratio for 2020 vs 2019 (95% CI) | Mortality rate ratio for 2021 vs 2019 (95% CI) |
|---|---|---|---|---|---|---|---|---|---|---|---|
| | 2019 | 2020 | 2021 | 2019 | 2020 | 2021 | 2019 | 2020 | 2021 | | |
| Kota Padang Panjang | 53693 | 56311 | 56971 | 6 | 2 | 7 | 11 | 4 | 12 | 0·32 (0·07 - 1·45) | 1·1 (0·37 - 3·27) |
| Kota Bukittinggi | 130773 | 121028 | 121588 | 7 | 5 | 2 | 5 | 4 | 2 | 0·77 (0·25 - 2·42) | 0·31 (0·07 - 1·36) |
| Kota Payakumbuh | 135573 | 139576 | 141184 | 6 | 3 | 6 | 4 | 2 | 4 | 0·49 (0·13 - 1·89) | 0·96 (0·31 - 2·98) |
| Kota Pariaman | 88501 | 94224 | 95294 | 2 | 7 | 8 | 2 | 7 | 8 | 3·29 (0·75 - 14·48) | 3·71 (0·88 - 15·74) |
| **Riau** | | | | | | | | | | | |
| Kuantan Singingi | 327316 | 334943 | 339894 | 16 | 1 | 20 | 5 | 0 | 6 | 0·06 (0·01 - 0·27) | 1·2 (0·62 - 2·32) |
| Indragiri Hulu | 441789 | 444548 | 453241 | 3 | 25 | 29 | 1 | 6 | 6 | 8·28 (3·04 - 22·52) | 9·42 (3·57 - 24·9) |
| Indragiri Hilir | 740598 | 654909 | 658025 | 11 | 15 | 28 | 1 | 2 | 4 | 1·54 (0·71 - 3·34) | 2·86 (1·47 - 5·58) |
| Pelalawan | 483622 | 390046 | 399264 | 2 | 17 | 14 | 0 | 4 | 4 | 10·54 (3·26 - 34·1) | 8·48 (2·47 - 29·1) |
| Siak | 489996 | 457940 | 466683 | 12 | 14 | 12 | 2 | 3 | 3 | 1·25 (0·58 - 2·69) | 1·05 (0·47 - 2·34) |
| Kampar | 871117 | 841332 | 857752 | 9 | 24 | 34 | 1 | 3 | 4 | 2·76 (1·33 - 5·75) | 3·84 (1·94 - 7·59) |
| Rokan Hulu | 692120 | 561385 | 570952 | 19 | 19 | 14 | 3 | 3 | 2 | 1·23 (0·65 - 2·33) | 0·89 (0·45 - 1·78) |
| Bengkalis | 573003 | 565569 | 573504 | 11 | 26 | 18 | 2 | 5 | 3 | 2·39 (1·21 - 4·74) | 1·63 (0·78 - 3·44) |
| Rokan Hilir | 714497 | 637161 | 646791 | 55 | 47 | 48 | 8 | 7 | 7 | 0·96 (0·65 - 1·41) | 0·96 (0·65 - 1·42) |
| Kepulauan Meranti | 185516 | 206116 | 209460 | 30 | 15 | 21 | 16 | 7 | 10 | 0·45 (0·25 - 0·82) | 0·62 (0·36 - 1·08) |
| Kota Pekanbaru | 1143359 | 983356 | 994585 | 98 | 116 | 124 | 9 | 12 | 12 | 1·38 (1·05 - 1·8) | 1·45 (1·12 - 1·89) |
| Kota Dumai | 308812 | 316782 | 323452 | 26 | 47 | 15 | 8 | 15 | 5 | 1·76 (1·1 - 2·83) | 0·55 (0·29 - 1·03) |
| **Jambi** | | | | | | | | | | | |
| Kerinci | 238682 | 250300 | 251900 | 0 | 7 | 5 | 0 | 3 | 2 | NA | NA |
| Merangin | 388928 | 354100 | 355700 | 2 | 17 | 15 | 1 | 5 | 4 | 9·34 (2·81 - 30·99) | 8·2 (2·39 - 28·19) |
| Sarolangun | 301908 | 290100 | 293600 | 2 | 2 | 13 | 1 | 1 | 4 | 1·04 (0·15 - 7·39) | 6·68 (1·85 - 24·19) |
| Batang Hari | 272879 | 301700 | 306700 | 19 | 17 | 26 | 7 | 6 | 8 | 0·81 (0·42 - 1·56) | 1·22 (0·67 - 2·2) |
| Muaro Jambi | 443364 | 402000 | 406700 | 1 | 7 | 21 | 0 | 2 | 5 | 7·72 (1·31 - 45·35) | 22·89 (5·8 - 90·33) |
| Tanjung Jabung Timur | 219985 | 229800 | 231800 | 0 | 7 | 7 | 0 | 3 | 3 | NA | NA |
| Tanjung Jabung Barat | 333932 | 317500 | 320600 | 6 | 16 | 11 | 2 | 5 | 3 | 2·8 (1·14 - 6·88) | 1·91 (0·72 - 5·08) |

| District | Number of populations | | | Number of reported mortalities | | | Mortality rate per 100,000 population | | | Mortality rate ratio for 2020 vs 2019 (95% CI) | Mortality rate ratio for 2021 vs 2019 (95% CI) |
|---|---|---|---|---|---|---|---|---|---|---|---|
| | 2019 | 2020 | 2021 | 2019 | 2020 | 2021 | 2019 | 2020 | 2021 | | |
| Tebo | 354485 | 337700 | 340900 | 3 | 13 | 3 | 1 | 4 | 1 | 4·55 (1·45 - 14·26) | 1·04 (0·21 - 5·15) |
| Bungo | 374770 | 362400 | 367200 | 11 | 17 | 15 | 3 | 5 | 4 | 1·6 (0·75 - 3·39) | 1·39 (0·64 - 3·02) |
| Kota Jambi | 604736 | 606200 | 612200 | 31 | 52 | 54 | 5 | 9 | 9 | 1·67 (1·08 - 2·6) | 1·72 (1·11 - 2·66) |
| Kota Sungai Penuh | 90910 | 96600 | 97800 | 3 | 3 | 6 | 3 | 3 | 6 | 0·94 (0·19 - 4·66) | 1·86 (0·48 - 7·27) |
| **South Sumatera** | | | | | | | | | | | |
| Ogan Komering Ulu | 368756 | 367603 | 371106 | 0 | 9 | 13 | 0 | 2 | 4 | NA | NA |
| Ogan Komering Ilir | 832151 | 769348 | 772742 | 30 | 19 | 37 | 4 | 2 | 5 | 0·69 (0·39 - 1·21) | 1·33 (0·82 - 2·15) |
| Muara Enim | 637556 | 612900 | 617846 | 47 | 45 | 30 | 7 | 7 | 5 | 1 (0·66 - 1·5) | 0·66 (0·42 - 1·04) |
| Lahat | 409348 | 430071 | 434939 | 4 | 12 | 7 | 1 | 3 | 2 | 2·86 (0·97 - 8·42) | 1·65 (0·49 - 5·56) |
| Musi Rawas | 405175 | 395570 | 398732 | 27 | 31 | 31 | 7 | 8 | 8 | 1·18 (0·7 - 1·97) | 1·17 (0·7 - 1·95) |
| Musi Banyuasin | 649085 | 622206 | 627070 | 21 | 44 | 42 | 3 | 7 | 7 | 2·19 (1·32 - 3·63) | 2·07 (1·24 - 3·46) |
| Banyu Asin | 857097 | 836914 | 843871 | 12 | 29 | 23 | 1 | 3 | 3 | 2·47 (1·29 - 4·74) | 1·95 (0·98 - 3·86) |
| Ogan Komering Ulu Selatan | 363004 | 408981 | 416616 | 0 | 9 | 10 | 0 | 2 | 2 | NA | NA |
| Ogan Komering Ulu Timur | 677080 | 649853 | 653062 | 23 | 22 | 24 | 3 | 3 | 4 | 1 (0·56 - 1·79) | 1·08 (0·61 - 1·92) |
| Ogan Ilir | 429595 | 416549 | 419401 | 22 | 26 | 21 | 5 | 6 | 5 | 1·22 (0·69 - 2·15) | 0·98 (0·54 - 1·78) |
| Empat Lawang | 250465 | 333622 | 343839 | 1 | 2 | 2 | 0 | 1 | 1 | 1·5 (0·14 - 16·29) | 1·46 (0·13 - 15·84) |
| Penukal Abab | 190062 | 194900 | 197290 | 3 | 11 | 5 | 2 | 6 | 3 | 3·58 (1·08 - 11·8) | 1·61 (0·39 - 6·63) |
| Musi Rawas Utara | 192540 | 188861 | 190420 | 0 | 9 | 13 | 0 | 5 | 7 | NA | NA |
| Kota Palembang | 1674243 | 1668848 | 1686073 | 65 | 93 | 76 | 4 | 6 | 5 | 1·44 (1·05 - 1·97) | 1·16 (0·83 - 1·62) |
| Kota Prabumulih | 188669 | 193196 | 195748 | 5 | 0 | 9 | 3 | 0 | 5 | NA | 1·73 (0·59 - 5·11) |
| Kota Pagar Alam | 139192 | 143844 | 145266 | 2 | 4 | 5 | 1 | 3 | 3 | 1·94 (0·37 - 10·25) | 2·4 (0·49 - 11·73) |
| Kota Lubuklinggau | 233178 | 234166 | 236828 | 12 | 10 | 13 | 5 | 4 | 5 | 0·83 (0·36 - 1·92) | 1·07 (0·49 - 2·34) |
| **Bengkulu** | | | | | | | | | | | |
| Bengkulu Selatan | 158410 | 166250 | 167989 | 16 | 9 | 8 | 10 | 5 | 5 | 0·54 (0·24 - 1·2) | 0·47 (0·21 - 1·08) |
| Rejang Lebong | 260900 | 276640 | 278793 | 21 | 8 | 9 | 8 | 3 | 3 | 0·36 (0·16 - 0·78) | 0·4 (0·19 - 0·85) |

| District | Number of populations | | | Number of reported mortalities | | | Mortality rate per 100,000 population | | | Mortality rate ratio for 2020 vs 2019 (95% CI) | Mortality rate ratio for 2021 vs 2019 (95% CI) |
|---|---|---|---|---|---|---|---|---|---|---|---|
| | 2019 | 2020 | 2021 | 2019 | 2020 | 2021 | 2019 | 2020 | 2021 | | |
| Bengkulu Utara | 310000 | 296520 | 299395 | 13 | 10 | 3 | 4 | 3 | 1 | 0·8 (0·35 - 1·83) | 0·24 (0·08 - 0·76) |
| Kaur | 121210 | 126550 | 127953 | 7 | 4 | 1 | 6 | 3 | 1 | 0·55 (0·16 - 1·84) | 0·14 (0·02 - 0·8) |
| Seluma | 193800 | 207880 | 210505 | 3 | 0 | 2 | 2 | 0 | 1 | NA | 0·61 (0·1 - 3·61) |
| Mukomuko | 193880 | 190500 | 193196 | 1 | 6 | 14 | 1 | 3 | 7 | 6·11 (0·96 - 38·98) | 14·05 (3·01 - 65·53) |
| Lebong | 116610 | 106290 | 106767 | 1 | 15 | 13 | 1 | 14 | 12 | 16·46 (3·72 - 72·87) | 14·2 (3·04 - 66·27) |
| Kepahiang | 137190 | 149740 | 151640 | 1 | 7 | 6 | 1 | 5 | 4 | 6·41 (1·04 - 39·65) | 5·43 (0·82 - 35·72) |
| Bengkulu Tengah | 114700 | 116710 | 118100 | 5 | 6 | 11 | 4 | 5 | 9 | 1·18 (0·36 - 3·86) | 2·14 (0·76 - 6) |
| Kota Bengkulu | 385140 | 373590 | 378604 | 11 | 11 | 10 | 3 | 3 | 3 | 1·03 (0·45 - 2·38) | 0·92 (0·39 - 2·18) |
| **Lampung** | | | | | | | | | | | |
| Lampung Barat | 302828 | 304874 | 302749 | 18 | 15 | 7 | 6 | 5 | 2 | 0·83 (0·42 - 1·64) | 0·39 (0·17 - 0·9) |
| Tanggamus | 598299 | 603706 | 645807 | 48 | 45 | 19 | 8 | 7 | 3 | 0·93 (0·62 - 1·4) | 0·37 (0·22 - 0·61) |
| Lampung Selatan | 1011286 | 1019789 | 1071727 | 3 | 30 | 23 | 0 | 3 | 2 | 9·92 (3·79 - 25·94) | 7·23 (2·59 - 20·21) |
| Lampung Timur | 1044320 | 1051994 | 1118115 | 4 | 42 | 15 | 0 | 4 | 1 | 10·42 (4·57 - 23·75) | 3·5 (1·25 - 9·85) |
| Lampung Tengah | 1281310 | 1290407 | 1477395 | 3 | 30 | 25 | 0 | 2 | 2 | 9·93 (3·8 - 25·97) | 7·23 (2·6 - 20·09) |
| Lampung Utara | 616897 | 618818 | 634117 | 7 | 18 | 20 | 1 | 3 | 3 | 2·56 (1·1 - 5·95) | 2·78 (1·22 - 6·34) |
| Way Kanan | 450109 | 453921 | 476871 | 4 | 11 | 16 | 1 | 2 | 3 | 2·73 (0·91 - 8·17) | 3·78 (1·36 - 10·46) |
| Tulangbawang | 450902 | 455891 | 430630 | 15 | 35 | 19 | 3 | 8 | 4 | 2·31 (1·28 - 4·15) | 1·33 (0·68 - 2·6) |
| Pesawaran | 444380 | 448410 | 481708 | 20 | 9 | 13 | 5 | 2 | 3 | 0·45 (0·21 - 0·96) | 0·6 (0·3 - 1·2) |
| Pringsewu | 400187 | 403115 | 406823 | 10 | 22 | 15 | 2 | 5 | 4 | 2·18 (1·05 - 4·53) | 1·48 (0·67 - 3·27) |
| Mesuji | 200198 | 200999 | 229772 | 3 | 13 | 7 | 1 | 6 | 3 | 4·32 (1·37 - 13·63) | 2·03 (0·54 - 7·65) |
| Tulang Bawang Barat | 273215 | 274905 | 287707 | 3 | 9 | 5 | 1 | 3 | 2 | 2·98 (0·86 - 10·34) | 1·58 (0·38 - 6·54) |
| Pesisir Barat | 154895 | 155883 | 163641 | 6 | 14 | 13 | 4 | 9 | 8 | 2·32 (0·92 - 5·87) | 2·05 (0·8 - 5·29) |
| Kota Bandar Lampung | 1051500 | 1068982 | 1184949 | 30 | 68 | 44 | 3 | 6 | 4 | 2·23 (1·47 - 3·39) | 1·3 (0·82 - 2·07) |
| Kota Metro | 167411 | 169507 | 169781 | 12 | 12 | 8 | 7 | 7 | 5 | 0·99 (0·44 - 2·2) | 0·66 (0·27 - 1·6) |
| **Bangka Belitung** | | | | | | | | | | | |

| District | Number of populations | | | Number of reported mortalities | | | Mortality rate per 100,000 population | | | Mortality rate ratio for 2020 vs 2019 (95% CI) | Mortality rate ratio for 2021 vs 2019 (95% CI) |
|---|---|---|---|---|---|---|---|---|---|---|---|
| | 2019 | 2020 | 2021 | 2019 | 2020 | 2021 | 2019 | 2020 | 2021 | | |
| Bangka | 337337 | 343821 | 337286 | 33 | 23 | 29 | 10 | 7 | 9 | 0·68 (0·4 - 1·16) | 0·88 (0·53 - 1·45) |
| Belitung | 189824 | 193493 | 189752 | 14 | 21 | 11 | 7 | 11 | 6 | 1·47 (0·75 - 2·88) | 0·79 (0·36 - 1·73) |
| Bangka Barat | 213163 | 217332 | 213282 | 12 | 6 | 10 | 6 | 3 | 5 | 0·49 (0·19 - 1·28) | 0·83 (0·36 - 1·93) |
| Bangka Tengah | 196196 | 200016 | 196060 | 13 | 17 | 8 | 7 | 8 | 4 | 1·28 (0·62 - 2·64) | 0·62 (0·26 - 1·47) |
| Bangka Selatan | 209973 | 213966 | 209815 | 6 | 12 | 8 | 3 | 6 | 4 | 1·96 (0·75 - 5·13) | 1·33 (0·46 - 3·83) |
| Belitung Timur | 129572 | 132069 | 129411 | 7 | 6 | 8 | 5 | 5 | 6 | 0·84 (0·28 - 2·5) | 1·14 (0·42 - 3·15) |
| Kota Pangkal Pinang | 212727 | 216893 | 212639 | 11 | 22 | 8 | 5 | 10 | 4 | 1·96 (0·96 - 3·99) | 0·73 (0·29 - 1·8) |
| **Riau Island** | | | | | | | | | | | |
| Karimun | 232800 | 253457 | 259450 | 24 | 14 | 4 | 10 | 6 | 2 | 0·54 (0·28 - 1·02) | 0·15 (0·06 - 0·37) |
| Bintan | 159400 | 159518 | 162560 | 11 | 11 | 4 | 7 | 7 | 2 | 1 (0·43 - 2·3) | 0·36 (0·12 - 1·07) |
| Natuna | 77770 | 81495 | 83360 | 3 | 3 | 1 | 4 | 4 | 1 | 0·95 (0·19 - 4·73) | 0·31 (0·04 - 2·64) |
| Lingga | 89780 | 98633 | 100660 | 12 | 15 | 5 | 13 | 15 | 5 | 1·14 (0·53 - 2·43) | 0·37 (0·14 - 1·01) |
| Kepulauan Anambas | 42310 | 47402 | 48740 | 1 | 0 | 2 | 2 | 0 | 4 | NA | 1·74 (0·16 - 18·58) |
| Kota Batam | 1376010 | 1196396 | 1230100 | 68 | 90 | 98 | 5 | 8 | 8 | 1·52 (1·11 - 2·08) | 1·61 (1·19 - 2·19) |
| Kota Tanjung Pinang | 211580 | 227663 | 233370 | 28 | 29 | 30 | 13 | 13 | 13 | 0·96 (0·57 - 1·62) | 0·97 (0·58 - 1·63) |
| **DKI Jakarta** | | | | | | | | | | | |
| Kepulauan Seribu | 24300 | 27750 | 28240 | 4 | 5 | 1 | 16 | 18 | 4 | 1·09 (0·29 - 4·07) | 0·22 (0·03 - 1·57) |
| Kota Jakarta Selatan | 2264700 | 2226810 | 2233855 | 202 | 143 | 116 | 9 | 6 | 5 | 0·72 (0·58 - 0·89) | 0·58 (0·46 - 0·73) |
| Kota Jakarta Timur | 2937860 | 3037140 | 3056300 | 241 | 200 | 143 | 8 | 7 | 5 | 0·8 (0·67 - 0·97) | 0·57 (0·47 - 0·7) |
| Kota Jakarta Pusat | 928110 | 1056900 | 1066460 | 262 | 350 | 224 | 28 | 33 | 21 | 1·17 (1 - 1·38) | 0·74 (0·62 - 0·89) |
| Kota Jakarta Barat | 2589930 | 2434510 | 2440073 | 65 | 93 | 60 | 3 | 4 | 2 | 1·52 (1·11 - 2·08) | 0·98 (0·69 - 1·39) |
| Kota Jakarta Utara | 1812910 | 1778980 | 1784753 | 75 | 43 | 51 | 4 | 2 | 3 | 0·58 (0·4 - 0·85) | 0·69 (0·48 - 0·98) |
| **West Java** | | | | | | | | | | | |
| Bogor | 5965410 | 6088233 | 5489540 | 166 | 216 | 174 | 3 | 4 | 3 | 1·27 (1·04 - 1·56) | 1·14 (0·92 - 1·41) |
| Cianjur | 2466272 | 2470219 | 2761480 | 5 | 16 | 21 | 0 | 1 | 1 | 3·19 (1·24 - 8·26) | 3·75 (1·51 - 9·3) |

| District | Number of populations | | | Number of reported mortalities | | | Mortality rate per 100,000 population | | | Mortality rate ratio for 2020 vs 2019 (95% CI) | Mortality rate ratio for 2021 vs 2019 (95% CI) |
|---|---|---|---|---|---|---|---|---|---|---|---|
| | 2019 | 2020 | 2021 | 2019 | 2020 | 2021 | 2019 | 2020 | 2021 | | |
| Sukabumi | 2263072 | 2264328 | 2506680 | 20 | 82 | 61 | 1 | 4 | 2 | 4·1 (2·61 - 6·43) | 2·75 (1·7 - 4·47) |
| Bandung | 3775279 | 3831505 | 3666160 | 147 | 183 | 123 | 4 | 5 | 3 | 1·23 (0·99 - 1·52) | 0·86 (0·68 - 1·09) |
| Garut | 2622425 | 2636637 | 2604790 | 62 | 89 | 76 | 2 | 3 | 3 | 1·43 (1·03 - 1·97) | 1·23 (0·88 - 1·72) |
| Tasikmalaya | 1754128 | 1755710 | 1883730 | 132 | 84 | 100 | 8 | 5 | 5 | 0·64 (0·48 - 0·83) | 0·71 (0·54 - 0·91) |
| Ciamis | 1195176 | 1201685 | 1237730 | 28 | 40 | 41 | 2 | 3 | 3 | 1·42 (0·88 - 2·3) | 1·41 (0·88 - 2·28) |
| Kuningan | 1080804 | 1087105 | 1180390 | 78 | 62 | 58 | 7 | 6 | 5 | 0·79 (0·57 - 1·1) | 0·68 (0·49 - 0·95) |
| Cirebon | 2192903 | 2209633 | 2290970 | 123 | 105 | 65 | 6 | 5 | 3 | 0·85 (0·65 - 1·1) | 0·51 (0·38 - 0·68) |
| Majalengka | 1205034 | 1210709 | 1318970 | 23 | 33 | 29 | 2 | 3 | 2 | 1·43 (0·84 - 2·43) | 1·15 (0·67 - 1·99) |
| Sumedang | 1152400 | 1154428 | 1159350 | 71 | 54 | 51 | 6 | 5 | 4 | 0·76 (0·53 - 1·08) | 0·71 (0·5 - 1·02) |
| Indramayu | 1728469 | 1737624 | 1851380 | 79 | 34 | 26 | 5 | 2 | 1 | 0·43 (0·29 - 0·63) | 0·31 (0·2 - 0·47) |
| Subang | 1595825 | 1612576 | 1608590 | 90 | 76 | 66 | 6 | 5 | 4 | 0·84 (0·62 - 1·13) | 0·73 (0·53 - 1) |
| Purwakarta | 962893 | 971889 | 191147 | 24 | 33 | 44 | 2 | 3 | 23 | 1·36 (0·81 - 2·3) | 9·24 (6·14 - 13·89) |
| Karawang | 2353915 | 2370488 | 2468580 | 84 | 123 | 111 | 4 | 5 | 4 | 1·45 (1·1 - 1·92) | 1·26 (0·95 - 1·67) |
| Bekasi | 3763886 | 3899017 | 3157960 | 48 | 47 | 54 | 1 | 1 | 2 | 0·95 (0·63 - 1·41) | 1·34 (0·91 - 1·98) |
| Bandung Barat | 1699896 | 1714982 | 1814230 | 51 | 42 | 41 | 3 | 2 | 2 | 0·82 (0·54 - 1·23) | 0·75 (0·5 - 1·13) |
| Pangandaran | 399284 | 401493 | 427610 | 2 | 16 | 7 | 1 | 4 | 2 | 7·96 (2·31 - 27·39) | 3·27 (0·74 - 14·4) |
| Kota Bogor | 1112081 | 1126927 | 1052360 | 35 | 38 | 49 | 3 | 3 | 5 | 1·07 (0·68 - 1·7) | 1·48 (0·96 - 2·28) |
| Kota Sukabumi | 328680 | 330691 | 350800 | 13 | 28 | 25 | 4 | 8 | 7 | 2·14 (1·13 - 4·07) | 1·8 (0·93 - 3·49) |
| Kota Bandung | 2507888 | 2510103 | 2452940 | 196 | 274 | 196 | 8 | 11 | 8 | 1·4 (1·16 - 1·68) | 1·02 (0·84 - 1·25) |
| Kota Cirebon | 319312 | 322322 | 336860 | 18 | 23 | 25 | 6 | 7 | 7 | 1·27 (0·68 - 2·34) | 1·32 (0·72 - 2·41) |
| Kota Bekasi | 3003923 | 3075690 | 2564940 | 78 | 88 | 72 | 3 | 3 | 3 | 1·1 (0·81 - 1·49) | 1·08 (0·78 - 1·49) |
| Kota Depok | 2406826 | 2484186 | 2085940 | 88 | 59 | 85 | 4 | 2 | 4 | 0·65 (0·47 - 0·9) | 1·11 (0·83 - 1·5) |
| Kota Cimahi | 614304 | 620393 | 571630 | 20 | 24 | 16 | 3 | 4 | 3 | 1·19 (0·66 - 2·15) | 0·86 (0·45 - 1·66) |
| Kota Tasikmalaya | 663517 | 663986 | 723920 | 40 | 23 | 37 | 6 | 3 | 5 | 0·57 (0·35 - 0·95) | 0·85 (0·54 - 1·33) |
| Kota Banjar | 183110 | 183299 | 203420 | 5 | 6 | 5 | 3 | 3 | 2 | 1·2 (0·37 - 3·92) | 0·9 (0·26 - 3·11) |

| District | Number of populations | | | Number of reported mortalities | | | Mortality rate per 100,000 population | | | Mortality rate ratio for 2020 vs 2019 (95% CI) | Mortality rate ratio for 2021 vs 2019 (95% CI) |
|---|---|---|---|---|---|---|---|---|---|---|---|
| | 2019 | 2020 | 2021 | 2019 | 2020 | 2021 | 2019 | 2020 | 2021 | | |
| **Central Java** | | | | | | | | | | | |
| Cilacap | 1727098 | 1944857 | 1963824 | 89 | 113 | 92 | 5 | 6 | 5 | 1·13 (0·85 - 1·49) | 0·91 (0·68 - 1·22) |
| Banyumas | 1693006 | 1776918 | 1789630 | 66 | 165 | 141 | 4 | 9 | 8 | 2·38 (1·81 - 3·14) | 2·02 (1·52 - 2·69) |
| Purbalingga | 933989 | 998561 | 1007794 | 37 | 36 | 41 | 4 | 4 | 4 | 0·91 (0·58 - 1·44) | 1·03 (0·66 - 1·6) |
| Banjarnegara | 923192 | 1017767 | 1026866 | 34 | 31 | 17 | 4 | 3 | 2 | 0·83 (0·51 - 1·34) | 0·45 (0·25 - 0·79) |
| Kebumen | 1197982 | 1350438 | 1361913 | 59 | 127 | 111 | 5 | 9 | 8 | 1·91 (1·41 - 2·59) | 1·65 (1·21 - 2·26) |
| Purworejo | 718316 | 769880 | 773588 | 33 | 37 | 36 | 5 | 5 | 5 | 1·05 (0·65 - 1·67) | 1·01 (0·63 - 1·62) |
| Wonosobo | 790504 | 879124 | 886613 | 7 | 28 | 28 | 1 | 3 | 3 | 3·6 (1·66 - 7·8) | 3·57 (1·64 - 7·74) |
| Magelang | 1290591 | 1299859 | 1305512 | 30 | 23 | 20 | 2 | 2 | 2 | 0·76 (0·44 - 1·31) | 0·66 (0·38 - 1·16) |
| Boyolali | 984807 | 1062713 | 1070247 | 21 | 24 | 22 | 2 | 2 | 2 | 1·06 (0·59 - 1·9) | 0·96 (0·53 - 1·75) |
| Klaten | 1174986 | 1260506 | 1267272 | 81 | 25 | 30 | 7 | 2 | 2 | 0·29 (0·19 - 0·44) | 0·34 (0·23 - 0·51) |
| Sukoharjo | 891912 | 907587 | 911603 | 34 | 28 | 20 | 4 | 3 | 2 | 0·81 (0·49 - 1·33) | 0·58 (0·33 - 0·99) |
| Wonogiri | 959492 | 1043177 | 1049292 | 55 | 52 | 31 | 6 | 5 | 3 | 0·87 (0·6 - 1·27) | 0·52 (0·33 - 0·79) |
| Karanganyar | 886519 | 931963 | 938808 | 6 | 8 | 8 | 1 | 1 | 1 | 1·27 (0·44 - 3·65) | 1·26 (0·44 - 3·62) |
| Sragen | 890518 | 976951 | 983641 | 22 | 21 | 19 | 2 | 2 | 2 | 0·87 (0·48 - 1·58) | 0·78 (0·42 - 1·44) |
| Grobogan | 1377788 | 1453526 | 1460873 | 34 | 19 | 16 | 2 | 1 | 1 | 0·53 (0·3 - 0·92) | 0·44 (0·25 - 0·79) |
| Blora | 865013 | 884333 | 886147 | 37 | 69 | 36 | 4 | 8 | 4 | 1·82 (1·23 - 2·7) | 0·95 (0·6 - 1·5) |
| Rembang | 638188 | 645333 | 647766 | 31 | 34 | 31 | 5 | 5 | 5 | 1·08 (0·67 - 1·76) | 0·99 (0·6 - 1·62) |
| Pati | 1259590 | 1324188 | 1330983 | 112 | 76 | 88 | 9 | 6 | 7 | 0·65 (0·48 - 0·86) | 0·74 (0·56 - 0·98) |
| Kudus | 871311 | 849184 | 852443 | 35 | 44 | 48 | 4 | 5 | 6 | 1·29 (0·83 - 2·01) | 1·4 (0·91 - 2·16) |
| Jepara | 1257912 | 1184947 | 1188510 | 26 | 35 | 13 | 2 | 3 | 1 | 1·43 (0·86 - 2·37) | 0·53 (0·27 - 1·02) |
| Demak | 1162805 | 1203956 | 1212377 | 25 | 27 | 24 | 2 | 2 | 2 | 1·04 (0·61 - 1·8) | 0·92 (0·53 - 1·61) |
| Semarang | 1053786 | 1053094 | 1059844 | 11 | 12 | 6 | 1 | 1 | 1 | 1·09 (0·48 - 2·47) | 0·54 (0·2 - 1·44) |
| Temanggung | 772018 | 790174 | 794403 | 25 | 38 | 27 | 3 | 5 | 3 | 1·49 (0·9 - 2·45) | 1·05 (0·61 - 1·81) |
| Kendal | 971086 | 1018505 | 1025020 | 28 | 42 | 23 | 3 | 4 | 2 | 1·43 (0·89 - 2·3) | 0·78 (0·45 - 1·35) |

| District | Number of populations | | | Number of reported mortalities | | | Mortality rate per 100,000 population | | | Mortality rate ratio for 2020 vs 2019 (95% CI) | Mortality rate ratio for 2021 vs 2019 (95% CI) |
|---|---|---|---|---|---|---|---|---|---|---|---|
| | 2019 | 2020 | 2021 | 2019 | 2020 | 2021 | 2019 | 2020 | 2021 | | |
| Batang | 768583 | 801718 | 807005 | 40 | 35 | 16 | 5 | 4 | 2 | 0·84 (0·53 - 1·32) | 0·38 (0·22 - 0·67) |
| Pekalongan | 897711 | 968821 | 976504 | 22 | 44 | 22 | 2 | 5 | 2 | 1·85 (1·12 - 3·07) | 0·92 (0·51 - 1·66) |
| Pemalang | 1302813 | 1471489 | 1484209 | 25 | 23 | 19 | 2 | 2 | 1 | 0·81 (0·46 - 1·43) | 0·67 (0·37 - 1·21) |
| Tegal | 1440698 | 1596996 | 1608611 | 153 | 128 | 39 | 11 | 8 | 2 | 0·75 (0·6 - 0·95) | 0·23 (0·17 - 0·31) |
| Brebes | 1809096 | 1978759 | 1992685 | 37 | 63 | 47 | 2 | 3 | 2 | 1·56 (1·04 - 2·33) | 1·15 (0·75 - 1·77) |
| Kota Magelang | 122111 | 121526 | 121610 | 38 | 29 | 30 | 31 | 24 | 25 | 0·77 (0·47 - 1·24) | 0·79 (0·49 - 1·28) |
| Kota Surakarta | 519587 | 522364 | 522728 | 94 | 64 | 55 | 18 | 12 | 11 | 0·68 (0·49 - 0·93) | 0·58 (0·42 - 0·81) |
| Kota Salatiga | 194084 | 192322 | 193525 | 8 | 12 | 7 | 4 | 6 | 4 | 1·51 (0·62 - 3·68) | 0·88 (0·32 - 2·42) |
| Kota Semarang | 1814110 | 1653524 | 1656564 | 108 | 149 | 130 | 6 | 9 | 8 | 1·51 (1·18 - 1·94) | 1·32 (1·02 - 1·7) |
| Kota Pekalongan | 307097 | 307150 | 308310 | 33 | 17 | 14 | 11 | 6 | 5 | 0·52 (0·29 - 0·91) | 0·42 (0·23 - 0·77) |
| Kota Tegal | 249905 | 273825 | 275781 | 19 | 18 | 9 | 8 | 7 | 3 | 0·86 (0·45 - 1·65) | 0·43 (0·2 - 0·93) |
| **Di Yogyakarta** | | | | | | | | | | | |
| Kulon Progo | 432058 | 437373 | 442724 | 24 | 22 | 12 | 6 | 5 | 3 | 0·91 (0·51 - 1·61) | 0·49 (0·25 - 0·96) |
| Bantul | 1022788 | 1036489 | 1050308 | 87 | 28 | 28 | 9 | 3 | 3 | 0·32 (0·21 - 0·48) | 0·31 (0·21 - 0·47) |
| Gunung Kidul | 749229 | 758316 | 767464 | 25 | 27 | 19 | 3 | 4 | 2 | 1·07 (0·62 - 1·84) | 0·74 (0·41 - 1·34) |
| Sleman | 1231246 | 1248258 | 1256429 | 43 | 41 | 38 | 3 | 3 | 3 | 0·94 (0·61 - 1·44) | 0·87 (0·56 - 1·34) |
| Kota Yogyakarta | 433267 | 438761 | 444295 | 90 | 50 | 48 | 21 | 11 | 11 | 0·55 (0·39 - 0·77) | 0·52 (0·37 - 0·73) |
| **East Java** | | | | | | | | | | | |
| Pacitan | 555304 | 586110 | 589108 | 37 | 41 | 11 | 7 | 7 | 2 | 1·05 (0·67 - 1·64) | 0·28 (0·15 - 0·53) |
| Ponorogo | 871370 | 949318 | 955839 | 60 | 60 | 31 | 7 | 6 | 3 | 0·92 (0·64 - 1·31) | 0·47 (0·31 - 0·72) |
| Trenggalek | 696295 | 731125 | 734888 | 18 | 40 | 16 | 3 | 5 | 2 | 2·12 (1·23 - 3·64) | 0·84 (0·43 - 1·65) |
| Tulungagung | 1039284 | 1089775 | 1096588 | 84 | 59 | 38 | 8 | 5 | 3 | 0·67 (0·48 - 0·93) | 0·43 (0·3 - 0·62) |
| Blitar | 1160677 | 1223745 | 1231013 | 50 | 43 | 30 | 4 | 4 | 2 | 0·82 (0·54 - 1·23) | 0·57 (0·36 - 0·88) |
| Kediri | 1574272 | 1635294 | 1644400 | 42 | 98 | 65 | 3 | 6 | 4 | 2·25 (1·58 - 3·19) | 1·48 (1·01 - 2·18) |
| Malang | 2606204 | 2654448 | 2668296 | 111 | 108 | 106 | 4 | 4 | 4 | 0·96 (0·73 - 1·25) | 0·93 (0·71 - 1·22) |

| District | Number of populations | | | Number of reported mortalities | | | Mortality rate per 100,000 population | | | Mortality rate ratio for 2020 vs 2019 (95% CI) | Mortality rate ratio for 2021 vs 2019 (95% CI) |
|---|---|---|---|---|---|---|---|---|---|---|---|
| | 2019 | 2020 | 2021 | 2019 | 2020 | 2021 | 2019 | 2020 | 2021 | | |
| Lumajang | 1042395 | 1119251 | 1127094 | 48 | 60 | 45 | 5 | 5 | 4 | 1·16 (0·8 - 1·7) | 0·87 (0·58 - 1·3) |
| Jember | 2450668 | 2536729 | 2550360 | 174 | 160 | 121 | 7 | 6 | 5 | 0·89 (0·72 - 1·1) | 0·67 (0·53 - 0·84) |
| Banyuwangi | 1613991 | 1708114 | 1718462 | 152 | 134 | 96 | 9 | 8 | 6 | 0·83 (0·66 - 1·05) | 0·59 (0·46 - 0·76) |
| Bondowoso | 775715 | 776151 | 778525 | 54 | 72 | 62 | 7 | 9 | 8 | 1·33 (0·94 - 1·89) | 1·14 (0·79 - 1·65) |
| Situbondo | 682978 | 685967 | 688337 | 38 | 46 | 36 | 6 | 7 | 5 | 1·21 (0·78 - 1·85) | 0·94 (0·6 - 1·48) |
| Probolinggo | 1168503 | 1152537 | 1155894 | 93 | 78 | 57 | 8 | 7 | 5 | 0·85 (0·63 - 1·15) | 0·62 (0·45 - 0·86) |
| Pasuruan | 1627396 | 1605969 | 1611805 | 103 | 64 | 62 | 6 | 4 | 4 | 0·63 (0·46 - 0·86) | 0·61 (0·44 - 0·83) |
| Sidoarjo | 2249476 | 2082801 | 2091930 | 111 | 103 | 41 | 5 | 5 | 2 | 1 (0·77 - 1·31) | 0·4 (0·28 - 0·56) |
| Mojokerto | 1117688 | 1119209 | 1125522 | 24 | 33 | 26 | 2 | 3 | 2 | 1·37 (0·81 - 2·32) | 1·08 (0·62 - 1·87) |
| Jombang | 1263814 | 1318062 | 1325914 | 68 | 73 | 37 | 5 | 6 | 3 | 1·03 (0·74 - 1·43) | 0·52 (0·35 - 0·77) |
| Nganjuk | 1054611 | 1103902 | 1109683 | 21 | 17 | 22 | 2 | 2 | 2 | 0·77 (0·41 - 1·46) | 1 (0·55 - 1·81) |
| Madiun | 682684 | 744350 | 750143 | 45 | 33 | 24 | 7 | 4 | 3 | 0·67 (0·43 - 1·05) | 0·49 (0·3 - 0·79) |
| Magetan | 628977 | 670812 | 674133 | 23 | 26 | 22 | 4 | 4 | 3 | 1·06 (0·6 - 1·86) | 0·89 (0·5 - 1·6) |
| Ngawi | 830108 | 870057 | 873346 | 45 | 55 | 52 | 5 | 6 | 6 | 1·17 (0·79 - 1·73) | 1·1 (0·74 - 1·64) |
| Bojonegoro | 1249692 | 1301635 | 1307602 | 48 | 33 | 25 | 4 | 3 | 2 | 0·66 (0·43 - 1·02) | 0·5 (0·31 - 0·8) |
| Tuban | 1172790 | 1198012 | 1203127 | 56 | 64 | 31 | 5 | 5 | 3 | 1·12 (0·78 - 1·6) | 0·54 (0·35 - 0·83) |
| Lamongan | 1189106 | 1344165 | 1356027 | 63 | 54 | 80 | 5 | 4 | 6 | 0·76 (0·53 - 1·09) | 1·11 (0·8 - 1·55) |
| Gresik | 1312881 | 1311215 | 1320570 | 91 | 50 | 65 | 7 | 4 | 5 | 0·55 (0·39 - 0·77) | 0·71 (0·52 - 0·97) |
| Bangkalan | 986672 | 1060377 | 1071712 | 23 | 39 | 26 | 2 | 4 | 2 | 1·58 (0·95 - 2·63) | 1·04 (0·59 - 1·82) |
| Sampang | 978875 | 969694 | 976020 | 42 | 45 | 51 | 4 | 5 | 5 | 1·08 (0·71 - 1·65) | 1·22 (0·81 - 1·83) |
| Pamekasan | 879992 | 850057 | 853507 | 68 | 58 | 35 | 8 | 7 | 4 | 0·88 (0·62 - 1·25) | 0·53 (0·36 - 0·79) |
| Sumenep | 1088910 | 1124436 | 1129822 | 91 | 78 | 82 | 8 | 7 | 7 | 0·83 (0·61 - 1·12) | 0·87 (0·64 - 1·17) |
| Kota Kediri | 287409 | 286796 | 287962 | 33 | 27 | 42 | 11 | 9 | 15 | 0·82 (0·49 - 1·36) | 1·27 (0·81 - 2) |
| Kota Blitar | 141876 | 149149 | 150371 | 18 | 24 | 14 | 13 | 16 | 9 | 1·27 (0·69 - 2·33) | 0·73 (0·37 - 1·47) |
| Kota Malang | 870682 | 843810 | 844933 | 41 | 47 | 71 | 5 | 6 | 8 | 1·18 (0·78 - 1·8) | 1·78 (1·22 - 2·61) |

| District | Number of populations | | | Number of reported mortalities | | | Mortality rate per 100,000 population | | | Mortality rate ratio for 2020 vs 2019 (95% CI) | Mortality rate ratio for 2021 vs 2019 (95% CI) |
|---|---|---|---|---|---|---|---|---|---|---|---|
| | 2019 | 2020 | 2021 | 2019 | 2020 | 2021 | 2019 | 2020 | 2021 | | |
| Kota Probolinggo | 237208 | 239649 | 241202 | 52 | 16 | 21 | 22 | 7 | 9 | 0·3 (0·18 - 0·52) | 0·4 (0·24 - 0·65) |
| Kota Pasuruan | 200422 | 208006 | 209528 | 24 | 17 | 16 | 12 | 8 | 8 | 0·68 (0·37 - 1·27) | 0·64 (0·34 - 1·19) |
| Kota Mojokerto | 129014 | 132434 | 133272 | 17 | 14 | 23 | 13 | 11 | 17 | 0·8 (0·4 - 1·63) | 1·31 (0·7 - 2·45) |
| Kota Madiun | 177007 | 195175 | 196917 | 34 | 37 | 24 | 19 | 19 | 12 | 0·99 (0·62 - 1·57) | 0·63 (0·38 - 1·07) |
| Kota Surabaya | 2896195 | 2874314 | 2880284 | 174 | 144 | 91 | 6 | 5 | 3 | 0·83 (0·67 - 1·04) | 0·53 (0·41 - 0·67) |
| Kota Batu | 207490 | 213046 | 214653 | 4 | 4 | 3 | 2 | 2 | 1 | 0·97 (0·24 - 3·89) | 0·72 (0·16 - 3·22) |
| **Banten** | | | | | | | | | | | |
| Pandeglang | 1211909 | 1272687 | 1288314 | 68 | 71 | 43 | 6 | 6 | 3 | 0·99 (0·71 - 1·39) | 0·59 (0·41 - 0·87) |
| Lebak | 1302608 | 1386793 | 1407857 | 48 | 31 | 21 | 4 | 2 | 1 | 0·61 (0·39 - 0·95) | 0·4 (0·25 - 0·66) |
| Tangerang | 3800787 | 3245619 | 3293533 | 123 | 104 | 77 | 3 | 3 | 2 | 0·99 (0·76 - 1·29) | 0·72 (0·54 - 0·96) |
| Serang | 1508397 | 1622630 | 1647790 | 61 | 61 | 41 | 4 | 4 | 2 | 0·93 (0·65 - 1·33) | 0·62 (0·42 - 0·91) |
| Kota Tangerang | 2229901 | 1895486 | 1911914 | 15 | 80 | 84 | 1 | 4 | 4 | 6·27 (3·88 - 10·15) | 6·53 (4·06 - 10·52) |
| Kota Cilegon | 437205 | 434896 | 441761 | 63 | 42 | 38 | 14 | 10 | 9 | 0·67 (0·45 - 0·99) | 0·6 (0·4 - 0·89) |
| Kota Serang | 688603 | 692101 | 704618 | 36 | 50 | 45 | 5 | 7 | 6 | 1·38 (0·9 - 2·12) | 1·22 (0·79 - 1·89) |
| Kota Tangerang Selatan | 1747906 | 1354350 | 1365688 | 61 | 53 | 45 | 3 | 4 | 3 | 1·12 (0·78 - 1·62) | 0·94 (0·64 - 1·39) |
| **Bali** | | | | | | | | | | | |
| Jembrana | 278700 | 317100 | 321900 | 9 | 27 | 16 | 3 | 9 | 5 | 2·64 (1·28 - 5·45) | 1·54 (0·68 - 3·46) |
| Tabanan | 446700 | 461600 | 465300 | 20 | 27 | 31 | 4 | 6 | 7 | 1·31 (0·73 - 2·33) | 1·49 (0·85 - 2·6) |
| Badung | 678900 | 548200 | 549300 | 22 | 27 | 16 | 3 | 5 | 3 | 1·52 (0·87 - 2·66) | 0·9 (0·47 - 1·71) |
| Gianyar | 514300 | 515300 | 519500 | 16 | 22 | 14 | 3 | 4 | 3 | 1·37 (0·72 - 2·61) | 0·87 (0·42 - 1·77) |
| Klungkung | 179100 | 206900 | 210100 | 4 | 11 | 3 | 2 | 5 | 1 | 2·38 (0·79 - 7·22) | 0·64 (0·14 - 2·82) |
| Bangli | 227600 | 258700 | 262500 | 7 | 4 | 6 | 3 | 2 | 2 | 0·5 (0·15 - 1·68) | 0·74 (0·25 - 2·2) |
| Karangasem | 417000 | 492400 | 500800 | 33 | 32 | 17 | 8 | 6 | 3 | 0·82 (0·51 - 1·33) | 0·43 (0·24 - 0·76) |
| Buleleng | 661900 | 791800 | 806600 | 69 | 59 | 72 | 10 | 7 | 9 | 0·71 (0·51 - 1·01) | 0·86 (0·62 - 1·19) |
| Kota Denpasar | 957800 | 725300 | 726600 | 142 | 118 | 89 | 15 | 16 | 12 | 1·1 (0·86 - 1·4) | 0·83 (0·63 - 1·08) |

| District | Number of populations | | | Number of reported mortalities | | | Mortality rate per 100,000 population | | | Mortality rate ratio for 2020 vs 2019 (95% CI) | Mortality rate ratio for 2021 vs 2019 (95% CI) |
|---|---|---|---|---|---|---|---|---|---|---|---|
| | 2019 | 2020 | 2021 | 2019 | 2020 | 2021 | 2019 | 2020 | 2021 | | |
| **West Nusa Tenggara** | | | | | | | | | | | |
| Lombok Barat | 694985 | 704586 | 731800 | 53 | 40 | 35 | 8 | 6 | 5 | 0·74 (0·49 - 1·12) | 0·63 (0·41 - 0·96) |
| Lombok Tengah | 947488 | 955411 | 1049700 | 39 | 52 | 42 | 4 | 5 | 4 | 1·32 (0·87 - 2) | 0·97 (0·63 - 1·5) |
| Lombok Timur | 1200612 | 1208594 | 1343900 | 55 | 51 | 68 | 5 | 4 | 5 | 0·92 (0·63 - 1·35) | 1·1 (0·77 - 1·58) |
| Sumbawa | 457671 | 461502 | 519800 | 26 | 27 | 23 | 6 | 6 | 4 | 1·03 (0·6 - 1·76) | 0·78 (0·45 - 1·36) |
| Dompu | 252288 | 255569 | 238200 | 8 | 10 | 4 | 3 | 4 | 2 | 1·23 (0·49 - 3·12) | 0·53 (0·16 - 1·72) |
| Bima | 488577 | 493198 | 520400 | 10 | 29 | 12 | 2 | 6 | 2 | 2·87 (1·45 - 5·71) | 1·13 (0·49 - 2·61) |
| Sumbawa Barat | 148606 | 152437 | 148500 | 3 | 10 | 13 | 2 | 7 | 9 | 3·25 (0·96 - 10·99) | 4·34 (1·37 - 13·69) |
| Lombok Utara | 220412 | 222212 | 251500 | 21 | 21 | 9 | 10 | 9 | 4 | 0·99 (0·54 - 1·82) | 0·38 (0·18 - 0·8) |
| Kota Mataram | 486715 | 495681 | 432000 | 43 | 51 | 55 | 9 | 10 | 13 | 1·16 (0·78 - 1·75) | 1·44 (0·97 - 2·14) |
| Kota Bima | 173031 | 176432 | 156200 | 12 | 0 | 1 | 7 | 0 | 1 | NA | 0·09 (0·02 - 0·47) |
| **East Nusa Tenggara** | | | | | | | | | | | |
| Sumba Barat | 129710 | 131600 | 148252 | 6 | 6 | 7 | 5 | 5 | 5 | 0·99 (0·32 - 3·06) | 1·02 (0·34 - 3·04) |
| Sumba Timur | 258486 | 261503 | 246618 | 6 | 8 | 6 | 2 | 3 | 2 | 1·32 (0·46 - 3·79) | 1·05 (0·34 - 3·25) |
| Kupang | 403582 | 421618 | 372101 | 9 | 24 | 14 | 2 | 6 | 4 | 2·55 (1·22 - 5·34) | 1·69 (0·74 - 3·86) |
| Timor Tengah Selatan | 467990 | 469673 | 457406 | 12 | 32 | 15 | 3 | 7 | 3 | 2·66 (1·4 - 5·03) | 1·28 (0·6 - 2·73) |
| Timor Tengah Utara | 254171 | 256299 | 262698 | 11 | 18 | 13 | 4 | 7 | 5 | 1·62 (0·77 - 3·41) | 1·14 (0·51 - 2·55) |
| Belu | 220115 | 223176 | 220764 | 12 | 17 | 5 | 5 | 8 | 2 | 1·4 (0·67 - 2·92) | 0·42 (0·15 - 1·14) |
| Alor | 205599 | 206806 | 213994 | 28 | 29 | 25 | 14 | 14 | 12 | 1·03 (0·61 - 1·73) | 0·86 (0·5 - 1·47) |
| Lembata | 143074 | 145685 | 137631 | 3 | 5 | 6 | 2 | 3 | 4 | 1·64 (0·4 - 6·75) | 2·08 (0·54 - 8·06) |
| Flores Timur | 255916 | 257785 | 281001 | 8 | 20 | 11 | 3 | 8 | 4 | 2·48 (1·12 - 5·48) | 1·25 (0·5 - 3·11) |
| Sikka | 320401 | 321790 | 324252 | 25 | 39 | 23 | 8 | 12 | 7 | 1·55 (0·94 - 2·56) | 0·91 (0·52 - 1·6) |
| Ende | 273929 | 274599 | 272078 | 4 | 3 | 7 | 1 | 1 | 3 | 0·75 (0·17 - 3·33) | 1·76 (0·52 - 5·92) |
| Ngada | 163217 | 165314 | 167396 | 4 | 4 | 9 | 2 | 2 | 5 | 0·99 (0·25 - 3·95) | 2·19 (0·7 - 6·91) |
| Manggarai | 338424 | 342908 | 315041 | 8 | 28 | 22 | 2 | 8 | 7 | 3·45 (1·65 - 7·22) | 2·95 (1·37 - 6·39) |

| District | Number of populations | | | Number of reported mortalities | | | Mortality rate per 100,000 population | | | Mortality rate ratio for 2020 vs 2019 (95% CI) | Mortality rate ratio for 2021 vs 2019 (95% CI) |
|---|---|---|---|---|---|---|---|---|---|---|---|
| | 2019 | 2020 | 2021 | 2019 | 2020 | 2021 | 2019 | 2020 | 2021 | | |
| Rote Ndao | 172104 | 178805 | 145972 | 14 | 11 | 5 | 8 | 6 | 3 | 0·76 (0·34 - 1·66) | 0·42 (0·16 - 1·13) |
| Manggarai Barat | 274689 | 280412 | 259566 | 5 | 11 | 13 | 2 | 4 | 5 | 2·16 (0·77 - 6·05) | 2·75 (1·02 - 7·39) |
| Sumba Tengah | 72800 | 73820 | 87630 | 0 | 3 | 4 | 0 | 4 | 5 | NA | NA |
| Sumba Barat Daya | 344720 | 350923 | 305689 | 4 | 19 | 27 | 1 | 5 | 9 | 4·67 (1·75 - 12·41) | 7·61 (3·13 - 18·52) |
| Nagekeo | 145826 | 147189 | 162463 | 3 | 15 | 7 | 2 | 10 | 4 | 4·95 (1·62 - 15·13) | 2·09 (0·56 - 7·86) |
| Manggarai Timur | 287207 | 289836 | 277914 | 4 | 3 | 7 | 1 | 1 | 3 | 0·74 (0·17 - 3·3) | 1·81 (0·54 - 6·07) |
| Sabu Raijua | 97379 | 100684 | 90837 | 5 | 6 | 4 | 5 | 6 | 4 | 1·16 (0·35 - 3·8) | 0·86 (0·23 - 3·19) |
| Malaka | 191892 | 194776 | 185809 | 0 | 13 | 6 | 0 | 7 | 3 | NA | NA |
| Kota Kupang | 434972 | 446193 | 452626 | 25 | 28 | 7 | 6 | 6 | 2 | 1·09 (0·64 - 1·87) | 0·27 (0·12 - 0·59) |
| **West Kalimantan** | | | | | | | | | | | |
| Sambas | 535725 | 629905 | 637811 | 11 | 25 | 27 | 2 | 4 | 4 | 1·93 (0·96 - 3·88) | 2·06 (1·04 - 4·09) |
| Bengkayang | 255261 | 286366 | 290943 | 11 | 14 | 12 | 4 | 5 | 4 | 1·13 (0·52 - 2·5) | 0·96 (0·42 - 2·17) |
| Landak | 377305 | 397610 | 401103 | 12 | 14 | 6 | 3 | 4 | 1 | 1·11 (0·51 - 2·39) | 0·47 (0·18 - 1·22) |
| Mempawah | 264225 | 301560 | 305673 | 29 | 27 | 19 | 11 | 9 | 6 | 0·82 (0·48 - 1·38) | 0·57 (0·32 - 1) |
| Sanggau | 470224 | 484836 | 488527 | 22 | 35 | 31 | 5 | 7 | 6 | 1·54 (0·91 - 2·62) | 1·36 (0·79 - 2·34) |
| Ketapang | 512783 | 570657 | 579927 | 22 | 42 | 20 | 4 | 7 | 3 | 1·72 (1·03 - 2·86) | 0·8 (0·44 - 1·47) |
| Sintang | 418785 | 421306 | 423674 | 4 | 14 | 9 | 1 | 3 | 2 | 3·48 (1·23 - 9·87) | 2·22 (0·71 - 7) |
| Kapuas Hulu | 263207 | 252609 | 253740 | 8 | 8 | 9 | 3 | 3 | 4 | 1·04 (0·39 - 2·78) | 1·17 (0·45 - 3·02) |
| Sekadau | 201578 | 211559 | 212878 | 1 | 8 | 10 | 0 | 4 | 5 | 7·62 (1·31 - 44·32) | 9·47 (1·77 - 50·76) |
| Melawi | 208417 | 228270 | 231242 | 0 | 6 | 4 | 0 | 3 | 2 | NA | NA |
| Kayong Utara | 112715 | 126571 | 128550 | 6 | 0 | 4 | 5 | 0 | 3 | NA | 0·58 (0·17 - 2·04) |
| Kubu Raya | 579331 | 609392 | 615125 | 5 | 32 | 23 | 1 | 5 | 4 | 6·08 (2·66 - 13·89) | 4·33 (1·79 - 10·5) |
| Kota Pontianak | 646661 | 658685 | 663713 | 48 | 60 | 48 | 7 | 9 | 7 | 1·23 (0·84 - 1·79) | 0·97 (0·65 - 1·45) |
| Kota Singkawang | 222910 | 235064 | 237891 | 47 | 22 | 21 | 21 | 9 | 9 | 0·44 (0·27 - 0·73) | 0·42 (0·25 - 0·69) |
| **Central Kalimantan** | | | | | | | | | | | |

| District | Number of populations | | | Number of reported mortalities | | | Mortality rate per 100,000 population | | | Mortality rate ratio for 2020 vs 2019 (95% CI) | Mortality rate ratio for 2021 vs 2019 (95% CI) |
|---|---|---|---|---|---|---|---|---|---|---|---|
| | 2019 | 2020 | 2021 | 2019 | 2020 | 2021 | 2019 | 2020 | 2021 | | |
| Kotawaringin Barat | 312911 | 270388 | 272531 | 1 | 4 | 3 | 0 | 1 | 1 | 4·63 (0·63 - 33·89) | 3·44 (0·41 - 28·84) |
| Kotawaringin Timur | 466366 | 428895 | 432283 | 8 | 3 | 10 | 2 | 1 | 2 | 0·41 (0·11 - 1·47) | 1·35 (0·53 - 3·41) |
| Kapuas | 358820 | 410446 | 416181 | 28 | 19 | 24 | 8 | 5 | 6 | 0·59 (0·33 - 1·06) | 0·74 (0·43 - 1·27) |
| Barito Selatan | 136796 | 131140 | 131606 | 1 | 3 | 4 | 1 | 2 | 3 | 3·13 (0·37 - 26·73) | 4·16 (0·55 - 31·23) |
| Barito Utara | 130713 | 154812 | 157231 | 7 | 5 | 6 | 5 | 3 | 4 | 0·6 (0·19 - 1·88) | 0·71 (0·24 - 2·11) |
| Sukamara | 64342 | 63464 | 64941 | 2 | 0 | 2 | 3 | 0 | 3 | NA | 0·99 (0·14 - 7·03) |
| Lamandau | 82680 | 97611 | 100535 | 9 | 8 | 10 | 11 | 8 | 10 | 0·75 (0·29 - 1·95) | 0·91 (0·37 - 2·25) |
| Seruyan | 205880 | 162906 | 164378 | 8 | 5 | 13 | 4 | 3 | 8 | 0·79 (0·26 - 2·41) | 2·04 (0·86 - 4·82) |
| Katingan | 169997 | 162222 | 163099 | 0 | 5 | 7 | 0 | 3 | 4 | NA | NA |
| Pulang Pisau | 127910 | 134499 | 135336 | 9 | 3 | 0 | 7 | 2 | 0 | 0·32 (0·09 - 1·09) | NA |
| Gunung Mas | 119910 | 135373 | 138407 | 4 | 3 | 3 | 3 | 2 | 2 | 0·66 (0·15 - 2·94) | 0·65 (0·15 - 2·87) |
| Barito Timur | 126874 | 113229 | 114243 | 3 | 9 | 5 | 2 | 8 | 4 | 3·36 (0·98 - 11·5) | 1·85 (0·45 - 7·57) |
| Murung Raya | 120785 | 111527 | 112445 | 10 | 4 | 3 | 8 | 4 | 3 | 0·43 (0·14 - 1·34) | 0·32 (0·09 - 1·1) |
| Kota Palangka Raya | 291785 | 293457 | 298954 | 9 | 8 | 12 | 3 | 3 | 4 | 0·88 (0·34 - 2·29) | 1·3 (0·55 - 3·08) |
| **South Kalimantan** | | | | | | | | | | | |
| Tanah Laut | 343890 | 348966 | 354340 | 22 | 12 | 13 | 6 | 3 | 4 | 0·54 (0·27 - 1·07) | 0·57 (0·29 - 1·13) |
| Kotabaru | 342217 | 325622 | 329483 | 21 | 16 | 7 | 6 | 5 | 2 | 0·8 (0·42 - 1·53) | 0·35 (0·15 - 0·78) |
| Banjar | 588066 | 565635 | 572109 | 28 | 38 | 44 | 5 | 7 | 8 | 1·41 (0·87 - 2·29) | 1·62 (1·01 - 2·58) |
| Barito Kuala | 313595 | 313021 | 316963 | 7 | 8 | 6 | 2 | 3 | 2 | 1·14 (0·42 - 3·15) | 0·85 (0·29 - 2·52) |
| Tapin | 191372 | 189475 | 191801 | 16 | 12 | 7 | 8 | 6 | 4 | 0·76 (0·36 - 1·6) | 0·44 (0·18 - 1·03) |
| Hulu Sungai Selatan | 237702 | 228006 | 229960 | 6 | 13 | 11 | 3 | 6 | 5 | 2·26 (0·88 - 5·79) | 1·9 (0·71 - 5·04) |
| Hulu Sungai Tengah | 272419 | 258721 | 260754 | 35 | 26 | 33 | 13 | 10 | 13 | 0·78 (0·47 - 1·3) | 0·99 (0·61 - 1·58) |
| Hulu Sungai Utara | 237573 | 226727 | 228831 | 12 | 14 | 14 | 5 | 6 | 6 | 1·22 (0·57 - 2·64) | 1·21 (0·56 - 2·62) |
| Tabalong | 254322 | 253305 | 256903 | 12 | 14 | 10 | 5 | 6 | 4 | 1·17 (0·54 - 2·53) | 0·82 (0·36 - 1·91) |
| Tanah Bumbu | 360187 | 322646 | 328146 | 14 | 24 | 5 | 4 | 7 | 2 | 1·91 (1 - 3·66) | 0·39 (0·15 - 1·05) |

| District | Number of populations | | | Number of reported mortalities | | | Mortality rate per 100,000 population | | | Mortality rate ratio for 2020 vs 2019 (95% CI) | Mortality rate ratio for 2021 vs 2019 (95% CI) |
|---|---|---|---|---|---|---|---|---|---|---|---|
| | 2019 | 2020 | 2021 | 2019 | 2020 | 2021 | 2019 | 2020 | 2021 | | |
| Balangan | 131428 | 130355 | 132213 | 9 | 0 | 6 | 7 | 0 | 5 | NA | 0·66 (0·24 - 1·85) |
| Kota Banjarmasin | 708606 | 657663 | 662320 | 55 | 15 | 19 | 8 | 2 | 3 | 0·29 (0·17 - 0·5) | 0·37 (0·22 - 0·61) |
| Kota Banjar Baru | 262719 | 253442 | 258753 | 20 | 8 | 4 | 8 | 3 | 2 | 0·41 (0·19 - 0·92) | 0·2 (0·08 - 0·53) |
| **East Kalimantan** | | | | | | | | | | | |
| Paser | 285894 | 275452 | 277602 | 33 | 39 | 23 | 12 | 14 | 8 | 1·23 (0·77 - 1·95) | 0·72 (0·42 - 1·22) |
| Kutai Barat | 148020 | 172288 | 173982 | 5 | 6 | 13 | 3 | 3 | 7 | 1·03 (0·31 - 3·38) | 2·21 (0·81 - 6·04) |
| Kutai Kartanegara | 786122 | 729382 | 733626 | 35 | 32 | 24 | 4 | 4 | 3 | 0·99 (0·61 - 1·59) | 0·73 (0·44 - 1·23) |
| Kutai Timur | 376111 | 434459 | 449161 | 7 | 25 | 15 | 2 | 6 | 3 | 3·09 (1·4 - 6·85) | 1·79 (0·74 - 4·35) |
| Berau | 232287 | 248035 | 252648 | 17 | 20 | 8 | 7 | 8 | 3 | 1·1 (0·58 - 2·1) | 0·43 (0·19 - 0·98) |
| Penajam Paser Utara | 160912 | 178681 | 180657 | 11 | 11 | 8 | 7 | 6 | 4 | 0·9 (0·39 - 2·08) | 0·65 (0·26 - 1·6) |
| Mahakam Hulu | 26375 | 32513 | 32969 | 0 | 0 | 1 | 0 | 0 | 3 | NA | NA |
| Kota Balikpapan | 655178 | 688318 | 695287 | 24 | 45 | 30 | 4 | 7 | 4 | 1·78 (1·09 - 2·91) | 1·18 (0·69 - 2·01) |
| Kota Samarinda | 872768 | 827994 | 831460 | 61 | 55 | 71 | 7 | 7 | 9 | 0·95 (0·66 - 1·37) | 1·22 (0·87 - 1·72) |
| Kota Bontang | 177722 | 178917 | 180843 | 17 | 29 | 17 | 10 | 16 | 9 | 1·69 (0·94 - 3·06) | 0·98 (0·5 - 1·92) |
| **North Kalimantan** | | | | | | | | | | | |
| Malinau | 84609 | 82500 | 83800 | 4 | 7 | 4 | 5 | 8 | 5 | 1·79 (0·53 - 6·03) | 1·01 (0·25 - 4·04) |
| Bulungan | 133166 | 151800 | 154500 | 8 | 10 | 10 | 6 | 7 | 6 | 1·1 (0·43 - 2·78) | 1·08 (0·43 - 2·73) |
| Nunukan | 26607 | 25600 | 26400 | 6 | 8 | 2 | 23 | 31 | 8 | 1·39 (0·48 - 3·98) | 0·34 (0·07 - 1·54) |
| Tana Tidung | 196918 | 199100 | 203200 | 0 | 1 | 1 | 0 | 1 | 0 | NA | NA |
| Kota Tarakan | 254262 | 242800 | 245700 | 59 | 30 | 41 | 23 | 12 | 17 | 0·53 (0·35 - 0·82) | 0·72 (0·48 - 1·07) |
| **North Sulawesi** | | | | | | | | | | | |
| Bolaang Mongondow | 247811 | 248751 | 250478 | 13 | 29 | 23 | 5 | 12 | 9 | 2·22 (1·18 - 4·2) | 1·75 (0·89 - 3·43) |
| Minahasa | 341176 | 347290 | 348673 | 19 | 22 | 25 | 6 | 6 | 7 | 1·14 (0·62 - 2·1) | 1·29 (0·71 - 2·33) |
| Kepulauan Sangihe | 131163 | 139262 | 139684 | 5 | 2 | 2 | 4 | 1 | 1 | 0·38 (0·08 - 1·82) | 0·38 (0·08 - 1·82) |
| Kepulauan Talaud | 92475 | 94521 | 94983 | 1 | 12 | 4 | 1 | 13 | 4 | 11·74 (2·36 - 58·32) | 3·89 (0·51 - 29·69) |

| District | Number of populations | | | Number of reported mortalities | | | Mortality rate per 100,000 population | | | Mortality rate ratio for 2020 vs 2019 (95% CI) | Mortality rate ratio for 2021 vs 2019 (95% CI) |
|---|---|---|---|---|---|---|---|---|---|---|---|
| | 2019 | 2020 | 2021 | 2019 | 2020 | 2021 | 2019 | 2020 | 2021 | | |
| Minahasa Selatan | 210695 | 236463 | 238746 | 9 | 9 | 16 | 4 | 4 | 7 | 0·89 (0·35 - 2·24) | 1·57 (0·7 - 3·53) |
| Minahasa Utara | 203624 | 224993 | 226915 | 3 | 19 | 8 | 1 | 8 | 4 | 5·73 (1·95 - 16·81) | 2·39 (0·66 - 8·66) |
| Bolaang Mongondow Utara | 80313 | 83112 | 83743 | 0 | 9 | 11 | 0 | 11 | 13 | NA | NA |
| Kep, Siau Tagulandang Biaro | 66403 | 71817 | 72135 | 5 | 5 | 7 | 8 | 7 | 10 | 0·92 (0·27 - 3·19) | 1·29 (0·41 - 4·05) |
| Minahasa Tenggara | 106899 | 116323 | 117079 | 16 | 11 | 11 | 15 | 9 | 9 | 0·63 (0·3 - 1·35) | 0·63 (0·29 - 1·34) |
| Bolaang Mongondow Selatan | 66071 | 69791 | 70529 | 2 | 2 | 0 | 3 | 3 | 0 | 0·95 (0·13 - 6·72) | NA |
| Bolaang Mongondow Timur | 72408 | 88241 | 89981 | 11 | 9 | 10 | 15 | 10 | 11 | 0·67 (0·28 - 1·61) | 0·73 (0·31 - 1·72) |
| Kota Manado | 433635 | 451916 | 453182 | 1 | 16 | 12 | 0 | 4 | 3 | 15·35 (3·4 - 69·25) | 11·48 (2·3 - 57·42) |
| Kota Bitung | 219004 | 225134 | 227177 | 19 | 14 | 23 | 9 | 6 | 10 | 0·72 (0·36 - 1·43) | 1·17 (0·64 - 2·14) |
| Kota Tomohon | 106917 | 100587 | 100853 | 13 | 26 | 14 | 12 | 26 | 14 | 2·13 (1·11 - 4·07) | 1·14 (0·54 - 2·43) |
| Kota Kotamobagu | 128387 | 123722 | 124473 | 7 | 6 | 7 | 5 | 5 | 6 | 0·89 (0·3 - 2·64) | 1·03 (0·36 - 2·94) |
| **Central Sulawesi** | | | | | | | | | | | |
| Banggai Kepulauan | 118401 | 120142 | 121680 | 9 | 15 | 7 | 8 | 12 | 6 | 1·64 (0·72 - 3·72) | 0·76 (0·28 - 2·03) |
| Banggai | 376808 | 362275 | 366220 | 41 | 51 | 37 | 11 | 14 | 10 | 1·29 (0·86 - 1·95) | 0·93 (0·6 - 1·45) |
| Morowali | 121296 | 161727 | 167910 | 10 | 16 | 7 | 8 | 10 | 4 | 1·2 (0·55 - 2·64) | 0·51 (0·2 - 1·3) |
| Poso | 256393 | 244875 | 248350 | 8 | 18 | 6 | 3 | 7 | 2 | 2·36 (1·05 - 5·28) | 0·77 (0·27 - 2·23) |
| Donggala | 304110 | 300436 | 302910 | 11 | 9 | 16 | 4 | 3 | 5 | 0·83 (0·34 - 2) | 1·46 (0·68 - 3·13) |
| Toli-Toli | 235800 | 225154 | 226800 | 8 | 9 | 21 | 3 | 4 | 9 | 1·18 (0·46 - 3·05) | 2·73 (1·25 - 5·96) |
| Buol | 162179 | 145254 | 146630 | 2 | 7 | 5 | 1 | 5 | 3 | 3·91 (0·91 - 16·76) | 2·77 (0·57 - 13·31) |
| Parigi Moutong | 490915 | 440015 | 443170 | 36 | 45 | 35 | 7 | 10 | 8 | 1·39 (0·9 - 2·16) | 1·08 (0·68 - 1·71) |
| Tojo Una-Una | 153991 | 163829 | 166340 | 6 | 7 | 7 | 4 | 4 | 4 | 1·1 (0·37 - 3·26) | 1·08 (0·36 - 3·21) |
| Sigi | 239421 | 257585 | 261680 | 22 | 30 | 25 | 9 | 12 | 10 | 1·27 (0·73 - 2·19) | 1·04 (0·59 - 1·84) |
| Banggai Laut | 75003 | 70435 | 70870 | 4 | 9 | 13 | 5 | 13 | 18 | 2·4 (0·77 - 7·5) | 3·44 (1·2 - 9·85) |
| Morowali Utara | 128323 | 120789 | 122240 | 1 | 4 | 6 | 1 | 3 | 5 | 4·25 (0·57 - 31·75) | 6·3 (0·99 - 39·87) |
| Kota Palu | 391383 | 373218 | 377030 | 9 | 20 | 46 | 2 | 5 | 12 | 2·33 (1·09 - 5) | 5·31 (2·8 - 10·04) |

| District | Number of populations | | | Number of reported mortalities | | | Mortality rate per 100,000 population | | | Mortality rate ratio for 2020 vs 2019 (95% CI) | Mortality rate ratio for 2021 vs 2019 (95% CI) |
|---|---|---|---|---|---|---|---|---|---|---|---|
| | 2019 | 2020 | 2021 | 2019 | 2020 | 2021 | 2019 | 2020 | 2021 | | |
| **South Sulawesi** | | | | | | | | | | | |
| Kepulauan Selayar | 135624 | 136871 | 136118 | 8 | 14 | 9 | 6 | 10 | 7 | 1·73 (0·74 - 4·09) | 1·12 (0·43 - 2·9) |
| Bulukumba | 420603 | 423012 | 421959 | 31 | 20 | 34 | 7 | 5 | 8 | 0·64 (0·37 - 1·12) | 1·09 (0·67 - 1·78) |
| Bantaeng | 187626 | 188495 | 189202 | 14 | 20 | 22 | 7 | 11 | 12 | 1·42 (0·72 - 2·81) | 1·56 (0·8 - 3·03) |
| Jeneponto | 363792 | 365610 | 367160 | 14 | 18 | 25 | 4 | 5 | 7 | 1·28 (0·64 - 2·57) | 1·77 (0·93 - 3·37) |
| Takalar | 298688 | 301424 | 298717 | 24 | 23 | 24 | 8 | 8 | 8 | 0·95 (0·54 - 1·68) | 1 (0·57 - 1·76) |
| Gowa | 772684 | 784511 | 780138 | 14 | 55 | 72 | 2 | 7 | 9 | 3·87 (2·25 - 6·67) | 5·09 (3·05 - 8·52) |
| Sinjai | 244125 | 245389 | 245501 | 36 | 24 | 19 | 15 | 10 | 8 | 0·66 (0·4 - 1·11) | 0·52 (0·3 - 0·91) |
| Maros | 353121 | 356195 | 357320 | 40 | 37 | 25 | 11 | 10 | 7 | 0·92 (0·59 - 1·43) | 0·62 (0·38 - 1·01) |
| Pangkajene Dan Kepulauan | 335514 | 338219 | 339575 | 56 | 36 | 52 | 17 | 11 | 15 | 0·64 (0·42 - 0·97) | 0·92 (0·63 - 1·34) |
| Barru | 174323 | 174989 | 175023 | 1 | 17 | 17 | 1 | 10 | 10 | 16·94 (3·88 - 73·92) | 16·93 (3·88 - 73·91) |
| Bone | 758589 | 762073 | 757741 | 41 | 55 | 62 | 5 | 7 | 8 | 1·34 (0·89 - 2) | 1·51 (1·02 - 2·24) |
| Soppeng | 226991 | 227208 | 229539 | 1 | 8 | 9 | 0 | 4 | 4 | 7·99 (1·39 - 45·85) | 8·9 (1·62 - 48·99) |
| Wajo | 397814 | 398784 | 406091 | 54 | 38 | 31 | 14 | 10 | 8 | 0·7 (0·46 - 1·06) | 0·56 (0·36 - 0·87) |
| Sidenreng Rappang | 301972 | 304826 | 302918 | 30 | 21 | 25 | 10 | 7 | 8 | 0·69 (0·4 - 1·21) | 0·83 (0·49 - 1·41) |
| Pinrang | 377119 | 379402 | 381114 | 41 | 22 | 31 | 11 | 6 | 8 | 0·53 (0·32 - 0·89) | 0·75 (0·47 - 1·19) |
| Enrekang | 206387 | 207800 | 209974 | 5 | 14 | 16 | 2 | 7 | 8 | 2·78 (1·05 - 7·39) | 3·15 (1·21 - 8·14) |
| Luwu | 362027 | 364680 | 369924 | 23 | 26 | 44 | 6 | 7 | 12 | 1·12 (0·64 - 1·97) | 1·87 (1·14 - 3·07) |
| Tana Toraja | 234002 | 235103 | 239516 | 3 | 7 | 10 | 1 | 3 | 4 | 2·32 (0·62 - 8·64) | 3·26 (0·96 - 11·01) |
| Luwu Utara | 312883 | 315202 | 318064 | 5 | 36 | 26 | 2 | 11 | 8 | 7·15 (3·21 - 15·91) | 5·12 (2·17 - 12·07) |
| Luwu Timur | 299673 | 305407 | 303479 | 5 | 12 | 10 | 2 | 4 | 3 | 2·35 (0·86 - 6·48) | 1·97 (0·69 - 5·66) |
| Toraja Utara | 231214 | 232394 | 237259 | 4 | 7 | 14 | 2 | 3 | 6 | 1·74 (0·52 - 5·86) | 3·41 (1·2 - 9·69) |
| Kota Makassar | 1526677 | 1545373 | 1555088 | 241 | 154 | 141 | 16 | 10 | 9 | 0·63 (0·52 - 0·77) | 0·57 (0·47 - 0·71) |
| Kota Parepare | 145178 | 146714 | 147090 | 28 | 19 | 16 | 19 | 13 | 11 | 0·67 (0·38 - 1·2) | 0·56 (0·31 - 1·03) |
| Kota Palopo | 184614 | 188323 | 187671 | 13 | 13 | 15 | 7 | 7 | 8 | 0·98 (0·45 - 2·11) | 1·14 (0·54 - 2·38) |

| District | Number of populations | | | Number of reported mortalities | | | Mortality rate per 100,000 population | | | Mortality rate ratio for 2020 vs 2019 (95% CI) | Mortality rate ratio for 2021 vs 2019 (95% CI) |
|---|---|---|---|---|---|---|---|---|---|---|---|
| | 2019 | 2020 | 2021 | 2019 | 2020 | 2021 | 2019 | 2020 | 2021 | | |
| **Southeast Sulawesi** | | | | | | | | | | | |
| Buton | 102641 | 103869 | 117040 | 10 | 7 | 7 | 10 | 7 | 6 | 0·69 (0·26 - 1·81) | 0·61 (0·24 - 1·6) |
| Muna | 224099 | 227289 | 218956 | 4 | 4 | 6 | 2 | 2 | 3 | 0·99 (0·25 - 3·94) | 1·54 (0·44 - 5·39) |
| Konawe | 254695 | 260411 | 261116 | 3 | 15 | 10 | 1 | 6 | 4 | 4·89 (1·6 - 14·96) | 3·25 (0·96 - 11) |
| Kolaka | 261664 | 266069 | 241366 | 20 | 29 | 17 | 8 | 11 | 7 | 1·43 (0·81 - 2·51) | 0·92 (0·48 - 1·76) |
| Konawe Selatan | 314785 | 319291 | 312674 | 27 | 32 | 25 | 9 | 10 | 8 | 1·17 (0·7 - 1·95) | 0·93 (0·54 - 1·61) |
| Bombana | 184570 | 189269 | 151910 | 19 | 19 | 12 | 10 | 10 | 8 | 0·98 (0·52 - 1·84) | 0·77 (0·37 - 1·58) |
| Wakatobi | 95892 | 96111 | 113122 | 3 | 2 | 1 | 3 | 2 | 1 | 0·67 (0·11 - 3·93) | 0·28 (0·03 - 2·35) |
| Kolaka Utara | 150831 | 153669 | 139234 | 4 | 9 | 9 | 3 | 6 | 6 | 2·21 (0·7 - 6·96) | 2·44 (0·78 - 7·62) |
| Buton Utara | 64072 | 64993 | 67714 | 0 | 2 | 3 | 0 | 3 | 4 | NA | NA |
| Konawe Utara | 63814 | 65183 | 68950 | 4 | 10 | 3 | 6 | 15 | 4 | 2·45 (0·8 - 7·51) | 0·69 (0·16 - 3·08) |
| Kolaka Timur | 133324 | 135569 | 120966 | 12 | 5 | 8 | 9 | 4 | 7 | 0·41 (0·15 - 1·12) | 0·73 (0·3 - 1·79) |
| Konawe Kepulauan | 34219 | 34666 | 37639 | 1 | 1 | 2 | 3 | 3 | 5 | 0·99 (0·06 - 15·78) | 1·82 (0·17 - 19·36) |
| Muna Barat | 81624 | 82785 | 84777 | 3 | 9 | 12 | 4 | 11 | 14 | 2·96 (0·85 - 10·27) | 3·85 (1·19 - 12·46) |
| Buton Tengah | 93091 | 94207 | 116599 | 10 | 5 | 3 | 11 | 5 | 3 | 0·49 (0·17 - 1·41) | 0·24 (0·07 - 0·78) |
| Buton Selatan | 80784 | 81752 | 95472 | 0 | 6 | 3 | 0 | 7 | 3 | NA | NA |
| Kota Kendari | 392830 | 404232 | 350267 | 21 | 10 | 20 | 5 | 2 | 6 | 0·46 (0·22 - 0·96) | 1·07 (0·58 - 1·97) |
| Kota Baubau | 171802 | 176224 | 161354 | 12 | 26 | 24 | 7 | 15 | 15 | 2·11 (1·08 - 4·12) | 2·13 (1·08 - 4·19) |
| **Gorontalo** | | | | | | | | | | | |
| Boalemo | 167024 | 145868 | 147038 | 8 | 17 | 15 | 5 | 12 | 10 | 2·43 (1·08 - 5·49) | 2·13 (0·92 - 4·92) |
| Gorontalo | 378527 | 393107 | 395635 | 0 | 17 | 32 | 0 | 4 | 8 | NA | NA |
| Pohuwato | 161373 | 146432 | 147689 | 0 | 12 | 8 | 0 | 8 | 5 | NA | NA |
| Bone Bolango | 161236 | 162778 | 164277 | 24 | 28 | 21 | 15 | 17 | 13 | 1·16 (0·67 - 1·99) | 0·86 (0·48 - 1·54) |
| Gorontalo Utara | 115072 | 124957 | 126521 | 11 | 13 | 11 | 10 | 10 | 9 | 1·09 (0·49 - 2·43) | 0·91 (0·39 - 2·1) |
| Kota Gorontalo | 219399 | 198539 | 199788 | 15 | 10 | 6 | 7 | 5 | 3 | 0·74 (0·33 - 1·63) | 0·44 (0·17 - 1·1) |

| District | Number of populations | | | Number of reported mortalities | | | Mortality rate per 100,000 population | | | Mortality rate ratio for 2020 vs 2019 (95% CI) | Mortality rate ratio for 2021 vs 2019 (95% CI) |
|---|---|---|---|---|---|---|---|---|---|---|---|
| | 2019 | 2020 | 2021 | 2019 | 2020 | 2021 | 2019 | 2020 | 2021 | | |
| **West Sulawesi** | | | | | | | | | | | |
| Majene | 173884 | 174407 | 175790 | 17 | 22 | 11 | 10 | 13 | 6 | 1·29 (0·69 - 2·43) | 0·64 (0·3 - 1·36) |
| Polewali Mandar | 442576 | 478534 | 483920 | 18 | 33 | 29 | 4 | 7 | 6 | 1·7 (0·96 - 2·99) | 1·47 (0·82 - 2·64) |
| Mamasa | 161971 | 163383 | 164800 | 3 | 4 | 3 | 2 | 2 | 2 | 1·32 (0·3 - 5·88) | 0·98 (0·2 - 4·87) |
| Mamuju | 293326 | 278764 | 281850 | 30 | 23 | 20 | 10 | 8 | 7 | 0·81 (0·47 - 1·39) | 0·69 (0·4 - 1·22) |
| Pasangkayu | 174471 | 188861 | 193100 | 12 | 11 | 8 | 7 | 6 | 4 | 0·85 (0·37 - 1·92) | 0·6 (0·25 - 1·46) |
| Mamuju Tengah | 134028 | 135280 | 137380 | 15 | 7 | 9 | 11 | 5 | 7 | 0·46 (2·05 - 10·45) | 0·59 (0·26 - 1·32) |
| **Maluku** | | | | | | | | | | | |
| Maluku Tenggara Barat | 113012 | 123572 | 124075 | 2 | 9 | 5 | 2 | 7 | 4 | 4·12 (1 - 16·88) | 2·28 (0·46 - 11·22) |
| Maluku Tenggara | 99790 | 121511 | 122640 | 1 | 0 | 2 | 1 | 0 | 2 | NA | 1·63 (0·15 - 17·53) |
| Maluku Tengah | 373378 | 423094 | 424730 | 1 | 10 | 14 | 0 | 2 | 3 | 8·82 (1·61 - 48·28) | 12·31 (2·53 - 59·75) |
| Buru | 143688 | 135238 | 136393 | 1 | 8 | 9 | 1 | 6 | 7 | 8·5 (1·51 - 47·92) | 9·48 (1·75 - 51·24) |
| Kepulauan Aru | 96114 | 102237 | 102916 | 0 | 4 | 0 | 0 | 4 | 0 | NA | NA |
| Seram Bagian Barat | 171586 | 212393 | 214733 | 7 | 11 | 5 | 4 | 5 | 2 | 1·27 (0·49 - 3·27) | 0·57 (0·18 - 1·77) |
| Seram Bagian Timur | 114677 | 137972 | 140271 | 0 | 4 | 4 | 0 | 3 | 3 | NA | NA |
| Maluku Barat Daya | 73103 | 81928 | 82187 | 0 | 3 | 1 | 0 | 4 | 1 | NA | NA |
| Buru Selatan | 63328 | 75410 | 76715 | 3 | 0 | 3 | 5 | 0 | 4 | NA | 0·83 (0·17 - 4·08) |
| Kota Ambon | 478616 | 347288 | 347644 | 31 | 23 | 22 | 6 | 7 | 6 | 1·02 (0·6 - 1·75) | 0·98 (0·57 - 1·69) |
| Kota Tual | 75578 | 88280 | 90322 | 1 | 8 | 1 | 1 | 9 | 1 | 6·85 (1·14 - 41·01) | 0·84 (0·05 - 13·33) |
| **North Maluku** | | | | | | | | | | | |
| Halmahera Barat | 118287 | 132349 | 134630 | 15 | 15 | 19 | 13 | 11 | 14 | 0·89 (0·44 - 1·83) | 1·11 (0·57 - 2·19) |
| Halmahera Tengah | 55728 | 56802 | 57809 | 6 | 4 | 0 | 11 | 7 | 0 | 0·65 (0·19 - 2·3) | NA |
| Kepulauan Sula | 102886 | 104082 | 105293 | 11 | 6 | 4 | 11 | 6 | 4 | 0·54 (0·2 - 1·44) | 0·36 (0·12 - 1·06) |
| Halmahera Selatan | 235090 | 248395 | 251690 | 26 | 6 | 20 | 11 | 2 | 8 | 0·22 (0·1 - 0·49) | 0·72 (0·4 - 1·28) |
| Halmahera Utara | 193851 | 197638 | 199936 | 25 | 18 | 13 | 13 | 9 | 7 | 0·71 (0·39 - 1·29) | 0·5 (0·26 - 0·97) |

| District | Number of populations | | | Number of reported mortalities | | | Mortality rate per 100,000 population | | | Mortality rate ratio for 2020 vs 2019 (95% CI) | Mortality rate ratio for 2021 vs 2019 (95% CI) |
|---|---|---|---|---|---|---|---|---|---|---|---|
| | 2019 | 2020 | 2021 | 2019 | 2020 | 2021 | 2019 | 2020 | 2021 | | |
| Halmahera Timur | 95005 | 91707 | 92954 | 6 | 2 | 4 | 6 | 2 | 4 | 0·35 (0·07 - 1·59) | 0·68 (0·19 - 2·4) |
| Pulau Morotai | 67284 | 74436 | 76102 | 4 | 4 | 5 | 6 | 5 | 7 | 0·9 (0·23 - 3·61) | 1·11 (0·3 - 4·11) |
| Pulau Taliabu | 53018 | 58047 | 58744 | 1 | 2 | 3 | 2 | 3 | 5 | 1·83 (0·17 - 19·43) | 2·71 (0·31 - 23·77) |
| Kota Ternate | 233208 | 205001 | 205870 | 23 | 16 | 19 | 10 | 8 | 9 | 0·79 (0·42 - 1·5) | 0·94 (0·51 - 1·72) |
| Kota Tidore Kepulauan | 101414 | 114480 | 116149 | 20 | 11 | 7 | 20 | 10 | 6 | 0·49 (0·24 - 1) | 0·31 (0·14 - 0·69) |
| **West Papua** | | | | | | | | | | | |
| Fakfak | 78686 | 85197 | 85817 | 11 | 12 | 17 | 14 | 14 | 20 | 1·01 (0·44 - 2·28) | 1·42 (0·67 - 3·01) |
| Kaimana | 60216 | 62256 | 62957 | 5 | 9 | 1 | 8 | 14 | 2 | 1·74 (0·59 - 5·12) | 0·19 (0·03 - 1·31) |
| Teluk Wondama | 32521 | 41644 | 42609 | 1 | 5 | 4 | 3 | 12 | 9 | 3·9 (0·53 - 28·55) | 3·05 (0·38 - 24·48) |
| Teluk Bintuni | 64406 | 87083 | 89418 | 19 | 5 | 7 | 30 | 6 | 8 | 0·19 (0·08 - 0·47) | 0·27 (0·12 - 0·59) |
| Manokwari | 175178 | 192663 | 194905 | 28 | 17 | 34 | 16 | 9 | 17 | 0·55 (0·3 - 1) | 1·09 (0·66 - 1·8) |
| Sorong Selatan | 46922 | 52469 | 53167 | 3 | 0 | 0 | 6 | 0 | 0 | NA | NA |
| Sorong | 88927 | 118679 | 121963 | 18 | 13 | 26 | 20 | 11 | 21 | 0·54 (0·27 - 1·09) | 1·05 (0·58 - 1·92) |
| Raja Ampat | 48493 | 64141 | 65403 | 1 | 2 | 2 | 2 | 3 | 3 | 1·51 (0·14 - 16·39) | 1·48 (0·14 - 16·1) |
| Tambrauw | 13879 | 28379 | 31385 | 0 | 0 | 0 | 0 | 0 | 0 | NA | NA |
| Maybrat | 40899 | 42991 | 43364 | 0 | 0 | 0 | 0 | 0 | 0 | NA | NA |
| Manokwari Selatan | 24220 | 35949 | 37149 | 1 | 2 | 3 | 4 | 6 | 8 | 1·35 (0·12 - 14·73) | 1·96 (0·21 - 18·03) |
| Pegunungan Arfak | 30976 | 38207 | 38936 | 0 | 0 | 0 | 0 | 0 | 0 | NA | NA |
| Kota Sorong | 254294 | 284410 | 289767 | 15 | 7 | 4 | 6 | 2 | 1 | 0·42 (0·17 - 1) | 0·23 (0·09 - 0·64) |
| **Papua** | | | | | | | | | | | |
| Merauke | 227411 | 230932 | 231696 | 40 | 37 | 21 | 18 | 16 | 9 | 0·91 (0·58 - 1·42) | 0·52 (0·31 - 0·87) |
| Jayawijaya | 217887 | 269553 | 273291 | 4 | 7 | 6 | 2 | 3 | 2 | 1·41 (0·42 - 4·8) | 1·2 (0·34 - 4·23) |
| Jayapura | 131802 | 166171 | 168476 | 58 | 34 | 24 | 44 | 20 | 14 | 0·46 (0·31 - 0·7) | 0·32 (0·21 - 0·51) |
| Nabire | 150308 | 169136 | 170914 | 41 | 31 | 27 | 27 | 18 | 16 | 0·67 (0·42 - 1·07) | 0·58 (0·36 - 0·94) |
| Kepulauan Yapen | 101204 | 112676 | 114210 | 13 | 3 | 1 | 13 | 3 | 1 | 0·21 (0·07 - 0·64) | 0·07 (0·01 - 0·31) |

| District | Number of populations | | | Number of reported mortalities | | | Mortality rate per 100,000 population | | | Mortality rate ratio for 2020 vs 2019 (95% CI) | Mortality rate ratio for 2021 vs 2019 (95% CI) |
|---|---|---|---|---|---|---|---|---|---|---|---|
| | 2019 | 2020 | 2021 | 2019 | 2020 | 2021 | 2019 | 2020 | 2021 | | |
| Biak Numfor | 152401 | 134650 | 135231 | 47 | 27 | 31 | 31 | 20 | 23 | 0·65 (0·41 - 1·04) | 0·74 (0·47 - 1·17) |
| Paniai | 177410 | 220410 | 223467 | 10 | 4 | 9 | 6 | 2 | 4 | 0·32 (0·11 - 0·97) | 0·71 (0·29 - 1·75) |
| Puncak Jaya | 129300 | 224527 | 227641 | 0 | 0 | 0 | 0 | 0 | 0 | NA | NA |
| Mimika | 219689 | 311969 | 316295 | 84 | 84 | 57 | 38 | 27 | 18 | 0·7 (0·52 - 0·95) | 0·47 (0·34 - 0·65) |
| Boven Digoel | 69211 | 64285 | 64716 | 13 | 7 | 5 | 19 | 11 | 8 | 0·58 (0·23 - 1·44) | 0·41 (0·15 - 1·12) |
| Mappi | 103292 | 108295 | 109579 | 33 | 21 | 24 | 32 | 19 | 22 | 0·61 (0·35 - 1·04) | 0·69 (0·41 - 1·16) |
| Asmat | 97490 | 110105 | 111632 | 22 | 38 | 13 | 23 | 35 | 12 | 1·53 (0·91 - 2·58) | 0·52 (0·26 - 1·01) |
| Yahukimo | 190887 | 350880 | 355746 | 0 | 0 | 0 | 0 | 0 | 0 | NA | NA |
| Pegunungan Bintang | 75788 | 77873 | 78178 | 0 | 2 | 1 | 0 | 3 | 1 | NA | NA |
| Tolikara | 139111 | 236986 | 240272 | 0 | 0 | 0 | 0 | 0 | 0 | NA | NA |
| Sarmi | 40515 | 41415 | 41849 | 0 | 3 | 2 | 0 | 7 | 5 | NA | NA |
| Keerom | 57100 | 61623 | 62157 | 5 | 5 | 12 | 9 | 8 | 19 | 0·93 (0·27 - 3·2) | 2·2 (0·8 - 6·09) |
| Waropen | 31514 | 33943 | 34414 | 6 | 4 | 0 | 19 | 12 | 0 | 0·62 (0·18 - 2·17) | NA |
| Supiori | 20710 | 22547 | 22860 | 5 | 1 | 3 | 24 | 4 | 13 | 0·18 (0·03 - 1·24) | 0·54 (0·13 - 2·23) |
| Mamberamo Raya | 24086 | 36483 | 36989 | 0 | 0 | 0 | 0 | 0 | 0 | NA | NA |
| Nduga | 98595 | 106533 | 107921 | 0 | 0 | 0 | 0 | 0 | 0 | NA | NA |
| Lanny Jaya | 178995 | 196339 | 198686 | 1 | 0 | 0 | 1 | 0 | 0 | NA | NA |
| Mamberamo Tengah | 48201 | 50685 | 51160 | 0 | 0 | 0 | 0 | 0 | 0 | NA | NA |
| Yalimo | 62605 | 101973 | 103387 | 0 | 2 | 1 | 0 | 2 | 1 | NA | NA |
| Puncak | 113204 | 114741 | 115474 | 0 | 0 | 0 | 0 | 0 | 0 | NA | NA |
| Dogiyai | 97902 | 116206 | 117818 | 0 | 0 | 0 | 0 | 0 | 0 | NA | NA |
| Intan Jaya | 49293 | 135043 | 136916 | 0 | 0 | 0 | 0 | 0 | 0 | NA | NA |
| Deiyai | 73199 | 99091 | 100466 | 0 | 0 | 0 | 0 | 0 | 0 | NA | NA |
| Kota Jayapura | 300192 | 398478 | 404004 | 40 | 40 | 31 | 13 | 10 | 8 | 0·75 (0·49 - 1·17) | 0·58 (0·36 - 0·92) |

**Supplementary table 3· Number of reported new TB cases, reported TB cases on treatment, TB treatment coverage rate, and TB treatment coverage rate ratio in 2020 and 2021 (pandemic), compared to in 2019 (pre-pandemic)**

| District | Number of cases | | | Number of cases on treatments | | | Treatment coverage | | | Treatment coverage ratio for 2020 vs 2019 (95% CI) | Treatment coverage ratio for 2021 vs 2019 (95% CI) |
|---|---|---|---|---|---|---|---|---|---|---|---|
| | 2019 | 2020 | 2021 | 2019 | 2020 | 2021 | 2019 | 2020 | 2021 | | |
| **Aceh** | | | | | | | | | | | |
| Simeulue | 124 | 166 | 118 | 124 | 163 | 80 | 100% | 98% | 68% | 0·98 (0·78-1·24) | 0·68 (0·51-0·9) |
| Aceh Singkil | 210 | 145 | 156 | 210 | 143 | 68 | 100% | 99% | 44% | 0·99 (0·8-1·22) | 0·44 (0·33-0·57) |
| Aceh Selatan | 375 | 349 | 533 | 375 | 307 | 401 | 100% | 88% | 75% | 0·88 (0·76-1·02) | 0·75 (0·65-0·87) |
| Aceh Tenggara | 130 | 126 | 347 | 130 | 123 | 164 | 100% | 98% | 47% | 0·98 (0·76-1·25) | 0·47 (0·38-0·59) |
| Aceh Timur | 567 | 613 | 639 | 567 | 579 | 615 | 100% | 94% | 96% | 0·94 (0·84-1·06) | 0·96 (0·86-1·08) |
| Aceh Tengah | 179 | 193 | 232 | 179 | 193 | 150 | 100% | 100% | 65% | 1 (0-0) | 0·65 (0·52-0·8) |
| Aceh Barat | 315 | 288 | 279 | 315 | 265 | 187 | 100% | 92% | 67% | 0·92 (0·78-1·08) | 0·67 (0·56-0·8) |
| Aceh Besar | 404 | 363 | 375 | 404 | 351 | 354 | 100% | 97% | 94% | 0·97 (0·84-1·12) | 0·94 (0·82-1·09) |
| Pidie | 814 | 673 | 773 | 814 | 601 | 718 | 100% | 89% | 93% | 0·89 (0·8-0·99) | 0·93 (0·84-1·03) |
| Bireuen | 859 | 757 | 820 | 859 | 751 | 738 | 100% | 99% | 90% | 0·99 (0·9-1·09) | 0·9 (0·82-0·99) |
| Aceh Utara | 909 | 970 | 763 | 909 | 957 | 661 | 100% | 99% | 87% | 0·99 (0·9-1·08) | 0·87 (0·78-0·96) |
| Aceh Barat Daya | 213 | 286 | 372 | 213 | 240 | 257 | 100% | 84% | 69% | 0·84 (0·7-1·01) | 0·69 (0·58-0·83) |
| Gayo Lues | 198 | 107 | 198 | 198 | 107 | 149 | 100% | 100% | 75% | 1 (0-0) | 0·75 (0·61-0·93) |
| Aceh Tamiang | 426 | 298 | 471 | 426 | 264 | 434 | 100% | 89% | 92% | 0·89 (0·76-1·03) | 0·92 (0·81-1·05) |
| Nagan Raya | 285 | 163 | 249 | 285 | 159 | 181 | 100% | 98% | 73% | 0·98 (0·8-1·18) | 0·73 (0·6-0·88) |
| Aceh Jaya | 213 | 161 | 164 | 213 | 158 | 120 | 100% | 98% | 73% | 0·98 (0·8-1·21) | 0·73 (0·59-0·91) |
| Bener Meriah | 71 | 109 | 95 | 71 | 101 | 62 | 100% | 93% | 65% | 0·93 (0·68-1·26) | 0·65 (0·47-0·92) |
| Pidie Jaya | 199 | 78 | 202 | 199 | 65 | 87 | 100% | 83% | 43% | 0·83 (0·63-1·1) | 0·43 (0·34-0·55) |
| Kota Banda Aceh | 846 | 510 | 786 | 846 | 398 | 593 | 100% | 78% | 75% | 0·78 (0·69-0·88) | 0·75 (0·68-0·84) |
| Kota Sabang | 26 | 29 | 17 | 26 | 29 | 11 | 100% | 100% | 65% | 1 (0-0) | 0·65 (0·32-1·3) |
| Kota Langsa | 412 | 298 | 360 | 412 | 265 | 301 | 100% | 89% | 84% | 0·89 (0·76-1·04) | 0·84 (0·72-0·97) |
| Kota Lhokseumawe | 593 | 317 | 672 | 593 | 300 | 569 | 100% | 95% | 85% | 0·95 (0·82-1·09) | 0·85 (0·75-0·95) |

| District | Number of cases | | | Number of cases on treatments | | | Treatment coverage | | | Treatment coverage ratio for 2020 vs 2019 (95% CI) | Treatment coverage ratio for 2021 vs 2019 (95% CI) |
|---|---|---|---|---|---|---|---|---|---|---|---|
| | 2019 | 2020 | 2021 | 2019 | 2020 | 2021 | 2019 | 2020 | 2021 | | |
| Kota Subulussalam | 176 | 227 | 247 | 176 | 207 | 195 | 100% | 91% | 79% | 0·91 (0·75-1·11) | 0·79 (0·64-0·97) |
| **North Sumatera** | | | | | | | | | | | |
| Nias | 300 | 205 | 436 | 300 | 98 | 239 | 100% | 48% | 55% | 0·48 (0·38-0·6) | 0·55 (0·46-0·65) |
| Mandailing Natal | 881 | 854 | 800 | 881 | 854 | 800 | 100% | 100% | 100% | 1 (0-0) | 1 (0-0) |
| Tapanuli Selatan | 656 | 452 | 402 | 656 | 448 | 369 | 100% | 99% | 92% | 0·99 (0·88-1·12) | 0·92 (0·81-1·04) |
| Tapanuli Tengah | 776 | 573 | 574 | 776 | 506 | 574 | 100% | 88% | 100% | 0·88 (0·79-0·99) | 1 (0-0) |
| Tapanuli Utara | 641 | 314 | 449 | 641 | 297 | 407 | 100% | 95% | 91% | 0·95 (0·82-1·09) | 0·91 (0·8-1·03) |
| Toba Samosir | 457 | 261 | 286 | 457 | 261 | 269 | 100% | 100% | 94% | 1 (0-0) | 0·94 (0·81-1·09) |
| Labuhan Batu | 1625 | 845 | 864 | 1625 | 794 | 711 | 100% | 94% | 82% | 0·94 (0·86-1·02) | 0·82 (0·75-0·9) |
| Asahan | 987 | 815 | 954 | 987 | 807 | 850 | 100% | 99% | 89% | 0·99 (0·9-1·09) | 0·89 (0·81-0·98) |
| Simalungun | 1733 | 1566 | 1444 | 1733 | 1523 | 1363 | 100% | 97% | 94% | 0·97 (0·91-1·04) | 0·94 (0·88-1·01) |
| Dairi | 577 | 485 | 482 | 577 | 485 | 473 | 100% | 100% | 98% | 1 (0-0) | 0·98 (0·87-1·11) |
| Karo | 824 | 597 | 577 | 824 | 559 | 483 | 100% | 94% | 84% | 0·94 (0·84-1·04) | 0·84 (0·75-0·94) |
| Deli Serdang | 3415 | 2794 | 3448 | 3415 | 2748 | 3098 | 100% | 98% | 90% | 0·98 (0·94-1·03) | 0·9 (0·86-0·94) |
| Langkat | 1833 | 1161 | 1297 | 1833 | 1083 | 1028 | 100% | 93% | 79% | 0·93 (0·87-1·01) | 0·79 (0·73-0·86) |
| Nias Selatan | 174 | 76 | 121 | 174 | 67 | 115 | 100% | 88% | 95% | 0·88 (0·67-1·17) | 0·95 (0·75-1·2) |
| Humbang Hasundutan | 257 | 203 | 284 | 257 | 192 | 220 | 100% | 95% | 77% | 0·95 (0·78-1·14) | 0·77 (0·65-0·93) |
| Pakpak Bharat | 125 | 95 | 90 | 125 | 91 | 79 | 100% | 96% | 88% | 0·96 (0·73-1·25) | 0·88 (0·66-1·16) |
| Samosir | 247 | 172 | 180 | 247 | 138 | 179 | 100% | 80% | 99% | 0·8 (0·65-0·99) | 0·99 (0·82-1·21) |
| Serdang Bedagai | 946 | 618 | 743 | 946 | 591 | 667 | 100% | 96% | 90% | 0·96 (0·86-1·06) | 0·9 (0·81-0·99) |
| Batu Bara | 461 | 469 | 527 | 461 | 460 | 464 | 100% | 98% | 88% | 0·98 (0·86-1·12) | 0·88 (0·77-1) |
| Padang Lawas Utara | 459 | 299 | 428 | 459 | 292 | 363 | 100% | 98% | 85% | 0·98 (0·84-1·13) | 0·85 (0·74-0·97) |
| Padang Lawas | 377 | 347 | 568 | 377 | 286 | 432 | 100% | 82% | 76% | 0·82 (0·71-0·96) | 0·76 (0·66-0·87) |
| Labuhan Batu Selatan | 455 | 368 | 365 | 455 | 334 | 333 | 100% | 91% | 91% | 0·91 (0·79-1·05) | 0·91 (0·79-1·05) |
| Labuhan Batu Utara | 583 | 518 | 595 | 583 | 483 | 585 | 100% | 93% | 98% | 0·93 (0·83-1·05) | 0·98 (0·88-1·1) |

| District | Number of cases | | | Number of cases on treatments | | | Treatment coverage | | | Treatment coverage ratio for 2020 vs 2019 (95% CI) | Treatment coverage ratio for 2021 vs 2019 (95% CI) |
|---|---|---|---|---|---|---|---|---|---|---|---|
| | 2019 | 2020 | 2021 | 2019 | 2020 | 2021 | 2019 | 2020 | 2021 | | |
| Nias Utara | 155 | 94 | 91 | 155 | 93 | 91 | 100% | 99% | 100% | 0·99 (0·77-1·28) | 1 (0-0) |
| Nias Barat | 94 | 49 | 54 | 94 | 49 | 54 | 100% | 100% | 100% | 1 (0-0) | 1 (0-0) |
| Kota Sibolga | 327 | 186 | 440 | 327 | 186 | 336 | 100% | 100% | 76% | 1 (0-0) | 0·76 (0·66-0·89) |
| Kota Tanjung Balai | 798 | 617 | 770 | 327 | 250 | 417 | 41% | 41% | 54% | 0·99 (0·84-1·17) | 1·32 (1·14-1·53) |
| Kota Pematang Siantar | 801 | 483 | 693 | 798 | 432 | 638 | 100% | 89% | 92% | 0·9 (0·8-1·01) | 0·92 (0·83-1·03) |
| Kota Tebing Tinggi | 236 | 226 | 341 | 236 | 222 | 277 | 100% | 98% | 81% | 0·98 (0·82-1·18) | 0·81 (0·68-0·97) |
| Kota Medan | 8186 | 5455 | 7329 | 8186 | 5109 | 5501 | 100% | 94% | 75% | 0·94 (0·9-0·97) | 0·75 (0·73-0·78) |
| Kota Binjai | 477 | 486 | 691 | 477 | 427 | 589 | 100% | 88% | 85% | 0·88 (0·77-1) | 0·85 (0·76-0·96) |
| Kota Padangsidimpuan | 572 | 347 | 454 | 572 | 347 | 374 | 100% | 100% | 82% | 1 (0-0) | 0·82 (0·72-0·94) |
| Kota Gunungsitoli | 217 | 129 | 84 | 217 | 129 | 84 | 100% | 100% | 100% | 1 (0-0) | 1 (0-0) |
| **West Sumatera** | | | | | | | | | | | |
| Kepulauan Mentawai | NA | NA | NA | NA | NA | NA | NA | NA | NA | NA | NA |
| Pesisir Selatan | 1204 | 634 | 705 | 1204 | 634 | 705 | 100% | 100% | 100% | 1 (0-0) | 1 (0-0) |
| Solok | 415 | 95 | 244 | 415 | 95 | 178 | 100% | 100% | 73% | 1 (0-0) | 0·73 (0·61-0·87) |
| Sijunjung | 331 | 114 | 218 | 331 | 100 | 170 | 100% | 88% | 78% | 0·88 (0·7-1·1) | 0·78 (0·65-0·94) |
| Tanah Datar | 461 | 188 | 300 | 461 | 169 | 300 | 100% | 90% | 100% | 0·9 (0·75-1·07) | 1 (0-0) |
| Padang Pariaman | 884 | 1311 | 1815 | 884 | 591 | 790 | 100% | 45% | 44% | 0·45 (0·41-0·5) | 0·44 (0·4-0·48) |
| Agam | 757 | 503 | 503 | 757 | 503 | 494 | 100% | 100% | 98% | 1 (0-0) | 0·98 (0·88-1·1) |
| Lima Puluh Kota | 513 | 308 | 333 | 513 | 308 | 332 | 100% | 100% | 100% | 1 (0-0) | 1 (0·87-1·14) |
| Pasaman | 550 | 338 | 527 | 550 | 338 | 441 | 100% | 100% | 84% | 1 (0-0) | 0·84 (0·74-0·95) |
| Solok Selatan | 279 | 196 | 257 | 279 | 190 | 233 | 100% | 97% | 91% | 0·97 (0·81-1·17) | 0·91 (0·76-1·08) |
| Dharmasraya | 411 | 135 | 214 | 411 | 112 | 149 | 100% | 83% | 70% | 0·83 (0·67-1·02) | 0·7 (0·58-0·84) |
| Pasaman Barat | 815 | 122 | 653 | 815 | 122 | 653 | 100% | 100% | 100% | 1 (0-0) | 1 (0-0) |
| Kota Padang | 3052 | 1644 | 2782 | 3052 | 1644 | 2536 | 100% | 100% | 91% | 1 (0-0) | 0·91 (0·86-0·96) |
| Kota Solok | 269 | 188 | 237 | 269 | 178 | 185 | 100% | 95% | 78% | 0·95 (0·78-1·14) | 0·78 (0·65-0·94) |

| District | Number of cases | | | Number of cases on treatments | | | Treatment coverage | | | Treatment coverage ratio for 2020 vs 2019 (95% CI) | Treatment coverage ratio for 2021 vs 2019 (95% CI) |
|---|---|---|---|---|---|---|---|---|---|---|---|
| | 2019 | 2020 | 2021 | 2019 | 2020 | 2021 | 2019 | 2020 | 2021 | | |
| Kota Sawah Lunto | NA | NA | NA | NA | NA | NA | NA | NA | NA | NA | NA |
| Kota Padang Panjang | 176 | 98 | 190 | 176 | 98 | 158 | 100% | 100% | 83% | 1 (0-0) | 0·83 (0·67-1·03) |
| Kota Bukittinggi | 487 | 316 | 432 | 487 | 283 | 313 | 100% | 90% | 72% | 0·9 (0·77-1·04) | 0·72 (0·63-0·83) |
| Kota Payakumbuh | 266 | 183 | 227 | 266 | 179 | 152 | 100% | 98% | 67% | 0·98 (0·81-1·18) | 0·67 (0·55-0·82) |
| Kota Pariaman | 289 | 93 | 238 | 289 | 67 | 202 | 100% | 72% | 85% | 0·72 (0·55-0·94) | 0·85 (0·71-1·02) |
| **Riau** | | | | | | | | | | | |
| Kuantan Singingi | 348 | 272 | 303 | 348 | 257 | 295 | 100% | 94% | 97% | 0·94 (0·8-1·11) | 0·97 (0·83-1·14) |
| Indragiri Hulu | 361 | 423 | 488 | 361 | 393 | 466 | 100% | 93% | 95% | 0·93 (0·81-1·07) | 0·95 (0·83-1·1) |
| Indragiri Hilir | 475 | 591 | 629 | 475 | 577 | 616 | 100% | 98% | 98% | 0·98 (0·86-1·1) | 0·98 (0·87-1·1) |
| Pelalawan | 656 | 576 | 486 | 656 | 556 | 481 | 100% | 97% | 99% | 0·97 (0·86-1·08) | 0·99 (0·88-1·11) |
| Siak | 480 | 370 | 349 | 480 | 370 | 349 | 100% | 100% | 100% | 1 (0-0) | 1 (0-0) |
| Kampar | 985 | 841 | 856 | 985 | 841 | 856 | 100% | 100% | 100% | 1 (0-0) | 1 (0-0) |
| Rokan Hulu | 1025 | 848 | 795 | 1025 | 830 | 790 | 100% | 98% | 99% | 0·98 (0·89-1·07) | 0·99 (0·91-1·09) |
| Bengkalis | NA | NA | NA | NA | NA | NA | NA | NA | NA | NA | NA |
| Rokan Hilir | 1302 | 1018 | 1064 | 1302 | 973 | 1026 | 100% | 96% | 96% | 0·96 (0·88-1·04) | 0·96 (0·89-1·05) |
| Kepulauan Meranti | 201 | 171 | 214 | 201 | 171 | 174 | 100% | 100% | 81% | 1 (0-0) | 0·81 (0·66-1) |
| Kota Pekanbaru | 3653 | 2825 | 3378 | 3653 | 2391 | 2885 | 100% | 85% | 85% | 0·85 (0·8-0·89) | 0·85 (0·81-0·9) |
| Kota Dumai | 704 | 627 | 685 | 704 | 573 | 616 | 100% | 91% | 90% | 0·91 (0·82-1·02) | 0·9 (0·81-1) |
| **Jambi** | | | | | | | | | | | |
| Kerinci | 135 | 206 | 151 | 135 | 135 | 135 | 100% | 66% | 89% | 0·66 (0·52-0·83) | 0·89 (0·7-1·13) |
| Merangin | 737 | 381 | 550 | 737 | 372 | 424 | 100% | 98% | 77% | 0·98 (0·86-1·11) | 0·77 (0·68-0·87) |
| Sarolangun | 527 | 461 | 463 | 527 | 346 | 409 | 100% | 75% | 88% | 0·75 (0·66-0·86) | 0·88 (0·78-1·01) |
| Batang Hari | 418 | 257 | 381 | 418 | 257 | 335 | 100% | 100% | 88% | 1 (0-0) | 0·88 (0·76-1·02) |
| Muaro Jambi | 455 | 198 | 375 | 455 | 196 | 363 | 100% | 99% | 97% | 0·99 (0·84-1·17) | 0·97 (0·84-1·11) |
| Tanjung Jabung Timur | 228 | 166 | 141 | 228 | 166 | 138 | 100% | 100% | 98% | 1 (0-0) | 0·98 (0·79-1·21) |

| District | Number of cases | | | Number of cases on treatments | | | Treatment coverage | | | Treatment coverage ratio for 2020 vs 2019 (95% CI) | Treatment coverage ratio for 2021 vs 2019 (95% CI) |
|---|---|---|---|---|---|---|---|---|---|---|---|
| | 2019 | 2020 | 2021 | 2019 | 2020 | 2021 | 2019 | 2020 | 2021 | | |
| Tanjung Jabung Barat | 545 | 290 | 319 | 545 | 290 | 289 | 100% | 100% | 91% | 1 (0-0) | 0·91 (0·79-1·04) |
| Tebo | 377 | 240 | 263 | 377 | 218 | 263 | 100% | 91% | 100% | 0·91 (0·77-1·07) | 1 (0-0) |
| Bungo | 291 | 357 | 455 | 291 | 303 | 359 | 100% | 85% | 79% | 0·85 (0·72-1) | 0·79 (0·68-0·92) |
| Kota Jambi | 1055 | 980 | 1229 | 1055 | 744 | 853 | 100% | 76% | 69% | 0·76 (0·69-0·83) | 0·69 (0·63-0·76) |
| Kota Sungai Penuh | 52 | 49 | 72 | 52 | 48 | 65 | 100% | 98% | 90% | 0·98 (0·66-1·45) | 0·9 (0·63-1·3) |
| **South Sumatera** | | | | | | | | | | | |
| Ogan Komering Ulu | 1150 | 398 | 562 | 1150 | 329 | 489 | 100% | 83% | 87% | 0·83 (0·73-0·93) | 0·87 (0·78-0·97) |
| Ogan Komering Ilir | 1711 | 688 | 1068 | 1711 | 650 | 852 | 100% | 94% | 80% | 0·94 (0·86-1·03) | 0·8 (0·73-0·87) |
| Muara Enim | 1732 | 1041 | 1155 | 1732 | 874 | 990 | 100% | 84% | 86% | 0·84 (0·77-0·91) | 0·86 (0·79-0·93) |
| Lahat | 712 | 387 | 586 | 712 | 346 | 368 | 100% | 89% | 63% | 0·89 (0·79-1·02) | 0·63 (0·55-0·71) |
| Musi Rawas | 775 | 515 | 624 | 775 | 468 | 565 | 100% | 91% | 91% | 0·91 (0·81-1·02) | 0·91 (0·81-1·01) |
| Musi Banyuasin | 1588 | 774 | 912 | 1588 | 772 | 887 | 100% | 100% | 97% | 1 (0·92-1·09) | 0·97 (0·9-1·06) |
| Banyu Asin | 2311 | 866 | 1538 | 2311 | 862 | 1538 | 100% | 100% | 100% | 1 (0·92-1·08) | 1 (0-0) |
| Ogan Komering Ulu Selatan | 365 | 186 | 263 | 365 | 165 | 254 | 100% | 89% | 97% | 0·89 (0·74-1·07) | 0·97 (0·82-1·13) |
| Ogan Komering Ulu Timur | 1287 | 497 | 627 | 1287 | 497 | 573 | 100% | 100% | 91% | 1 (0-0) | 0·91 (0·83-1·01) |
| Ogan Ilir | 1035 | 499 | 649 | 1035 | 499 | 568 | 100% | 100% | 88% | 1 (0-0) | 0·88 (0·79-0·97) |
| Empat Lawang | 398 | 293 | 282 | 398 | 268 | 185 | 100% | 91% | 66% | 0·91 (0·78-1·07) | 0·66 (0·55-0·78) |
| Penukal Abab | 410 | 282 | 252 | 410 | 273 | 232 | 100% | 97% | 92% | 0·97 (0·83-1·13) | 0·92 (0·78-1·08) |
| Musi Rawas Utara | 277 | 274 | 355 | 277 | 274 | 307 | 100% | 100% | 86% | 1 (0-0) | 0·86 (0·74-1·02) |
| Kota Palembang | 4589 | 3154 | 5730 | 4589 | 2866 | 4953 | 100% | 91% | 86% | 0·91 (0·87-0·95) | 0·86 (0·83-0·9) |
| Kota Prabumulih | 1057 | 412 | 584 | 1057 | 347 | 425 | 100% | 84% | 73% | 0·84 (0·75-0·95) | 0·73 (0·65-0·81) |
| Kota Pagar Alam | 256 | 140 | 157 | 256 | 105 | 127 | 100% | 75% | 81% | 0·75 (0·6-0·94) | 0·81 (0·65-1) |
| Kota Lubuklinggau | 975 | 292 | 643 | 975 | 277 | 443 | 100% | 95% | 69% | 0·95 (0·83-1·08) | 0·69 (0·62-0·77) |
| **Bengkulu** | | | | | | | | | | | |
| Bengkulu Selatan | 207 | 163 | 151 | 207 | 162 | 147 | 100% | 99% | 97% | 0·99 (0·81-1·22) | 0·97 (0·79-1·2) |

| District | Number of cases | | | Number of cases on treatments | | | Treatment coverage | | | Treatment coverage ratio for 2020 vs 2019 (95% CI) | Treatment coverage ratio for 2021 vs 2019 (95% CI) |
|---|---|---|---|---|---|---|---|---|---|---|---|
| | 2019 | 2020 | 2021 | 2019 | 2020 | 2021 | 2019 | 2020 | 2021 | | |
| Rejang Lebong | 411 | 213 | 191 | 411 | 206 | 178 | 100% | 97% | 93% | 0·97 (0·82-1·14) | 0·93 (0·78-1·11) |
| Bengkulu Utara | 271 | 128 | 179 | 271 | 128 | 179 | 100% | 100% | 100% | 1 (0-0) | 1 (0·9-1·3) |
| Kaur | 121 | 63 | 36 | 121 | 63 | 29 | 100% | 100% | 81% | 1 (0-0) | 0·81 (0·54-1·21) |
| Seluma | 114 | 47 | 85 | 114 | 22 | 71 | 100% | 47% | 84% | 0·47 (0·3-0·73) | 0·84 (0·62-1·12) |
| Mukomuko | 542 | 154 | 281 | 542 | 154 | 216 | 100% | 100% | 77% | 1 (0-0) | 0·77 (0·66-0·9) |
| Lebong | 176 | 158 | 156 | 176 | 153 | 145 | 100% | 97% | 93% | 0·97 (0·78-1·2) | 0·93 (0·75-1·16) |
| Kepahiang | 310 | 269 | 436 | 310 | 242 | 353 | 100% | 90% | 81% | 0·9 (0·76-1·06) | 0·81 (0·7-0·94) |
| Bengkulu Tengah | 134 | 71 | 88 | 134 | 71 | 88 | 100% | 100% | 100% | 1 (0-0) | 1 (0-0) |
| Kota Bengkulu | 986 | 559 | 757 | 986 | 347 | 366 | 100% | 62% | 48% | 0·62 (0·55-0·7) | 0·48 (0·43-0·54) |
| **Lampung** | | | | | | | | | | | |
| Lampung Barat | 278 | 204 | 273 | 278 | 204 | 239 | 100% | 100% | 88% | 1 (0-0) | 0·88 (0·74-1·04) |
| Tanggamus | 847 | 788 | 627 | 847 | 788 | 608 | 100% | 100% | 97% | 1 (0-0) | 0·97 (0·87-1·08) |
| Lampung Selatan | 2268 | 1203 | 1454 | 2268 | 1203 | 1389 | 100% | 100% | 96% | 1 (0-0) | 0·96 (0·89-1·02) |
| Lampung Timur | 1367 | 937 | 1024 | 1367 | 937 | 1024 | 100% | 100% | 100% | 1 (0-0) | 1 (0-0) |
| Lampung Tengah | 2761 | 1999 | 1992 | 982 | 972 | 939 | 36% | 49% | 47% | 1·37 (1·25-1·49) | 1·33 (1·21-1·45) |
| Lampung Utara | 982 | 626 | 1088 | 982 | 626 | 1088 | 100% | 100% | 100% | 1 (0-0) | 1 (0-0) |
| Way Kanan | 728 | 578 | 568 | 728 | 571 | 525 | 100% | 99% | 92% | 0·99 (0·89-1·1) | 0·92 (0·83-1·03) |
| Tulangbawang | 735 | 590 | 620 | 735 | 590 | 591 | 100% | 100% | 95% | 1 (0-0) | 0·95 (0·86-1·06) |
| Pesawaran | 536 | 416 | 467 | 536 | 416 | 467 | 100% | 100% | 100% | 1 (0-0) | 1 (0-0) |
| Pringsewu | 650 | 691 | 767 | 650 | 617 | 533 | 100% | 89% | 69% | 0·89 (0·8-1) | 0·69 (0·62-0·78) |
| Mesuji | 302 | 266 | 237 | 302 | 266 | 200 | 100% | 100% | 84% | 1 (0-0) | 0·84 (0·71-1·01) |
| Tulang Bawang Barat | 416 | 398 | 391 | 416 | 398 | 390 | 100% | 100% | 100% | 1 (0-0) | 1 (0·87-1·15) |
| Pesisir Barat | 161 | 126 | 138 | 161 | 126 | 138 | 100% | 100% | 100% | 1 (0-0) | 1 (0-0) |
| Kota Bandar Lampung | 3571 | 2649 | 3066 | 3571 | 2269 | 2515 | 100% | 86% | 82% | 0·86 (0·81-0·9) | 0·82 (0·78-0·86) |
| Kota Metro | 520 | 499 | 528 | 520 | 360 | 357 | 100% | 72% | 68% | 0·72 (0·63-0·82) | 0·68 (0·59-0·77) |

| District | Number of cases | | | Number of cases on treatments | | | Treatment coverage | | | Treatment coverage ratio for 2020 vs 2019 (95% CI) | Treatment coverage ratio for 2021 vs 2019 (95% CI) |
|---|---|---|---|---|---|---|---|---|---|---|---|
| | 2019 | 2020 | 2021 | 2019 | 2020 | 2021 | 2019 | 2020 | 2021 | | |
| **Bangka Belitung** | | | | | | | | | | | |
| Bangka | 515 | 401 | 420 | 515 | 398 | 387 | 100% | 99% | 92% | 0·99 (0·87-1·13) | 0·92 (0·81-1·05) |
| Belitung | 281 | 238 | 239 | 281 | 227 | 215 | 100% | 95% | 90% | 0·95 (0·8-1·14) | 0·9 (0·75-1·07) |
| Bangka Barat | 203 | 154 | 174 | 203 | 154 | 136 | 100% | 100% | 78% | 1 (0-0) | 0·78 (0·63-0·97) |
| Bangka Tengah | 254 | 155 | 190 | 254 | 140 | 150 | 100% | 90% | 79% | 0·9 (0·73-1·11) | 0·79 (0·65-0·97) |
| Bangka Selatan | 234 | 127 | 153 | 234 | 127 | 151 | 100% | 100% | 99% | 1 (0-0) | 0·99 (0·8-1·21) |
| Belitung Timur | 160 | 149 | 128 | 160 | 149 | 124 | 100% | 100% | 97% | 1 (0-0) | 0·97 (0·77-1·22) |
| Kota Pangkal Pinang | 484 | 529 | 433 | 484 | 418 | 378 | 100% | 79% | 87% | 0·79 (0·69-0·9) | 0·87 (0·76-1) |
| **Riau Island** | | | | | | | | | | | |
| Karimun | 504 | 342 | 392 | 504 | 333 | 310 | 100% | 97% | 79% | 0·97 (0·85-1·12) | 0·79 (0·69-0·91) |
| Bintan | 273 | 211 | 248 | 273 | 210 | 221 | 100% | 100% | 89% | 1 (0·83-1·19) | 0·89 (0·75-1·06) |
| Natuna | 109 | 65 | 60 | 109 | 65 | 53 | 100% | 100% | 88% | 1 (0-0) | 0·88 (0·64-1·23) |
| Lingga | 124 | 109 | 112 | 124 | 108 | 104 | 100% | 99% | 93% | 0·99 (0·77-1·28) | 0·93 (0·72-1·2) |
| Kepulauan Anambas | 89 | 59 | 37 | 89 | 54 | 34 | 100% | 92% | 92% | 0·92 (0·65-1·28) | 0·92 (0·62-1·36) |
| Kota Batam | 3645 | 2701 | 3034 | 3645 | 2621 | 2851 | 100% | 97% | 94% | 0·97 (0·92-1·02) | 0·94 (0·89-0·99) |
| Kota Tanjung Pinang | 744 | 478 | 581 | 744 | 426 | 499 | 100% | 89% | 86% | 0·89 (0·79-1) | 0·86 (0·77-0·96) |
| **DKI Jakarta** | | | | | | | | | | | |
| Kepulauan Seribu | 73 | 48 | 51 | 73 | 42 | 38 | 100% | 88% | 75% | 0·88 (0·6-1·28) | 0·75 (0·5-1·1) |
| Kota Jakarta Selatan | 8116 | 5440 | 6226 | 8116 | 5056 | 5216 | 100% | 93% | 84% | 0·93 (0·9-0·96) | 0·84 (0·81-0·87) |
| Kota Jakarta Timur | 12764 | 8173 | 10763 | 12764 | 7524 | 8796 | 100% | 92% | 82% | 0·92 (0·89-0·95) | 0·82 (0·8-0·84) |
| Kota Jakarta Pusat | 7406 | 5205 | 6387 | 7406 | 4836 | 5183 | 100% | 93% | 81% | 0·93 (0·9-0·96) | 0·81 (0·78-0·84) |
| Kota Jakarta Barat | 8374 | 4903 | 6758 | 8374 | 4634 | 5273 | 100% | 95% | 78% | 0·95 (0·91-0·98) | 0·78 (0·75-0·81) |
| Kota Jakarta Utara | 5333 | 3620 | 5097 | 5333 | 3051 | 4132 | 100% | 84% | 81% | 0·84 (0·81-0·88) | 0·81 (0·78-0·84) |
| **West Java** | | | | | | | | | | | |
| Bogor | 16769 | 11434 | 13466 | 16769 | 10714 | 11982 | 100% | 94% | 89% | 0·94 (0·91-0·96) | 0·89 (0·87-0·91) |

| District | Number of cases | | | Number of cases on treatments | | | Treatment coverage | | | Treatment coverage ratio for 2020 vs 2019 (95% CI) | Treatment coverage ratio for 2021 vs 2019 (95% CI) |
|---|---|---|---|---|---|---|---|---|---|---|---|
| | 2019 | 2020 | 2021 | 2019 | 2020 | 2021 | 2019 | 2020 | 2021 | | |
| Cianjur | 6459 | 4014 | 5364 | 5358 | 3799 | 4637 | 83% | 95% | 86% | 1·14 (1·09-1·19) | 1·04 (1-1·08) |
| Sukabumi | 5358 | 5015 | 5471 | 5358 | 3866 | 4765 | 100% | 77% | 87% | 0·77 (0·74-0·8) | 0·87 (0·84-0·91) |
| Bandung | 7967 | 6284 | 6507 | 7967 | 6115 | 5707 | 100% | 97% | 88% | 0·97 (0·94-1·01) | 0·88 (0·85-0·91) |
| Garut | 5095 | 4529 | 5551 | 5095 | 3720 | 4751 | 100% | 82% | 86% | 0·82 (0·79-0·86) | 0·86 (0·82-0·89) |
| Tasikmalaya | 3016 | 1989 | 2451 | 3016 | 1794 | 2019 | 100% | 90% | 82% | 0·9 (0·85-0·96) | 0·82 (0·78-0·87) |
| Ciamis | 1501 | 1569 | 1771 | 1501 | 1481 | 1587 | 100% | 94% | 90% | 0·94 (0·88-1·01) | 0·9 (0·84-0·96) |
| Kuningan | 2463 | 1964 | 1975 | 2463 | 1831 | 1650 | 100% | 93% | 84% | 0·93 (0·88-0·99) | 0·84 (0·79-0·89) |
| Cirebon | 7879 | 3498 | 3782 | 7879 | 3397 | 3386 | 100% | 97% | 90% | 0·97 (0·93-1·01) | 0·9 (0·86-0·93) |
| Majalengka | 2317 | 1899 | 1999 | 2317 | 1791 | 1707 | 100% | 94% | 85% | 0·94 (0·89-1) | 0·85 (0·8-0·91) |
| Sumedang | 2093 | 1780 | 1778 | 2093 | 1297 | 1362 | 100% | 73% | 77% | 0·73 (0·68-0·78) | 0·77 (0·72-0·82) |
| Indramayu | 3210 | 1679 | 2097 | 3210 | 1529 | 1690 | 100% | 91% | 81% | 0·91 (0·86-0·97) | 0·81 (0·76-0·85) |
| Subang | 3411 | 3353 | 3180 | 3411 | 3164 | 2915 | 100% | 94% | 92% | 0·94 (0·9-0·99) | 0·92 (0·87-0·96) |
| Purwakarta | 2119 | 1936 | 3027 | 2119 | 1722 | 2390 | 100% | 89% | 79% | 0·89 (0·83-0·95) | 0·79 (0·74-0·84) |
| Karawang | 6920 | 5171 | 6043 | 6920 | 4047 | 4509 | 100% | 78% | 75% | 0·78 (0·75-0·81) | 0·75 (0·72-0·77) |
| Bekasi | 9728 | 4896 | 5441 | 9728 | 4577 | 4735 | 100% | 93% | 87% | 0·93 (0·9-0·97) | 0·87 (0·84-0·9) |
| Bandung Barat | 2197 | 1547 | 1985 | 2197 | 1501 | 1745 | 100% | 97% | 88% | 0·97 (0·91-1·04) | 0·88 (0·83-0·94) |
| Pangandaran | 311 | 432 | 528 | 311 | 415 | 383 | 100% | 96% | 73% | 0·96 (0·83-1·11) | 0·73 (0·62-0·84) |
| Kota Bogor | 3834 | 2378 | 4897 | 3834 | 2018 | 4531 | 100% | 85% | 93% | 0·85 (0·8-0·9) | 0·93 (0·89-0·97) |
| Kota Sukabumi | 2103 | 1250 | 1656 | 2103 | 1220 | 1467 | 100% | 98% | 89% | 0·98 (0·91-1·05) | 0·89 (0·83-0·95) |
| Kota Bandung | 13038 | 9227 | 10749 | 13038 | 8746 | 8951 | 100% | 95% | 83% | 0·95 (0·92-0·97) | 0·83 (0·81-0·86) |
| Kota Cirebon | 1862 | 1525 | 2358 | 1862 | 1315 | 1930 | 100% | 86% | 82% | 0·86 (0·8-0·93) | 0·82 (0·77-0·87) |
| Kota Bekasi | 11016 | 5876 | 7018 | 11016 | 5173 | 5969 | 100% | 88% | 85% | 0·88 (0·85-0·91) | 0·85 (0·82-0·88) |
| Kota Depok | 6686 | 3427 | 4609 | 6686 | 3352 | 4063 | 100% | 98% | 88% | 0·98 (0·94-1·02) | 0·88 (0·85-0·92) |
| Kota Cimahi | 2557 | 2022 | 2078 | 2557 | 1722 | 1765 | 100% | 85% | 85% | 0·85 (0·8-0·91) | 0·85 (0·8-0·9) |
| Kota Tasikmalaya | 1671 | 1233 | 1729 | 1671 | 1036 | 1512 | 100% | 84% | 87% | 0·84 (0·78-0·91) | 0·87 (0·82-0·94) |

| District | Number of cases | | | Number of cases on treatments | | | Treatment coverage | | | Treatment coverage ratio for 2020 vs 2019 (95% CI) | Treatment coverage ratio for 2021 vs 2019 (95% CI) |
|---|---|---|---|---|---|---|---|---|---|---|---|
| | 2019 | 2020 | 2021 | 2019 | 2020 | 2021 | 2019 | 2020 | 2021 | | |
| Kota Banjar | 834 | 318 | 382 | 834 | 314 | 267 | 100% | 99% | 70% | 0·99 (0·87-1·12) | 0·7 (0·61-0·8) |
| **Central Java** | | | | | | | | | | | |
| Cilacap | 4077 | 2749 | 2851 | 4077 | 2732 | 2765 | 100% | 99% | 97% | 0·99 (0·95-1·04) | 0·97 (0·92-1·02) |
| Banyumas | 4271 | 3608 | 4426 | 4271 | 3277 | 3711 | 100% | 91% | 84% | 0·91 (0·87-0·95) | 0·84 (0·8-0·88) |
| Purbalingga | 1376 | 1016 | 1167 | 1376 | 1016 | 1046 | 100% | 100% | 90% | 1 (0-0) | 0·9 (0·83-0·97) |
| Banjarnegara | 1258 | 911 | 937 | 1258 | 884 | 850 | 100% | 97% | 91% | 0·97 (0·89-1·06) | 0·91 (0·83-0·99) |
| Kebumen | 2491 | 1904 | 2225 | 2491 | 1867 | 2109 | 100% | 98% | 95% | 0·98 (0·92-1·04) | 0·95 (0·89-1) |
| Purworejo | 704 | 577 | 610 | 704 | 516 | 552 | 100% | 89% | 90% | 0·89 (0·8-1) | 0·9 (0·81-1·01) |
| Wonosobo | 1857 | 1102 | 1387 | 1857 | 1089 | 1294 | 100% | 99% | 93% | 0·99 (0·92-1·06) | 0·93 (0·87-1) |
| Magelang | 751 | 509 | 549 | 751 | 509 | 544 | 100% | 100% | 99% | 1 (0-0) | 0·99 (0·89-1·11) |
| Boyolali | 1452 | 427 | 561 | 1452 | 425 | 527 | 100% | 100% | 94% | 1 (0·89-1·11) | 0·94 (0·85-1·04) |
| Klaten | 1282 | 681 | 1094 | 1282 | 681 | 1008 | 100% | 100% | 92% | 1 (0-0) | 0·92 (0·85-1) |
| Sukoharjo | 749 | 639 | 734 | 749 | 611 | 713 | 100% | 96% | 97% | 0·96 (0·86-1·06) | 0·97 (0·88-1·08) |
| Wonogiri | 1077 | 683 | 684 | 1077 | 683 | 625 | 100% | 100% | 91% | 1 (0-0) | 0·91 (0·83-1·01) |
| Karanganyar | 571 | 414 | 352 | 571 | 407 | 351 | 100% | 98% | 100% | 0·98 (0·87-1·12) | 1 (0·87-1·14) |
| Sragen | 960 | 570 | 523 | 960 | 570 | 505 | 100% | 100% | 97% | 1 (0-0) | 0·97 (0·87-1·08) |
| Grobogan | 1094 | 908 | 991 | 1094 | 877 | 922 | 100% | 97% | 93% | 0·97 (0·88-1·06) | 0·93 (0·85-1·02) |
| Blora | 1366 | 975 | 935 | 1366 | 929 | 755 | 100% | 95% | 81% | 0·95 (0·88-1·04) | 0·81 (0·74-0·88) |
| Rembang | 972 | 578 | 655 | 972 | 578 | 621 | 100% | 100% | 95% | 1 (0-0) | 0·95 (0·86-1·05) |
| Pati | 1770 | 1615 | 1685 | 1770 | 1361 | 1402 | 100% | 84% | 83% | 0·84 (0·79-0·9) | 0·83 (0·78-0·89) |
| Kudus | 1610 | 1239 | 1899 | 1610 | 1087 | 1778 | 100% | 88% | 94% | 0·88 (0·81-0·95) | 0·94 (0·88-1) |
| Jepara | 1182 | 942 | 938 | 1182 | 924 | 868 | 100% | 98% | 93% | 0·98 (0·9-1·07) | 0·93 (0·85-1·01) |
| Demak | 1488 | 1432 | 1113 | 1488 | 1284 | 1056 | 100% | 90% | 95% | 0·9 (0·83-0·97) | 0·95 (0·88-1·03) |
| Semarang | 993 | 632 | 663 | 993 | 632 | 582 | 100% | 100% | 88% | 1 (0-0) | 0·88 (0·79-0·97) |
| Temanggung | 650 | 483 | 543 | 650 | 483 | 520 | 100% | 100% | 96% | 1 (0-0) | 0·96 (0·85-1·07) |

| District | Number of cases | | | Number of cases on treatments | | | Treatment coverage | | | Treatment coverage ratio for 2020 vs 2019 (95% CI) | Treatment coverage ratio for 2021 vs 2019 (95% CI) |
|---|---|---|---|---|---|---|---|---|---|---|---|
| | 2019 | 2020 | 2021 | 2019 | 2020 | 2021 | 2019 | 2020 | 2021 | | |
| Kendal | 1507 | 1386 | 1406 | 1507 | 1342 | 1317 | 100% | 97% | 94% | 0·97 (0·9-1·04) | 0·94 (0·87-1·01) |
| Batang | 1204 | 862 | 912 | 1204 | 857 | 861 | 100% | 99% | 94% | 0·99 (0·91-1·09) | 0·94 (0·87-1·03) |
| Pekalongan | 1255 | 1101 | 1426 | 1255 | 1101 | 1312 | 100% | 100% | 92% | 1 (0-0) | 0·92 (0·85-0·99) |
| Pemalang | 1899 | 1445 | 1790 | 1899 | 1359 | 1625 | 100% | 94% | 91% | 0·94 (0·88-1·01) | 0·91 (0·85-0·97) |
| Tegal | 3855 | 3022 | 3075 | 3855 | 2820 | 2824 | 100% | 93% | 92% | 0·93 (0·89-0·98) | 0·92 (0·87-0·96) |
| Brebes | 3028 | 2057 | 2825 | 3028 | 1931 | 2384 | 100% | 94% | 84% | 0·94 (0·89-0·99) | 0·84 (0·8-0·89) |
| Kota Magelang | 1070 | 883 | 951 | 1070 | 623 | 728 | 100% | 71% | 77% | 0·71 (0·64-0·78) | 0·77 (0·7-0·84) |
| Kota Surakarta | 1902 | 1303 | 1523 | 1902 | 1145 | 1260 | 100% | 88% | 83% | 0·88 (0·82-0·95) | 0·83 (0·77-0·89) |
| Kota Salatiga | 855 | 503 | 588 | 855 | 324 | 408 | 100% | 64% | 69% | 0·64 (0·57-0·73) | 0·69 (0·62-0·78) |
| Kota Semarang | 5403 | 2876 | 3878 | 5403 | 2604 | 3295 | 100% | 91% | 85% | 0·91 (0·86-0·95) | 0·85 (0·81-0·89) |
| Kota Pekalongan | 826 | 839 | 886 | 826 | 554 | 561 | 100% | 66% | 63% | 0·66 (0·59-0·73) | 0·63 (0·57-0·7) |
| Kota Tegal | 2463 | 2245 | 2512 | 2463 | 2013 | 2047 | 100% | 90% | 81% | 0·9 (0·85-0·95) | 0·81 (0·77-0·86) |
| **Di Yogyakarta** | | | | | | | | | | | |
| Kulon Progo | 323 | 240 | 248 | 323 | 240 | 219 | 100% | 100% | 88% | 1 (0-0) | 0·88 (0·74-1·05) |
| Bantul | 1072 | 752 | 793 | 1072 | 683 | 694 | 100% | 91% | 88% | 0·91 (0·83-1) | 0·88 (0·8-0·96) |
| Gunung Kidul | 438 | 328 | 286 | 438 | 308 | 248 | 100% | 94% | 87% | 0·94 (0·81-1·09) | 0·87 (0·74-1·01) |
| Sleman | 1208 | 1177 | 1255 | 1208 | 934 | 1013 | 100% | 79% | 81% | 0·79 (0·73-0·86) | 0·81 (0·74-0·88) |
| Kota Yogyakarta | 1109 | 962 | 1048 | 1109 | 813 | 888 | 100% | 85% | 85% | 0·85 (0·77-0·93) | 0·85 (0·78-0·93) |
| **East Java** | | | | | | | | | | | |
| Pacitan | 362 | 292 | 220 | 362 | 292 | 214 | 100% | 100% | 97% | 1 (0-0) | 0·97 (0·82-1·15) |
| Ponorogo | 1158 | 992 | 762 | 1158 | 921 | 691 | 100% | 93% | 91% | 0·93 (0·85-1·01) | 0·91 (0·83-1) |
| Trenggalek | 551 | 405 | 250 | 551 | 404 | 241 | 100% | 100% | 96% | 1 (0·88-1·13) | 0·96 (0·83-1·12) |
| Tulungagung | 1283 | 963 | 808 | 1283 | 835 | 721 | 100% | 87% | 89% | 0·87 (0·79-0·95) | 0·89 (0·81-0·98) |
| Blitar | 856 | 580 | 485 | 856 | 580 | 485 | 100% | 100% | 100% | 1 (0-0) | 1 (0-0) |
| Kediri | 2013 | 1645 | 1374 | 2013 | 1568 | 1273 | 100% | 95% | 93% | 0·95 (0·89-1·02) | 0·93 (0·86-0·99) |

| District | Number of cases | | | Number of cases on treatments | | | Treatment coverage | | | Treatment coverage ratio for 2020 vs 2019 (95% CI) | Treatment coverage ratio for 2021 vs 2019 (95% CI) |
|---|---|---|---|---|---|---|---|---|---|---|---|
| | 2019 | 2020 | 2021 | 2019 | 2020 | 2021 | 2019 | 2020 | 2021 | | |
| Malang | 2794 | 2084 | 1985 | 2794 | 1837 | 1777 | 100% | 88% | 90% | 0·88 (0·83-0·93) | 0·9 (0·84-0·95) |
| Lumajang | 1924 | 1316 | 1467 | 1924 | 1168 | 1217 | 100% | 89% | 83% | 0·89 (0·83-0·95) | 0·83 (0·77-0·89) |
| Jember | 4355 | 3626 | 3562 | 4355 | 3228 | 3072 | 100% | 89% | 86% | 0·89 (0·85-0·93) | 0·86 (0·82-0·9) |
| Banyuwangi | 2776 | 2094 | 2144 | 2776 | 2014 | 1909 | 100% | 96% | 89% | 0·96 (0·91-1·02) | 0·89 (0·84-0·94) |
| Bondowoso | 1442 | 878 | 1035 | 1442 | 878 | 939 | 100% | 100% | 91% | 1 (0-0) | 0·91 (0·84-0·98) |
| Situbondo | 1249 | 1079 | 1109 | 1249 | 987 | 919 | 100% | 91% | 83% | 0·91 (0·84-0·99) | 0·83 (0·76-0·9) |
| Probolinggo | 1785 | 1182 | 1305 | 1785 | 1174 | 1195 | 100% | 99% | 92% | 0·99 (0·92-1·07) | 0·92 (0·85-0·99) |
| Pasuruan | 3516 | 1945 | 2164 | 3516 | 1795 | 1711 | 100% | 92% | 79% | 0·92 (0·87-0·98) | 0·79 (0·75-0·84) |
| Sidoarjo | 4009 | 2599 | 3231 | 4009 | 2562 | 2729 | 100% | 99% | 84% | 0·99 (0·94-1·04) | 0·84 (0·8-0·89) |
| Mojokerto | 1563 | 989 | 1147 | 1563 | 989 | 1041 | 100% | 100% | 91% | 1 (0-0) | 0·91 (0·84-0·98) |
| Jombang | 1730 | 1299 | 1369 | 1730 | 1299 | 1260 | 100% | 100% | 92% | 1 (0-0) | 0·92 (0·86-0·99) |
| Nganjuk | 1094 | 744 | 738 | 1094 | 730 | 685 | 100% | 98% | 93% | 0·98 (0·89-1·08) | 0·93 (0·84-1·02) |
| Madiun | 1234 | 673 | 648 | 1234 | 598 | 564 | 100% | 89% | 87% | 0·89 (0·81-0·98) | 0·87 (0·79-0·96) |
| Magetan | 766 | 506 | 427 | 766 | 506 | 427 | 100% | 100% | 100% | 1 (0-0) | 1 (0·93-1·17) |
| Ngawi | 1045 | 736 | 692 | 1045 | 736 | 664 | 100% | 100% | 96% | 1 (0-0) | 0·96 (0·87-1·06) |
| Bojonegoro | 1844 | 1463 | 1455 | 1844 | 1443 | 1225 | 100% | 99% | 84% | 0·99 (0·92-1·06) | 0·84 (0·78-0·9) |
| Tuban | 2007 | 1411 | 1333 | 2007 | 1359 | 1250 | 100% | 96% | 94% | 0·96 (0·9-1·03) | 0·94 (0·87-1·01) |
| Lamongan | 2244 | 1627 | 1924 | 2244 | 1539 | 1663 | 100% | 95% | 86% | 0·95 (0·89-1·01) | 0·86 (0·81-0·92) |
| Gresik | 2642 | 1620 | 2023 | 2642 | 1501 | 1814 | 100% | 93% | 90% | 0·93 (0·87-0·99) | 0·9 (0·84-0·95) |
| Bangkalan | 1443 | 1142 | 1192 | 1443 | 1009 | 996 | 100% | 88% | 84% | 0·88 (0·82-0·96) | 0·84 (0·77-0·91) |
| Sampang | 1106 | 768 | 969 | 1106 | 768 | 921 | 100% | 100% | 95% | 1 (0-0) | 0·95 (0·87-1·04) |
| Pamekasan | 1118 | 1025 | 1053 | 1118 | 747 | 804 | 100% | 73% | 76% | 0·73 (0·66-0·8) | 0·76 (0·7-0·84) |
| Sumenep | 1864 | 1606 | 1625 | 1864 | 1606 | 1512 | 100% | 100% | 93% | 1 (0-0) | 0·93 (0·87-1) |
| Kota Kediri | 865 | 738 | 768 | 865 | 588 | 652 | 100% | 80% | 85% | 0·8 (0·72-0·88) | 0·85 (0·77-0·94) |
| Kota Blitar | 280 | 291 | 257 | 280 | 233 | 165 | 100% | 80% | 64% | 0·8 (0·67-0·95) | 0·64 (0·53-0·78) |

| District | Number of cases | | | Number of cases on treatments | | | Treatment coverage | | | Treatment coverage ratio for 2020 vs 2019 (95% CI) | Treatment coverage ratio for 2021 vs 2019 (95% CI) |
|---|---|---|---|---|---|---|---|---|---|---|---|
| | 2019 | 2020 | 2021 | 2019 | 2020 | 2021 | 2019 | 2020 | 2021 | | |
| Kota Malang | 2197 | 1400 | 1593 | 2197 | 1386 | 1352 | 100% | 99% | 85% | 0·99 (0·93-1·06) | 0·85 (0·79-0·91) |
| Kota Probolinggo | 790 | 419 | 457 | 790 | 365 | 362 | 100% | 87% | 79% | 0·87 (0·77-0·99) | 0·79 (0·7-0·9) |
| Kota Pasuruan | 580 | 591 | 706 | 580 | 520 | 590 | 100% | 88% | 84% | 0·88 (0·78-0·99) | 0·84 (0·75-0·94) |
| Kota Mojokerto | 453 | 540 | 629 | 453 | 354 | 477 | 100% | 66% | 76% | 0·66 (0·57-0·75) | 0·76 (0·67-0·86) |
| Kota Madiun | 734 | 655 | 599 | 734 | 535 | 448 | 100% | 82% | 75% | 0·82 (0·73-0·91) | 0·75 (0·67-0·84) |
| Kota Surabaya | 8805 | 4456 | 6182 | 8805 | 4198 | 4742 | 100% | 94% | 77% | 0·94 (0·91-0·98) | 0·77 (0·74-0·79) |
| Kota Batu | 239 | 261 | 187 | 239 | 188 | 137 | 100% | 72% | 73% | 0·72 (0·6-0·87) | 0·73 (0·59-0·9) |
| **Banten** | | | | | | | | | | | |
| Pandeglang | 2331 | 2184 | 2224 | 2331 | 2103 | 2035 | 100% | 96% | 92% | 0·96 (0·91-1·02) | 0·92 (0·86-0·97) |
| Lebak | 2977 | 2023 | 2561 | 2977 | 1973 | 2273 | 100% | 98% | 89% | 0·98 (0·92-1·03) | 0·89 (0·84-0·94) |
| Tangerang | 7859 | 5937 | 7546 | 7859 | 5874 | 6400 | 100% | 99% | 85% | 0·99 (0·96-1·02) | 0·85 (0·82-0·88) |
| Serang | 4019 | 2731 | 3371 | 4019 | 2610 | 2940 | 100% | 96% | 87% | 0·96 (0·91-1) | 0·87 (0·83-0·91) |
| Kota Tangerang | 6174 | 4479 | 6017 | 6174 | 3785 | 4603 | 100% | 85% | 76% | 0·85 (0·81-0·88) | 0·76 (0·74-0·79) |
| Kota Cilegon | 1292 | 1200 | 1162 | 1292 | 866 | 846 | 100% | 72% | 73% | 0·72 (0·66-0·79) | 0·73 (0·67-0·79) |
| Kota Serang | 2299 | 1315 | 1540 | 2299 | 1315 | 1422 | 100% | 100% | 92% | 1 (0-0) | 0·92 (0·86-0·99) |
| Kota Tangerang Selatan | 4064 | 2782 | 3491 | 4064 | 2593 | 3054 | 100% | 93% | 87% | 0·93 (0·89-0·98) | 0·87 (0·83-0·92) |
| **Bali** | | | | | | | | | | | |
| Jembrana | 194 | 128 | 148 | 194 | 128 | 148 | 100% | 100% | 100% | 1 (0-0) | 1 (0-0) |
| Tabanan | 277 | 182 | 190 | 277 | 182 | 190 | 100% | 100% | 100% | 1 (0-0) | 1 (0-0) |
| Badung | 587 | 379 | 385 | 587 | 379 | 385 | 100% | 100% | 100% | 1 (0-0) | 1 (0-0) |
| Gianyar | 347 | 224 | 250 | 347 | 205 | 219 | 100% | 92% | 88% | 0·92 (0·77-1·09) | 0·88 (0·74-1·04) |
| Klungkung | 105 | 131 | 127 | 105 | 115 | 112 | 100% | 88% | 88% | 0·88 (0·67-1·14) | 0·88 (0·68-1·15) |
| Bangli | 88 | 27 | 53 | 88 | 27 | 53 | 100% | 100% | 100% | 1 (0-0) | 1 (0-0) |
| Karangasem | 322 | 193 | 223 | 322 | 193 | 223 | 100% | 100% | 100% | 1 (0-0) | 1 (0-0) |
| Buleleng | 663 | 528 | 691 | 663 | 517 | 628 | 100% | 98% | 91% | 0·98 (0·87-1·1) | 0·91 (0·81-1·01) |

| District | Number of cases | | | Number of cases on treatments | | | Treatment coverage | | | Treatment coverage ratio for 2020 vs 2019 (95% CI) | Treatment coverage ratio for 2021 vs 2019 (95% CI) |
|---|---|---|---|---|---|---|---|---|---|---|---|
| | 2019 | 2020 | 2021 | 2019 | 2020 | 2021 | 2019 | 2020 | 2021 | | |
| Kota Denpasar | 1622 | 1215 | 1306 | 1622 | 1069 | 1065 | 100% | 88% | 82% | 0·88 (0·81-0·95) | 0·82 (0·75-0·88) |
| **West Nusa Tenggara** | | | | | | | | | | | |
| Lombok Barat | 1090 | 742 | 926 | 1090 | 742 | 860 | 100% | 100% | 93% | 1 (0-0) | 0·93 (0·85-1·02) |
| Lombok Tengah | 1096 | 909 | 976 | 1096 | 909 | 976 | 100% | 100% | 100% | 1 (0-0) | 1 (0-0) |
| Lombok Timur | 1501 | 1273 | 1566 | 1501 | 1145 | 1401 | 100% | 90% | 89% | 0·9 (0·83-0·97) | 0·89 (0·83-0·96) |
| Sumbawa | 704 | 447 | 504 | 704 | 435 | 491 | 100% | 97% | 97% | 0·97 (0·86-1·1) | 0·97 (0·87-1·09) |
| Dompu | 379 | 297 | 365 | 379 | 291 | 336 | 100% | 98% | 92% | 0·98 (0·84-1·14) | 0·92 (0·79-1·07) |
| Bima | 717 | 525 | 636 | 717 | 515 | 545 | 100% | 98% | 86% | 0·98 (0·88-1·1) | 0·86 (0·77-0·96) |
| Sumbawa Barat | 276 | 203 | 204 | 276 | 199 | 193 | 100% | 98% | 95% | 0·98 (0·82-1·18) | 0·95 (0·79-1·14) |
| Lombok Utara | 320 | 295 | 251 | 320 | 295 | 228 | 100% | 100% | 91% | 1 (0-0) | 0·91 (0·77-1·08) |
| Kota Mataram | 927 | 850 | 941 | 927 | 639 | 735 | 100% | 75% | 78% | 0·75 (0·68-0·83) | 0·78 (0·71-0·86) |
| Kota Bima | 262 | 210 | 319 | 262 | 210 | 275 | 100% | 100% | 86% | 1 (0-0) | 0·86 (0·73-1·02) |
| **East Nusa Tenggara** | | | | | | | | | | | |
| Sumba Barat | 406 | 270 | 276 | 406 | 192 | 169 | 100% | 71% | 61% | 0·71 (0·6-0·84) | 0·61 (0·51-0·73) |
| Sumba Timur | 394 | 245 | 257 | 394 | 200 | 225 | 100% | 82% | 88% | 0·82 (0·69-0·97) | 0·88 (0·74-1·03) |
| Kupang | 491 | 332 | 317 | 491 | 328 | 298 | 100% | 99% | 94% | 0·99 (0·86-1·14) | 0·94 (0·81-1·09) |
| Timor Tengah Selatan | 528 | 417 | 416 | 528 | 416 | 379 | 100% | 100% | 91% | 1 (0·88-1·13) | 0·91 (0·8-1·04) |
| Timor Tengah Utara | 235 | 247 | 191 | 235 | 210 | 154 | 100% | 85% | 81% | 0·85 (0·71-1·02) | 0·81 (0·66-0·99) |
| Belu | 693 | 453 | 381 | 693 | 453 | 330 | 100% | 100% | 87% | 1 (0-0) | 0·87 (0·76-0·99) |
| Alor | 246 | 274 | 289 | 246 | 261 | 262 | 100% | 95% | 91% | 0·95 (0·8-1·13) | 0·91 (0·76-1·08) |
| Lembata | 201 | 94 | 113 | 201 | 92 | 92 | 100% | 98% | 81% | 0·98 (0·76-1·25) | 0·81 (0·64-1·04) |
| Flores Timur | 227 | 229 | 188 | 227 | 187 | 177 | 100% | 82% | 94% | 0·82 (0·67-0·99) | 0·94 (0·77-1·15) |
| Sikka | 460 | 394 | 499 | 460 | 394 | 422 | 100% | 100% | 85% | 1 (0-0) | 0·85 (0·74-0·96) |
| Ende | 663 | 237 | 346 | 663 | 226 | 321 | 100% | 95% | 93% | 0·95 (0·82-1·11) | 0·93 (0·81-1·06) |
| Ngada | 171 | 116 | 143 | 171 | 113 | 110 | 100% | 97% | 77% | 0·97 (0·77-1·24) | 0·77 (0·61-0·98) |

| District | Number of cases | | | Number of cases on treatments | | | Treatment coverage | | | Treatment coverage ratio for 2020 vs 2019 (95% CI) | Treatment coverage ratio for 2021 vs 2019 (95% CI) |
|---|---|---|---|---|---|---|---|---|---|---|---|
| | 2019 | 2020 | 2021 | 2019 | 2020 | 2021 | 2019 | 2020 | 2021 | | |
| Manggarai | 359 | 242 | 230 | 359 | 203 | 211 | 100% | 84% | 92% | 0·84 (0·71-1) | 0·92 (0·77-1·09) |
| Rote Ndao | 120 | 100 | 58 | 120 | 77 | 52 | 100% | 77% | 90% | 0·77 (0·58-1·02) | 0·9 (0·65-1·24) |
| Manggarai Barat | 326 | 263 | 273 | 326 | 248 | 230 | 100% | 94% | 84% | 0·94 (0·8-1·11) | 0·84 (0·71-1) |
| Sumba Tengah | 114 | 77 | 92 | 114 | 72 | 92 | 100% | 94% | 100% | 0·94 (0·7-1·26) | 1 (0-0) |
| Sumba Barat Daya | 472 | 550 | 832 | 472 | 512 | 495 | 100% | 93% | 59% | 0·93 (0·82-1·05) | 0·59 (0·53-0·67) |
| Nagekeo | 162 | 151 | 126 | 162 | 146 | 116 | 100% | 97% | 92% | 0·97 (0·77-1·21) | 0·92 (0·73-1·17) |
| Manggarai Timur | 205 | 143 | 191 | 205 | 137 | 189 | 100% | 96% | 99% | 0·96 (0·77-1·19) | 0·99 (0·81-1·21) |
| Sabu Raijua | 85 | 86 | 51 | 85 | 86 | 51 | 100% | 100% | 100% | 1 (0-0) | 1 (0-0) |
| Malaka | 370 | 379 | 376 | 370 | 210 | 200 | 100% | 55% | 53% | 0·55 (0·47-0·65) | 0·53 (0·45-0·63) |
| Kota Kupang | 667 | 567 | 613 | 667 | 518 | 491 | 100% | 91% | 80% | 0·91 (0·81-1·02) | 0·8 (0·71-0·9) |
| **West Kalimantan** | | | | | | | | | | | |
| Sambas | 1039 | 858 | 1171 | 1039 | 771 | 1017 | 100% | 90% | 87% | 0·9 (0·82-0·99) | 0·87 (0·8-0·95) |
| Bengkayang | 331 | 416 | 451 | 331 | 351 | 359 | 100% | 84% | 80% | 0·84 (0·73-0·98) | 0·8 (0·69-0·92) |
| Landak | 561 | 383 | 391 | 561 | 383 | 351 | 100% | 100% | 90% | 1 (0-0) | 0·9 (0·79-1·03) |
| Mempawah | 360 | 285 | 274 | 360 | 285 | 238 | 100% | 100% | 87% | 1 (0-0) | 0·87 (0·74-1·02) |
| Sanggau | 836 | 700 | 651 | 836 | 623 | 554 | 100% | 89% | 85% | 0·89 (0·8-0·99) | 0·85 (0·76-0·95) |
| Ketapang | 660 | 611 | 666 | 660 | 533 | 535 | 100% | 87% | 80% | 0·87 (0·78-0·98) | 0·8 (0·72-0·9) |
| Sintang | 1086 | 396 | 508 | 1086 | 353 | 470 | 100% | 89% | 93% | 0·89 (0·79-1·01) | 0·93 (0·83-1·03) |
| Kapuas Hulu | 459 | 69 | 377 | 459 | 69 | 239 | 100% | 100% | 63% | 1 (0-0) | 0·63 (0·54-0·74) |
| Sekadau | 313 | 185 | 255 | 313 | 154 | 218 | 100% | 83% | 85% | 0·83 (0·69-1·01) | 0·85 (0·72-1·02) |
| Melawi | 468 | 335 | 453 | 468 | 305 | 403 | 100% | 91% | 89% | 0·91 (0·79-1·05) | 0·89 (0·78-1·02) |
| Kayong Utara | 194 | 81 | 129 | 194 | 52 | 116 | 100% | 64% | 90% | 0·64 (0·47-0·87) | 0·9 (0·71-1·13) |
| Kubu Raya | 610 | 496 | 580 | 610 | 496 | 543 | 100% | 100% | 94% | 1 (0-0) | 0·94 (0·83-1·05) |
| Kota Pontianak | 1728 | 1466 | 1908 | 1728 | 1241 | 1606 | 100% | 85% | 84% | 0·85 (0·79-0·91) | 0·84 (0·79-0·9) |
| Kota Singkawang | 884 | 859 | 932 | 884 | 732 | 706 | 100% | 85% | 76% | 0·85 (0·77-0·94) | 0·76 (0·69-0·84) |

| District | Number of cases | | | Number of cases on treatments | | | Treatment coverage | | | Treatment coverage ratio for 2020 vs 2019 (95% CI) | Treatment coverage ratio for 2021 vs 2019 (95% CI) |
|---|---|---|---|---|---|---|---|---|---|---|---|
| | 2019 | 2020 | 2021 | 2019 | 2020 | 2021 | 2019 | 2020 | 2021 | | |
| **Central Kalimantan** | | | | | | | | | | | |
| Kotawaringin Barat | 526 | 320 | 393 | 526 | 317 | 393 | 100% | 99% | 100% | 0·99 (0·86-1·14) | 1 (0-0) |
| Kotawaringin Timur | 555 | 474 | 508 | 555 | 466 | 467 | 100% | 98% | 92% | 0·98 (0·87-1·11) | 0·92 (0·81-1·04) |
| Kapuas | 375 | 257 | 295 | 375 | 247 | 283 | 100% | 96% | 96% | 0·96 (0·82-1·13) | 0·96 (0·82-1·12) |
| Barito Selatan | 211 | 149 | 187 | 211 | 125 | 183 | 100% | 84% | 98% | 0·84 (0·67-1·05) | 0·98 (0·8-1·19) |
| Barito Utara | 252 | 146 | 159 | 252 | 110 | 102 | 100% | 75% | 64% | 0·75 (0·6-0·94) | 0·64 (0·51-0·81) |
| Sukamara | 98 | 81 | 87 | 98 | 48 | 66 | 100% | 59% | 76% | 0·59 (0·42-0·83) | 0·76 (0·56-1·04) |
| Lamandau | 154 | 91 | 115 | 154 | 91 | 103 | 100% | 100% | 90% | 1 (0-0) | 0·9 (0·7-1·15) |
| Seruyan | 208 | 150 | 353 | 208 | 140 | 163 | 100% | 93% | 46% | 0·93 (0·75-1·16) | 0·46 (0·38-0·56) |
| Katingan | 208 | 131 | 188 | 208 | 119 | 152 | 100% | 91% | 81% | 0·91 (0·73-1·14) | 0·81 (0·66-1) |
| Pulang Pisau | 116 | 59 | 75 | 116 | 59 | 55 | 100% | 100% | 73% | 1 (0-0) | 0·73 (0·53-1·01) |
| Gunung Mas | 178 | 154 | 132 | 178 | 132 | 117 | 100% | 86% | 89% | 0·86 (0·68-1·07) | 0·89 (0·7-1·12) |
| Barito Timur | 100 | 81 | 131 | 100 | 80 | 112 | 100% | 99% | 85% | 0·99 (0·74-1·33) | 0·85 (0·65-1·12) |
| Murung Raya | 244 | 205 | 202 | 244 | 195 | 190 | 100% | 95% | 94% | 0·95 (0·79-1·15) | 0·94 (0·78-1·14) |
| Kota Palangka Raya | 623 | 279 | 619 | 623 | 275 | 423 | 100% | 99% | 68% | 0·99 (0·86-1·14) | 0·68 (0·6-0·77) |
| **South Kalimantan** | | | | | | | | | | | |
| Tanah Laut | 448 | 218 | 309 | 448 | 198 | 254 | 100% | 91% | 82% | 0·91 (0·77-1·07) | 0·82 (0·7-0·96) |
| Kotabaru | 365 | 268 | 369 | 365 | 250 | 313 | 100% | 93% | 85% | 0·93 (0·79-1·1) | 0·85 (0·73-0·99) |
| Banjar | 1271 | 608 | 657 | 1271 | 601 | 561 | 100% | 99% | 85% | 0·99 (0·9-1·09) | 0·85 (0·77-0·94) |
| Barito Kuala | 269 | 205 | 235 | 269 | 132 | 196 | 100% | 64% | 83% | 0·64 (0·52-0·79) | 0·83 (0·69-1) |
| Tapin | 282 | 125 | 172 | 282 | 125 | 155 | 100% | 100% | 90% | 1 (0-0) | 0·9 (0·74-1·1) |
| Hulu Sungai Selatan | 421 | 174 | 290 | 421 | 168 | 196 | 100% | 97% | 68% | 0·97 (0·81-1·15) | 0·68 (0·57-0·8) |
| Hulu Sungai Tengah | 412 | 370 | 436 | 412 | 351 | 382 | 100% | 95% | 88% | 0·95 (0·82-1·09) | 0·88 (0·76-1·01) |
| Hulu Sungai Utara | 329 | 206 | 302 | 329 | 200 | 260 | 100% | 97% | 86% | 0·97 (0·81-1·16) | 0·86 (0·73-1·01) |
| Tabalong | 360 | 206 | 265 | 360 | 171 | 233 | 100% | 83% | 88% | 0·83 (0·69-1) | 0·88 (0·75-1·04) |

| District | Number of cases | | | Number of cases on treatments | | | Treatment coverage | | | Treatment coverage ratio for 2020 vs 2019 (95% CI) | Treatment coverage ratio for 2021 vs 2019 (95% CI) |
|---|---|---|---|---|---|---|---|---|---|---|---|
| | 2019 | 2020 | 2021 | 2019 | 2020 | 2021 | 2019 | 2020 | 2021 | | |
| Tanah Bumbu | 341 | 161 | 254 | 341 | 161 | 233 | 100% | 100% | 92% | 1 (0-0) | 0·92 (0·78-1·08) |
| Balangan | 184 | 69 | 149 | 184 | 68 | 116 | 100% | 99% | 78% | 0·99 (0·75-1·3) | 0·78 (0·62-0·98) |
| Kota Banjarmasin | 2155 | 886 | 1239 | 468 | 346 | 266 | 22% | 39% | 21% | 1·8 (1·57-2·06) | 0·99 (0·85-1·15) |
| Kota Banjar Baru | 468 | 269 | 305 | 468 | 269 | 305 | 100% | 100% | 100% | 1 (0-0) | 1 (0-0) |
| **East Kalimantan** | | | | | | | | | | | |
| Paser | 462 | 368 | 346 | 462 | 367 | 346 | 100% | 100% | 100% | 1 (0·87-1·14) | 1 (0-0) |
| Kutai Barat | 359 | 177 | 310 | 359 | 165 | 280 | 100% | 93% | 90% | 0·93 (0·78-1·12) | 0·9 (0·77-1·06) |
| Kutai Kartanegara | 956 | 601 | 773 | 956 | 554 | 702 | 100% | 92% | 91% | 0·92 (0·83-1·02) | 0·91 (0·82-1) |
| Kutai Timur | 631 | 447 | 476 | 631 | 365 | 435 | 100% | 82% | 91% | 0·82 (0·72-0·93) | 0·91 (0·81-1·03) |
| Berau | 498 | 234 | 281 | 498 | 226 | 226 | 100% | 97% | 80% | 0·97 (0·83-1·13) | 0·8 (0·69-0·94) |
| Penajam Paser Utara | 223 | 214 | 193 | 223 | 126 | 155 | 100% | 59% | 80% | 0·59 (0·47-0·73) | 0·8 (0·65-0·99) |
| Mahakam Hulu | 48 | 39 | 39 | 48 | 39 | 39 | 100% | 100% | 100% | 1 (0-0) | 1 (0-0) |
| Kota Balikpapan | 1797 | 952 | 1387 | 1797 | 930 | 1154 | 100% | 98% | 83% | 0·98 (0·9-1·06) | 0·83 (0·77-0·9) |
| Kota Samarinda | 1864 | 1726 | 1979 | 1864 | 1038 | 1434 | 100% | 60% | 72% | 0·6 (0·56-0·65) | 0·72 (0·68-0·78) |
| Kota Bontang | 899 | 581 | 547 | 899 | 525 | 437 | 100% | 90% | 80% | 0·9 (0·81-1·01) | 0·8 (0·71-0·9) |
| **North Kalimantan** | | | | | | | | | | | |
| Malinau | 285 | 169 | 172 | 285 | 169 | 169 | 100% | 100% | 98% | 1 (0-0) | 0·98 (0·81-1·19) |
| Bulungan | 355 | 180 | 198 | 355 | 164 | 171 | 100% | 91% | 86% | 0·91 (0·76-1·1) | 0·86 (0·72-1·04) |
| Nunukan | 424 | 186 | 254 | 424 | 169 | 210 | 100% | 91% | 83% | 0·91 (0·76-1·09) | 0·83 (0·7-0·98) |
| Tana Tidung | 47 | 17 | 24 | 47 | 17 | 24 | 100% | 100% | 100% | 1 (0-0) | 1 (0-0) |
| Kota Tarakan | 677 | 473 | 518 | 677 | 434 | 435 | 100% | 92% | 84% | 0·92 (0·81-1·04) | 0·84 (0·74-0·95) |
| **North Sulawesi** | | | | | | | | | | | |
| Bolaang Mongondow | 521 | 536 | 615 | 521 | 524 | 527 | 100% | 98% | 86% | 0·98 (0·87-1·1) | 0·86 (0·76-0·97) |
| Minahasa | 716 | 451 | 677 | 716 | 446 | 614 | 100% | 99% | 91% | 0·99 (0·88-1·11) | 0·91 (0·81-1·01) |
| Kepulauan Sangihe | 257 | 138 | 231 | 257 | 138 | 231 | 100% | 100% | 100% | 1 (0-0) | 1 (0-0) |

| District | Number of cases | | | Number of cases on treatments | | | Treatment coverage | | | Treatment coverage ratio for 2020 vs 2019 (95% CI) | Treatment coverage ratio for 2021 vs 2019 (95% CI) |
|---|---|---|---|---|---|---|---|---|---|---|---|
| | 2019 | 2020 | 2021 | 2019 | 2020 | 2021 | 2019 | 2020 | 2021 | | |
| Kepulauan Talaud | 202 | 197 | 214 | 202 | 167 | 163 | 100% | 85% | 76% | 0·85 (0·69-1·04) | 0·76 (0·62-0·94) |
| Minahasa Selatan | 505 | 259 | 290 | 202 | 205 | 213 | 40% | 79% | 73% | 1·98 (1·64-2·39) | 1·84 (1·52-2·22) |
| Minahasa Utara | 512 | 450 | 431 | 505 | 296 | 256 | 99% | 66% | 59% | 0·67 (0·58-0·77) | 0·6 (0·52-0·7) |
| Bolaang Mongondow Utara | 196 | 127 | 236 | 196 | 127 | 236 | 100% | 100% | 100% | 1 (0-0) | 1 (0-0) |
| Kep, Siau Tagulandang Biaro | 165 | 119 | 110 | 165 | 115 | 110 | 100% | 97% | 100% | 0·97 (0·76-1·23) | 1 (0-0) |
| Minahasa Tenggara | 193 | 163 | 245 | 193 | 116 | 154 | 100% | 71% | 63% | 0·71 (0·57-0·89) | 0·63 (0·51-0·78) |
| Bolaang Mongondow Selatan | 228 | 118 | 130 | 165 | 113 | 105 | 72% | 96% | 81% | 1·32 (1·04-1·68) | 1·12 (0·87-1·43) |
| Bolaang Mongondow Timur | 213 | 141 | 163 | 193 | 141 | 163 | 91% | 100% | 100% | 1·1 (0·89-1·37) | 1·1 (0·9-1·36) |
| Kota Manado | 2688 | 1471 | 1958 | 228 | 265 | 123 | 8% | 18% | 6% | 2·12 (1·79-2·52) | 0·74 (0·6-0·92) |
| Kota Bitung | 759 | 543 | 604 | 213 | 185 | 140 | 28% | 34% | 23% | 1·21 (1-1·48) | 0·83 (0·67-1·02) |
| Kota Tomohon | 347 | 229 | 366 | 347 | 229 | 366 | 100% | 100% | 100% | 1 (0-0) | 1 (0-0) |
| Kota Kotamobagu | 350 | 256 | 439 | 350 | 256 | 439 | 100% | 100% | 100% | 1 (0-0) | 1 (0-0) |
| **Central Sulawesi** | | | | | | | | | | | |
| Banggai Kepulauan | 263 | 159 | 195 | 263 | 159 | 195 | 100% | 100% | 100% | 1 (0-0) | 1 (0-0) |
| Banggai | 1094 | 835 | 743 | 350 | 340 | 307 | 32% | 41% | 41% | 1·27 (1·1-1·48) | 1·29 (1·11-1·5) |
| Morowali | 451 | 226 | 394 | 263 | 180 | 166 | 58% | 80% | 42% | 1·37 (1·13-1·65) | 0·72 (0·6-0·88) |
| Poso | 277 | 253 | 263 | 277 | 215 | 263 | 100% | 85% | 100% | 0·85 (0·71-1·02) | 1 (0-0) |
| Donggala | 539 | 269 | 306 | 277 | 214 | 218 | 51% | 80% | 71% | 1·55 (1·3-1·85) | 1·39 (1·16-1·65) |
| Toli-Toli | 497 | 299 | 321 | 497 | 283 | 293 | 100% | 95% | 91% | 0·95 (0·82-1·1) | 0·91 (0·79-1·05) |
| Buol | 226 | 153 | 224 | 226 | 153 | 224 | 100% | 100% | 100% | 1 (0-0) | 1 (0-0) |
| Parigi Moutong | 677 | 630 | 505 | 677 | 594 | 443 | 100% | 94% | 88% | 0·94 (0·84-1·05) | 0·88 (0·78-0·99) |
| Tojo Una-Una | 279 | 231 | 169 | 279 | 213 | 130 | 100% | 92% | 77% | 0·92 (0·77-1·1) | 0·77 (0·63-0·95) |
| Sigi | 433 | 353 | 275 | 433 | 353 | 252 | 100% | 100% | 92% | 1 (0-0) | 0·92 (0·78-1·07) |
| Banggai Laut | 119 | 147 | 167 | 119 | 126 | 127 | 100% | 86% | 76% | 0·86 (0·67-1·1) | 0·76 (0·59-0·98) |
| Morowali Utara | 188 | 147 | 169 | 188 | 147 | 143 | 100% | 100% | 85% | 1 (0-0) | 0·85 (0·68-1·05) |

| District | Number of cases | | | Number of cases on treatments | | | Treatment coverage | | | Treatment coverage ratio for 2020 vs 2019 (95% CI) | Treatment coverage ratio for 2021 vs 2019 (95% CI) |
|---|---|---|---|---|---|---|---|---|---|---|---|
| | 2019 | 2020 | 2021 | 2019 | 2020 | 2021 | 2019 | 2020 | 2021 | | |
| Kota Palu | 779 | 656 | 1030 | 779 | 582 | 764 | 100% | 89% | 74% | 0·89 (0·8-0·99) | 0·74 (0·67-0·82) |
| **South Sulawesi** | | | | | | | | | | | |
| Kepulauan Selayar | 220 | 197 | 247 | 220 | 157 | 222 | 100% | 80% | 90% | 0·8 (0·65-0·98) | 0·9 (0·75-1·08) |
| Bulukumba | 652 | 403 | 582 | 652 | 391 | 547 | 100% | 97% | 94% | 0·97 (0·86-1·1) | 0·94 (0·84-1·05) |
| Bantaeng | 347 | 367 | 466 | 347 | 292 | 325 | 100% | 80% | 70% | 0·8 (0·68-0·93) | 0·7 (0·6-0·81) |
| Jeneponto | 602 | 403 | 590 | 602 | 403 | 569 | 100% | 100% | 96% | 1 (0-0) | 0·96 (0·86-1·08) |
| Takalar | 696 | 449 | 728 | 696 | 440 | 633 | 100% | 98% | 87% | 0·98 (0·87-1·1) | 0·87 (0·78-0·97) |
| Gowa | 1805 | 875 | 1223 | 1805 | 875 | 1223 | 100% | 100% | 100% | 1 (0-0) | 1 (0-0) |
| Sinjai | 535 | 340 | 397 | 535 | 307 | 354 | 100% | 90% | 89% | 0·9 (0·78-1·04) | 0·89 (0·78-1·02) |
| Maros | 673 | 433 | 542 | 673 | 431 | 536 | 100% | 100% | 99% | 1 (0·88-1·12) | 0·99 (0·88-1·11) |
| Pangkajene Dan Kepulauan | 811 | 611 | 707 | 811 | 606 | 681 | 100% | 99% | 96% | 0·99 (0·89-1·1) | 0·96 (0·87-1·07) |
| Barru | 265 | 200 | 263 | 265 | 200 | 256 | 100% | 100% | 97% | 1 (0-0) | 0·97 (0·82-1·16) |
| Bone | 1282 | 851 | 1067 | 1282 | 831 | 1023 | 100% | 98% | 96% | 0·98 (0·89-1·07) | 0·96 (0·88-1·04) |
| Soppeng | 379 | 249 | 212 | 379 | 249 | 212 | 100% | 100% | 100% | 1 (0-0) | 1 (0-0) |
| Wajo | 877 | 575 | 773 | 877 | 575 | 708 | 100% | 100% | 92% | 1 (0-0) | 0·92 (0·83-1·01) |
| Sidenreng Rappang | 584 | 375 | 520 | 584 | 372 | 435 | 100% | 99% | 84% | 0·99 (0·87-1·13) | 0·84 (0·74-0·95) |
| Pinrang | 580 | 438 | 688 | 580 | 398 | 644 | 100% | 91% | 94% | 0·91 (0·8-1·03) | 0·94 (0·84-1·05) |
| Enrekang | 207 | 159 | 203 | 207 | 159 | 188 | 100% | 100% | 93% | 1 (0-0) | 0·93 (0·76-1·13) |
| Luwu | 622 | 415 | 496 | 622 | 415 | 496 | 100% | 100% | 100% | 1 (0-0) | 1 (0-0) |
| Tana Toraja | 295 | 162 | 204 | 295 | 155 | 170 | 100% | 96% | 83% | 0·96 (0·79-1·16) | 0·83 (0·69-1·01) |
| Luwu Utara | 609 | 328 | 383 | 609 | 324 | 372 | 100% | 99% | 97% | 0·99 (0·86-1·13) | 0·97 (0·85-1·1) |
| Luwu Timur | 444 | 345 | 449 | 444 | 308 | 301 | 100% | 89% | 67% | 0·89 (0·77-1·03) | 0·67 (0·58-0·78) |
| Toraja Utara | 230 | 222 | 270 | 230 | 202 | 260 | 100% | 91% | 96% | 0·91 (0·75-1·1) | 0·96 (0·81-1·15) |
| Kota Makassar | 6731 | 4849 | 5934 | 6731 | 3464 | 4046 | 100% | 71% | 68% | 0·71 (0·69-0·74) | 0·68 (0·66-0·71) |
| Kota Parepare | 460 | 427 | 399 | 460 | 302 | 292 | 100% | 71% | 73% | 0·71 (0·61-0·82) | 0·73 (0·63-0·85) |

| District | Number of cases | | | Number of cases on treatments | | | Treatment coverage | | | Treatment coverage ratio for 2020 vs 2019 (95% CI) | Treatment coverage ratio for 2021 vs 2019 (95% CI) |
|---|---|---|---|---|---|---|---|---|---|---|---|
| | 2019 | 2020 | 2021 | 2019 | 2020 | 2021 | 2019 | 2020 | 2021 | | |
| Kota Palopo | 454 | 387 | 606 | 454 | 315 | 411 | 100% | 81% | 68% | 0·81 (0·71-0·94) | 0·68 (0·59-0·77) |
| **Southeast Sulawesi** | | | | | | | | | | | |
| Buton | 239 | 155 | 194 | 239 | 141 | 166 | 100% | 91% | 86% | 0·91 (0·74-1·12) | 0·86 (0·7-1·04) |
| Muna | 406 | 257 | 285 | 406 | 257 | 269 | 100% | 100% | 94% | 1 (0-0) | 0·94 (0·81-1·1) |
| Konawe | 362 | 169 | 437 | 362 | 161 | 279 | 100% | 95% | 64% | 0·95 (0·79-1·15) | 0·64 (0·55-0·75) |
| Kolaka | 377 | 320 | 379 | 377 | 315 | 359 | 100% | 98% | 95% | 0·98 (0·85-1·14) | 0·95 (0·82-1·09) |
| Konawe Selatan | 475 | 330 | 357 | 475 | 304 | 320 | 100% | 92% | 90% | 0·92 (0·8-1·06) | 0·9 (0·78-1·03) |
| Bombana | 451 | 574 | 412 | 451 | 208 | 253 | 100% | 36% | 61% | 0·36 (0·31-0·42) | 0·61 (0·53-0·72) |
| Wakatobi | 131 | 46 | 90 | 131 | 38 | 78 | 100% | 83% | 87% | 0·83 (0·58-1·18) | 0·87 (0·65-1·15) |
| Kolaka Utara | 165 | 155 | 176 | 165 | 151 | 159 | 100% | 97% | 90% | 0·97 (0·78-1·21) | 0·9 (0·73-1·12) |
| Buton Utara | 81 | 40 | 39 | 81 | 36 | 26 | 100% | 90% | 67% | 0·9 (0·61-1·33) | 0·67 (0·43-1·03) |
| Konawe Utara | 140 | 100 | 128 | 140 | 97 | 122 | 100% | 97% | 95% | 0·97 (0·75-1·26) | 0·95 (0·75-1·21) |
| Kolaka Timur | 173 | 118 | 114 | 173 | 118 | 111 | 100% | 100% | 97% | 1 (0-0) | 0·97 (0·77-1·24) |
| Konawe Kepulauan | 45 | 71 | 62 | 45 | 69 | 59 | 100% | 97% | 95% | 0·97 (0·67-1·41) | 0·95 (0·65-1·4) |
| Muna Barat | 140 | 88 | 112 | 140 | 82 | 99 | 100% | 93% | 88% | 0·93 (0·71-1·22) | 0·88 (0·68-1·14) |
| Buton Tengah | 194 | 159 | 198 | 194 | 155 | 181 | 100% | 97% | 91% | 0·97 (0·79-1·2) | 0·91 (0·75-1·12) |
| Buton Selatan | 101 | 83 | 74 | 101 | 83 | 74 | 100% | 100% | 100% | 1 (0-0) | 1 (0-0) |
| Kota Kendari | 712 | 510 | 1177 | 712 | 467 | 815 | 100% | 92% | 69% | 0·92 (0·81-1·03) | 0·69 (0·63-0·77) |
| Kota Baubau | 332 | 342 | 523 | 332 | 227 | 328 | 100% | 66% | 63% | 0·66 (0·56-0·78) | 0·63 (0·54-0·73) |
| **Gorontalo** | | | | | | | | | | | |
| Boalemo | 377 | 283 | 363 | 377 | 283 | 296 | 100% | 100% | 82% | 1 (0-0) | 0·82 (0·7-0·95) |
| Gorontalo | 1417 | 631 | 1305 | 1417 | 622 | 962 | 100% | 99% | 74% | 0·99 (0·9-1·08) | 0·74 (0·68-0·8) |
| Pohuwato | 446 | 312 | 309 | 446 | 280 | 223 | 100% | 90% | 72% | 0·9 (0·77-1·04) | 0·72 (0·61-0·85) |
| Bone Bolango | 633 | 495 | 753 | 633 | 471 | 530 | 100% | 95% | 70% | 0·95 (0·84-1·07) | 0·7 (0·63-0·79) |
| Gorontalo Utara | 349 | 262 | 303 | 349 | 254 | 255 | 100% | 97% | 84% | 0·97 (0·82-1·14) | 0·84 (0·72-0·99) |

| District | Number of cases | | | Number of cases on treatments | | | Treatment coverage | | | Treatment coverage ratio for 2020 vs 2019 (95% CI) | Treatment coverage ratio for 2021 vs 2019 (95% CI) |
|---|---|---|---|---|---|---|---|---|---|---|---|
| | 2019 | 2020 | 2021 | 2019 | 2020 | 2021 | 2019 | 2020 | 2021 | | |
| Kota Gorontalo | 798 | 426 | 799 | 798 | 405 | 507 | 100% | 95% | 63% | 0·95 (0·84-1·07) | 0·63 (0·57-0·71) |
| **West Sulawesi** | | | | | | | | | | | |
| Majene | 512 | 439 | 482 | 512 | 417 | 450 | 100% | 95% | 93% | 0·95 (0·83-1·08) | 0·93 (0·82-1·06) |
| Polewali Mandar | 913 | 655 | 868 | 913 | 655 | 781 | 100% | 100% | 90% | 1 (0-0) | 0·9 (0·82-0·99) |
| Mamasa | 169 | 101 | 113 | 169 | 101 | 107 | 100% | 100% | 95% | 1 (0-0) | 0·95 (0·74-1·21) |
| Mamuju | 647 | 459 | 521 | 647 | 459 | 521 | 100% | 100% | 100% | 1 (0-0) | 1 (0-0) |
| Pasangkayu | 237 | 174 | 176 | 237 | 174 | 166 | 100% | 100% | 94% | 1 (0-0) | 0·94 (0·77-1·15) |
| Mamuju Tengah | 258 | 138 | 222 | 258 | 138 | 222 | 100% | 100% | 100% | 1 (0-0) | 1 (0-0) |
| **Maluku** | | | | | | | | | | | |
| Maluku Tenggara Barat | 350 | 85 | 222 | 350 | 78 | 200 | 100% | 92% | 90% | 0·92 (0·72-1·17) | 0·9 (0·76-1·07) |
| Maluku Tenggara | 382 | 249 | 326 | 382 | 242 | 240 | 100% | 97% | 74% | 0·97 (0·83-1·14) | 0·74 (0·63-0·86) |
| Maluku Tengah | 747 | 285 | 644 | 747 | 218 | 492 | 100% | 76% | 76% | 0·76 (0·66-0·89) | 0·76 (0·68-0·86) |
| Buru | 140 | 121 | 154 | 140 | 99 | 136 | 100% | 82% | 88% | 0·82 (0·63-1·06) | 0·88 (0·7-1·12) |
| Kepulauan Aru | 421 | 134 | 220 | 421 | 126 | 168 | 100% | 94% | 76% | 0·94 (0·77-1·15) | 0·76 (0·64-0·91) |
| Seram Bagian Barat | 206 | 191 | 206 | 206 | 191 | 187 | 100% | 100% | 91% | 1 (0-0) | 0·91 (0·74-1·11) |
| Seram Bagian Timur | 156 | 87 | 208 | 156 | 85 | 159 | 100% | 98% | 76% | 0·98 (0·75-1·27) | 0·76 (0·61-0·95) |
| Maluku Barat Daya | 122 | 127 | 127 | 122 | 121 | 123 | 100% | 95% | 97% | 0·95 (0·74-1·23) | 0·97 (0·75-1·24) |
| Buru Selatan | 81 | 27 | 70 | 81 | 22 | 59 | 100% | 81% | 84% | 0·81 (0·51-1·3) | 0·84 (0·6-1·18) |
| Kota Ambon | 1556 | 773 | 1663 | 1556 | 650 | 984 | 100% | 84% | 59% | 0·84 (0·77-0·92) | 0·59 (0·55-0·64) |
| Kota Tual | 216 | 196 | 176 | 216 | 196 | 146 | 100% | 100% | 83% | 1 (0-0) | 0·83 (0·67-1·02) |
| **North Maluku** | | | | | | | | | | | |
| Halmahera Barat | 201 | 185 | 186 | 201 | 185 | 179 | 100% | 100% | 96% | 1 (0-0) | 0·96 (0·79-1·18) |
| Halmahera Tengah | 81 | 42 | 99 | 81 | 42 | 74 | 100% | 100% | 75% | 1 (0-0) | 0·75 (0·55-1·02) |
| Kepulauan Sula | 111 | 89 | 118 | 111 | 81 | 113 | 100% | 91% | 96% | 0·91 (0·68-1·21) | 0·96 (0·74-1·24) |
| Halmahera Selatan | 419 | 290 | 339 | 419 | 265 | 288 | 100% | 91% | 85% | 0·91 (0·78-1·07) | 0·85 (0·73-0·99) |

| District | Number of cases | | | Number of cases on treatments | | | Treatment coverage | | | Treatment coverage ratio for 2020 vs 2019 (95% CI) | Treatment coverage ratio for 2021 vs 2019 (95% CI) |
|---|---|---|---|---|---|---|---|---|---|---|---|
| | 2019 | 2020 | 2021 | 2019 | 2020 | 2021 | 2019 | 2020 | 2021 | | |
| Halmahera Utara | 368 | 235 | 388 | 368 | 204 | 279 | 100% | 87% | 72% | 0·87 (0·73-1·03) | 0·72 (0·62-0·84) |
| Halmahera Timur | 137 | 62 | 85 | 137 | 55 | 77 | 100% | 89% | 91% | 0·89 (0·65-1·21) | 0·91 (0·69-1·2) |
| Pulau Morotai | 89 | 71 | 166 | 89 | 61 | 95 | 100% | 86% | 57% | 0·86 (0·62-1·19) | 0·57 (0·43-0·76) |
| Pulau Taliabu | 37 | 23 | 35 | 37 | 23 | 35 | 100% | 100% | 100% | 1 (0-0) | 1 (0-0) |
| Kota Ternate | 530 | 450 | 674 | 530 | 420 | 465 | 100% | 93% | 69% | 0·93 (0·82-1·06) | 0·69 (0·61-0·78) |
| Kota Tidore Kepulauan | 249 | 149 | 221 | 249 | 136 | 144 | 100% | 91% | 65% | 0·91 (0·74-1·12) | 0·65 (0·53-0·8) |
| **West Papua** | | | | | | | | | | | |
| Fakfak | 227 | 158 | 253 | 227 | 152 | 228 | 100% | 96% | 90% | 0·96 (0·78-1·18) | 0·9 (0·75-1·08) |
| Kaimana | 211 | 111 | 144 | 211 | 108 | 120 | 100% | 97% | 83% | 0·97 (0·77-1·23) | 0·83 (0·67-1·04) |
| Teluk Wondama | 109 | 71 | 125 | 109 | 68 | 90 | 100% | 96% | 72% | 0·96 (0·71-1·3) | 0·72 (0·55-0·95) |
| Teluk Bintuni | 302 | 202 | 276 | 302 | 133 | 198 | 100% | 66% | 72% | 0·66 (0·54-0·81) | 0·72 (0·6-0·86) |
| Manokwari | 758 | 519 | 699 | 758 | 501 | 598 | 100% | 97% | 86% | 0·97 (0·86-1·08) | 0·86 (0·77-0·95) |
| Sorong Selatan | 196 | 45 | 82 | 196 | 35 | 70 | 100% | 78% | 85% | 0·78 (0·54-1·11) | 0·85 (0·65-1·12) |
| Sorong | 434 | 142 | 269 | 434 | 142 | 228 | 100% | 100% | 85% | 1 (0-0) | 0·85 (0·72-0·99) |
| Raja Ampat | 110 | 67 | 53 | 110 | 66 | 44 | 100% | 99% | 83% | 0·99 (0·73-1·34) | 0·83 (0·59-1·18) |
| Tambrauw | NA | NA | NA | NA | NA | NA | NA | NA | NA | NA | NA |
| Maybrat | NA | NA | NA | NA | NA | NA | NA | NA | NA | NA | NA |
| Manokwari Selatan | 25 | 39 | 51 | 25 | 35 | 43 | 100% | 90% | 84% | 0·9 (0·54-1·5) | 0·84 (0·52-1·38) |
| Pegunungan Arfak | NA | NA | NA | NA | NA | NA | NA | NA | NA | NA | NA |
| Kota Sorong | 592 | 334 | 474 | 592 | 303 | 440 | 100% | 91% | 93% | 0·91 (0·79-1·04) | 0·93 (0·82-1·05) |
| **Papua** | | | | | | | | | | | |
| Merauke | 1010 | 937 | 818 | 1010 | 845 | 690 | 100% | 90% | 84% | 0·9 (0·82-0·99) | 0·84 (0·77-0·93) |
| Jayawijaya | 386 | 199 | 406 | 386 | 176 | 298 | 100% | 88% | 73% | 0·88 (0·74-1·06) | 0·73 (0·63-0·85) |
| Jayapura | 844 | 633 | 950 | 844 | 633 | 787 | 100% | 100% | 83% | 1 (0-0) | 0·83 (0·75-0·91) |
| Nabire | 1285 | 991 | 1133 | 1285 | 897 | 1040 | 100% | 91% | 92% | 0·91 (0·83-0·99) | 0·92 (0·85-1) |

| District | Number of cases | | | Number of cases on treatments | | | Treatment coverage | | | Treatment coverage ratio for 2020 vs 2019 (95% CI) | Treatment coverage ratio for 2021 vs 2019 (95% CI) |
|---|---|---|---|---|---|---|---|---|---|---|---|
| | 2019 | 2020 | 2021 | 2019 | 2020 | 2021 | 2019 | 2020 | 2021 | | |
| Kepulauan Yapen | 549 | 236 | 280 | 549 | 167 | 66 | 100% | 71% | 24% | 0·71 (0·6-0·84) | 0·24 (0·19-0·3) |
| Biak Numfor | 789 | 497 | 541 | 789 | 473 | 459 | 100% | 95% | 85% | 0·95 (0·85-1·07) | 0·85 (0·76-0·95) |
| Paniai | 443 | 383 | 378 | 443 | 381 | 351 | 100% | 99% | 93% | 0·99 (0·87-1·14) | 0·93 (0·81-1·07) |
| Puncak Jaya | 46 | 50 | 18 | 46 | 50 | 18 | 100% | 100% | 100% | 1 (0-0) | 1 (0-0) |
| Mimika | 1792 | 1524 | 1910 | 1792 | 1471 | 1723 | 100% | 97% | 90% | 0·97 (0·9-1·03) | 0·9 (0·84-0·96) |
| Boven Digoel | 360 | 302 | 343 | 360 | 286 | 328 | 100% | 95% | 96% | 0·95 (0·81-1·11) | 0·96 (0·82-1·11) |
| Mappi | 1067 | 1086 | 1158 | 1067 | 1085 | 1140 | 100% | 100% | 98% | 1 (0·92-1·09) | 0·98 (0·91-1·07) |
| Asmat | 399 | 384 | 275 | 399 | 361 | 258 | 100% | 94% | 94% | 0·94 (0·82-1·08) | 0·94 (0·8-1·1) |
| Yahukimo | 97 | 176 | 192 | 97 | 154 | 174 | 100% | 88% | 91% | 0·88 (0·68-1·13) | 0·91 (0·71-1·16) |
| Pegunungan Bintang | 62 | 60 | 68 | 62 | 59 | 68 | 100% | 98% | 100% | 0·98 (0·69-1·4) | 1 (0-0) |
| Tolikara | NA | NA | NA | NA | NA | NA | NA | NA | NA | NA | NA |
| Sarmi | 103 | 56 | 40 | 103 | 56 | 37 | 100% | 100% | 93% | 1 (0-0) | 0·93 (0·64-1·35) |
| Keerom | 108 | 145 | 120 | 108 | 38 | 118 | 100% | 26% | 98% | 0·26 (0·19-0·37) | 0·98 (0·76-1·28) |
| Waropen | 64 | 29 | 27 | 64 | 29 | 27 | 100% | 100% | 100% | 1 (0-0) | 1 (0-0) |
| Supiori | 65 | 26 | 71 | 65 | 16 | 55 | 100% | 62% | 77% | 0·62 (0·36-1·06) | 0·77 (0·54-1·11) |
| Mamberamo Raya | 41 | 23 | 40 | 41 | 22 | 38 | 100% | 96% | 95% | 0·96 (0·57-1·61) | 0·95 (0·61-1·48) |
| Nduga | NA | NA | NA | NA | NA | NA | NA | NA | NA | NA | NA |
| Lanny Jaya | 12 | 12 | 12 | 12 | 12 | 11 | 100% | 100% | 92% | 1 (0-0) | 0·92 (0·4-2·08) |
| Mamberamo Tengah | NA | NA | NA | NA | NA | NA | NA | NA | NA | NA | NA |
| Yalimo | 19 | 6 | 6 | 19 | 6 | 6 | 100% | 100% | 100% | 1 (0-0) | 1 (0-0) |
| Puncak | 16 | 15 | 6 | 16 | 15 | 2 | 100% | 100% | 33% | 1 (0-0) | 0·33 (0·08-1·35) |
| Dogiyai | 18 | 10 | 17 | 18 | 7 | 17 | 100% | 70% | 100% | 0·7 (0·29-1·67) | 1 (0-0) |
| Intan Jaya | NA | NA | NA | NA | NA | NA | NA | NA | NA | NA | NA |
| Deiyai | NA | NA | NA | NA | NA | NA | NA | NA | NA | NA | NA |
| Kota Jayapura | 2137 | 1536 | 2203 | 2137 | 1256 | 1640 | 100% | 82% | 74% | 0·82 (0·76-0·88) | 0·74 (0·7-0·79) |

**Supplementary Table 4. Multivariable analysis of factors associated with decrease in tuberculosis case notification rate, decrease in treatment coverage, and increase in mortality rate in Indonesia 2021**

| | Case notification rate | | Treatment coverage | | Mortality rate | |
|---|---|---|---|---|---|---|
| | **aOR (95% CI)** | **p** | **aOR (95% CI)** | **p** | **aOR (95% CI)** | **p** |
| COVID-19 incidence rate | | | | | | |
| Quartile 1 | 1 (reference) | | 1 (reference) | | **3·2 (1·0-9·8)** | **0·045** |
| Quartile 2 | 1·1 (0·5-2·3) | 0·748 | 0·8 (0·4-1·5) | 0·457 | 2·0 (1·0-4·2) | 0·206 |
| Quartile 3 | 0·9 (0·4-2·0) | 0·793 | 0·8 (0·4-1·9) | 0·642 | 1·8 (1·6-8·2) | 0·248 |
| Quartile 4 | 0·8 (0·3-2·1) | 0·686 | 1·0 (0·2-2·9) | 0·999 | 1 (reference) | |
| COVID-19 mortality rate | | | | | ·· | ·· |
| Quartile 1 | 1 (reference) | | 1 (reference) | | | |
| Quartile 2 | 1·2 (0·6-2·4) | 0·702 | 1·2 (0·6-2·4) | 0·681 | | |
| Quartile 3 | 0·9 (0·4-1·9) | 0·738 | 1·6 (0·7-3·7) | 0·251 | | |
| Quartile 4 | 1·2 (0·5-3·1) | 0·690 | 1·3 (0·5-3·6) | 0·561 | | |
| TB GeneXpert service per 100,000 population | | | | | ·· | ·· |
| Quartile 1 | 1·6 (0·7-3·3) | 0·247 | **0·4 (0·2-0·9)** | **0·021** | | |
| Quartile 2 | 1·2 (0·6-2·4) | 0·559 | 0·7 (0·3-1·4) | 0·282 | | |
| Quartile 3 | 1·3 (0·7-2·4) | 0·413 | 1·0 (0·5-1·9) | 0·995 | | |
| Quartile 4 | 1 (reference) | | 1 (reference) | | | |
| TB microscopy service per 100,000 population | ·· | ·· | | | ·· | ·· |
| Quartile 1 | | | 2·5 (0·9-6·7) | 0·077 | | |
| Quartile 2 | | | 1·6 (0·6-3·9) | 0·319 | | |
| Quartile 3 | | | 1·9 (0·9-4·1) | 0·101 | | |
| Quartile 4 | | | 1 (reference) | | | |
| Primary health centres per 100,000 population | | | | | | |
| Quartile 1 | 1·7 (0·7-4·0) | 0·218 | **3·5 (1·0-11·9)** | **0·042** | 3·1 (0·9-10·8) | 0·075 |
| Quartile 2 | 1·2 (0·6-2·1) | 0·799 | 1·1 (0·4-3·1) | 0·796 | **3·2 (1·0-9·6)** | **0·044** |
| Quartile 3 | 1·3 (0·6-2·1) | 0·791 | 1·1 (0·5-2·6) | 0·773 | 1·8 (0·6-5·3) | 0·291 |
| Quartile 4 | 1 (reference) | | 1 (reference) | | 1 (reference) | |
| Doctors per 100,000 population | ·· | ·· | | | ·· | ·· |
| Quartile 1 | | | 0·9 (0·4-2·1) | 0·885 | | |
| Quartile 2 | | | 0·8 (0·4-1·8) | 0·677 | | |
| Quartile 3 | | | 0·9 (0·5-1·8) | 0·781 | | |
| Quartile 4 | | | 1 (reference) | | | |
| Public health development index | ·· | ·· | | | ·· | ·· |
| Quartile 1 | | | 2·1 (0·8-5·6) | 0·144 | | |
| Quartile 2 | | | 1·3 (0·6-2·9) | 0·524 | | |
| Quartile 3 | | | 1·1 (0·6-2·3) | 0·738 | | |
| Quartile 4 | | | 1 (reference) | | | |
| Per capita domestic expenditure, USD | | | | | | |
| Quartile 1 | | | 1·0 (0·5-2·1) | 0·931 | 1·2 (0·5-3·2) | 0·707 |
| Quartile 2 | | | 1·6 (0·7-3·7) | 0·278 | 2·2 (0·8-5·9) | 0·136 |
| Quartile 3 | | | 2·0 (0·7-5·6) | 0·164 | 0·8 (0·2-2·9) | 0·747 |
| Quartile 4 | | | 1 (reference) | | 1 (reference) | |
| Formal education | ·· | ·· | | | ·· | ·· |
| Quartile 1 | | | 1 (reference) | ·· | | |
| Quartile 2 | | | 0·7 (0·4-1·4) | 0·297 | | |
| Quartile 3 | | | 1·0 (0·5-2·1) | 0·945 | | |
| Quartile 4 | | | **2·9 (1·2-7·2)** | **0·021** | | |

Province was treated as the random effect variable.
aOR: adjusted odds ratio· CI: confidence interval.
·· the variable did not enter the multivariable model therefore statistics were not estimated.
